# Relationships between increasing pre-load meal size, post-prandial acyl ghrelin, LEAP2, insulin, glucose, appetite, and food intake in adults without obesity

**DOI:** 10.64898/2026.07.29.26359169

**Authors:** Tasya Parastika, Jiaxun Liu, Raghav Bhargava, Wai In Ng, Jialin Guo, Marcela Rodriguez Flores, Xinyi Zhang, Mimoza Emini, Wanqian Li, Sandra Luur, Zoe Barley, Jeff Brunstrom, Anthony P Goldstone

## Abstract

The satiety cascade includes post-prandial increases in plasma glucose and insulin, and changes in appetitive gut hormones, including decreases in orexigenic stomach-derived ghrelin, and increases in liver-foregut-derived LEAP2 hormone, a ghrelin antagonist. However, which contribute to post-prandial attenuation of eating behaviour with increasing preload meal size is unclear.

In a randomised, single-blinded study, adults without obesity attended four visits, consuming 750mL liquid preloads 0, 600, 900, 1200 kcal, with assay of plasma glucose, acyl ghrelin (AG), LEAP2 and serum insulin over 0-3h with appetite ratings (n=17 participants, 65 visits). At 2h a virtual portion size creation task measured desired food intake (n=15, 58 visits, after data exclusion), and at 3h an *ad libitum* meal measured actual food intake (n=11, 43 visits).

The greater the pre-load meal size, the greater the post-prandial increase in plasma glucose and serum insulin, and greater the decrease in plasma AG and AG/LEAP2 ratio, associated with greater attenuation of eating behaviour (appetite ratings, desired/actual food intake). The strongest correlations of eating behaviour with blood measures were for serum insulin, plasma AG and AG/LEAP2 ratio, with weaker results for plasma glucose. Although there was a weak correlation of post-prandial plasma LEAP2 with pre-load meal size but not with eating behaviour.

Post-prandial increases in serum insulin and decreases in plasma AG and AG/LEAP2 ratio may be part of the satiety cascade related to preload meal size in adults without obesity. Future studies should investigate potential mediation of satiety in this context via other appetitive gut hormones, and in obesity.

## 1. INTRODUCTION

Eating behaviour is governed by a complex interplay of biological, psychological, and environmental factors that collectively ensure energy intake aligns with physiological requirements. Humans have evolved integrated systems to maintain energy balance, enabling food intake to meet the demands of health, growth, and metabolic function. These mechanisms span overlapping homeostatic and hedonic dimensions, reflecting the dual necessity of sustaining physiological needs while responding to the rewarding properties of food. It is importance to understanding these processes amidst rising global rates of obesity and diet-related disease, since dysregulation of appetite may contribute to excessive energy intake (Garutti et al., 2025).

### 1.1. Satiety cascade

In addition to longer-term weight changes, as part of homeostatic control of eating behaviour, the brain needs to be able to monitor the timing and nature, both energy intake and macronutrient composition, of food intake to regulate subsequent food intake. In the fed compared to the fasted state, there is reduced hunger and appetite, increased fullness, and reduced food appeal and cue reactivity, especially for high-energy foods, and *ad libitum* food intake (Fuhrer et al., 2008; Goldstone et al., 2009; Cameron et al., 2014; Goldstone et al., 2014; Bhargava et al., 2023).

The satiety cascade i s a biopsychological system regulating eating behaviour after food intake through three levels: psychological events (hunger perception, cravings, hedonic sensations); peripheral physiology and metabolic events; and neurotransmitter and metabolic interactions in the brain (Blundell, 1991).

The peripheral mediators of this satiety cascade are not clearly established and appear to include vagal innervation from stomach stretching, post-absorption nutrients including plasma glucose, and a variety of pancreatic-gut appetitive hormones (Blundell et al., 2010). The latter includes decreases in post-prandial orexigenic ghrelin from stomach (Muller et al., 2015), and post-prandial increases in satiety hormones PYY and GLP-1 from the intestine, and insulin from the pancreas. More recently identified is the potential importance of post-prandial increases in the novel satiety hormone liver-expressed antimicrobial peptide-2 (LEAP2) from liver, duodenum and jejunum, a hormone that is an inverse agonist at the ghrelin receptor (GHSR), and reciprocally regulated to ghrelin (Krause et al., 2003; Ge et al., 2018; Mani et al., 2019; Tolle et al., 2024).

For more fine-tuned monitoring of previous food intake, it would be important for there to be the ability to respond to different sizes of pre-load meals. It is well established that the greater the pre-load meal size, the greater the post-prandial fullness and lower post-prandial appetite ratings, and lower subsequent *ad libitum* food intake in people without and/or with obesity (Blom et al., 2005; le Roux et al., 2005). This will also be influenced by energy density and macronutrient composition, food texture and matrix and sensory-specific satiety of pre-load, and the presented portion size of *ad libitum* meals (Rolls, 1986).

Greater caloric content of pre-load meals results in greater post-prandial decreases in plasma total ghrelin in adults without obesity (though not in obesity where ghrelin is supressed), and acyl ghrelin (AG) in normal weight (le Roux et al., 2005; Ishii et al., 2016), and greater post-prandial increases in PYY, GIP, insulin and glucose in normal weight and obesity (Service et al., 1983; Ebert & Creutzfeldt, 1989; Blom et al., 2005; le Roux et al., 2006). There is a suggestion that the greater the pre-load meal size between 337-730 kcal, the greater the post-prandial increase in plasma LEAP2, but this is from analysis across nine different studies without and with obesity, with pre-load meals of different macronutrient composition, and different time points of blood collection between 60-150 min (Emini et al., 2024).

However, although these various blood biomarkers of satiety have been postulated to mediate the *physiological* effects of pre-load meal size on attenuation of human eating behaviour, this has not been previously examined directly through correlational analyses between the post-prandial changes in the potential mediator and eating behaviour (de Graaf et al., 2004; Flint et al., 2007; Horner et al., 2020). Indeed, under normal physiological meal sizes, post-prandial increases in satiety hormones such as GLP-1 and PYY may be insufficient to alter eating behaviour in the context of functional foods, unlike after gastric bypass surgery where increases are much higher (Lim & Poppitt, 2019; Lim et al., 2023; Hengist et al., 2024).

Instead, this role for appetitive gut hormones on human satiety has been generally progressed from exogenous administration of gut hormones, usually achieving supraphysiological plasma concentrations, correlations of endogenous plasma hormone concentrations with eating behaviour after single (rather than increasing) meal sizes, and rarely through administration of hormone antagonists (Steinert et al., 2017). Indeed, administration of the GLP-1 antagonist, exendin(9-39) has minimal effects on post-prandial human appetite ratings and no effects on *ad libitum* food intake in healthy adults without obesity, questioning the physiological importance of post-prandial GLP-1 in the satiety cascade in adults without obesity, at least after the 200-452 kcal preload meal sizes in these studies (see review (Gasbjerg et al., 2021) for individual papers).

### 1.2. Glucose

The evidence that post-prandial physiological increases in plasma glucose influence eating behaviour are limited (de Graaf et al., 2004; Flint et al., 2006; Flint et al., 2007; Horner et al., 2020). Many studies of exogenous intravenous glucose infusion achieve supraphysiological plasma glucose concentrations, there is a greater (‘incretin’) effect of intraduodenal vs. intravenous glucose administration to supresses appetite implicating other mediators e.g. insulin, GLP-1 (de Graaf et al., 2004; Schultes et al., 2016), while correlational studies implicate dynamic changes in plasma glucose, especially later post-prandial decreases, to be more important than absolute concentrations, in meal initiation (de Graaf et al., 2004; Wyatt et al., 2021).

### 1.3. Insulin

There is some evidence for post-prandial insulin contributing to attenuation of human eating behaviour, especially in normal weight, though this has not been studied in the context of mediation of effects of different pre-load meal sizes.

A comprehensive meta-analysis of human studies examining post-prandial insulin and appetite responses suggested that insulin may contribute to the enhancement of satiety across individuals with normal weight, overweight and obesity (Flint et al., 2007). However, whether insulin mediates the relationship between meal size and satiety remains unclear, as most studies have assessed appetite responses following either single fixed-calorie oral preload meals (Flint, 1998; Flint et al., 2001; Verdich et al., 2001; Asmar et al., 2010), meals differing simultaneously in energy content and glycaemic index (Speechly & Buffenstein, 2000; Bakhoj et al., 2003; Flint et al., 2007), or meals matched for energy but varying in macronutrient composition (Holt et al., 1996; Flint et al., 2007). However, increasing rates of *intraduodenal* glucose infusions in normal weight and overweight, did proportionally reduce *ad libitum* energy intake that was negatively correlated with plasma insulin (Pilichiewicz et al., 2007).

Studies using exogenous insulin administration with hyperinsulinaemic clamps have either shown increases in hunger under euglycaemic state in normal weight (Rodin et al., 1985), or no effects on appetite ratings, nor desired or *ad libitum* food intake under euglycaemic or hyperglycaemic states in normal weight or adults without obesity (Woo et al., 1984; Chapman et al., 1998; Gielkens et al., 1998; Belfort-DeAguiar et al., 2016).

Consequently, the independent contribution of insulin to meal size-dependent changes in satiety has yet to be fully established.

### 1.4. Ghrelin

Ghrelin is an orexigenic peptide hormone composed of 28 amino acids, synthesized and secreted by enteroendocrine cells primarily within the gastric fundus, existing in both unacylated/desacylated and acylated forms (Kojima & Kangawa, 2006; Romero et al., 2010; Stengel et al., 2011; Muller et al., 2015). The conversion of desacyl ghrelin (DAG) to its biologically active acyl ghrelin (AG) form is facilitated by the enzyme ghrelin O-acyltransferase. Only AG i s active at its growth hormone secretagogue receptor (GHSR). Ghrelin acts to increase growth hormone secretion, stimulate hepatic glucose production, diminish insulin secretion, and accelerate gastric emptying. stimulate appetite and food consumption, as part of the physiological response to fasting and weight loss where plasma AG increases (Briggs et al., 2013; Muller et al., 2015; Nymo et al., 2017; Coutinho et al., 2018; Nymo et al., 2018; Lyngstad et al., 2019; Peos et al., 2021; Ragland & Malin, 2023), though actions on GHSR on the vagus nerve, and several brain regions including the hypothalamus, hippocampus and midbrain dopaminergic system (Muller et al., 2015; Morris et al., 2018; Schulz et al., 2023).

In previous studies of adults without obesity, increasing pre-load meals size between 250-3000 kcal led to proportional reductions in plasma total ghrelin, though not in obesity where plasma ghrelin was suppressed, but pre-load macronutrient composition was not totally consistent (le Roux et al., 2005). Furthermore, pre-load meal sizes between 200-800 kcal proportionately lowered plasma AG over 4 hours (Ishii et al., 2016).

The evidence that AG plays a *physiological* role in regulation of human eating behaviour, especially post-prandially is contradictory. AG administration when fed certainly increases appetite ratings, *ad libitum* food intake and food cue reactivity in the brain reward system in humans without and with obesity, but this is with markedly supraphysiological plasma AG concentrations (Wren et al., 2001; Druce et al., 2005; Malik et al., 2008; Goldstone et al., 2014; Muller et al., 2015; Han et al., 2018). However, lower dose AG infusions in normal weight achieving physiological and moderately supraphysiological plasma AG concentrations did not alter hunger or satiety ratings nor increase *ad libitum* food intake (Lippl et al., 2012)

However, in a recent meta-analysis there was a positive correlation of endogenous plasma ghrelin with hunger ratings, that was stronger when measuring AG as opposed to DAG or total ghrelin (AG + DAG), but the effect was weaker in those with higher BMI (Anderson et al., 2023), where plasma ghrelin is lower (Wang et al., 2022). Interestingly, a positive correlation was not seen when examining studies in the period ≤3 hours after food intake, questioning a role for *post-prandial* changes in plasma ghrelin in mediating changes in hunger. Furthermore, studies in this meta-analysis have not included pre-load meal sizes varying markedly in caloric content with identical macronutrient composition (Anderson et al., 2023). Similarly, studies correlating post-prandial changes in plasma AG with changes in other ratings including appetite, fullness, volume able to eat and pleasantness to eat have also given highly variable results, again likely contributed to by differences in BMI status, which ghrelin hormone is being measured, and post-prandial timepoint and outcome measures (Mackelvie et al., 2007; Unick et al., 2010; Martens et al., 2012; Gibbons et al ., 2013; Deighton et al., 2014; Liu et al., 2015; Andarini et al., 2017; Nymo et al., 2018; Bhargava et al., 2023; Andreoli et al., 2024a). Furthermore, correlations of post-prandial AG with *ad libitum* food intake also show variable results, with very few demonstrating positive correlation (Unick et al., 2010; Gibbons et al., 2013; St-Onge et al., 2014; Roth et al., 2022; Bhargava et al ., 2023). Previous studies demonstrating the greater reduction in post-prandial total ghrelin and AG with increasing meal size have not related this to changes in eating behaviour (le Roux et al., 2005; Ishii et al., 2016). Furthermore, correlational evidence does not demonstrate causality, and AG may serve other roles such as influencing food reward and memory (Perello & Dickson, 2015; Suarez et al., 2019).

### 1.5. LEAP2

The recently identified hormone LEAP2) a 40-amino-acid peptide, predominantly synthesized in the liver, duodenum and jejunum, functions as an endogenous antagonist and inverse agonist to AG at the constitutively active GHSR (Krause et al., 2003; Ge et al., 2018; Cornejo et al., 2019; M’Kadmi et al., 2019; Mani et al., 2019; Wang et al., 2019; Islam et al., 2020; Fernandez et al., 2022; Islam et al., 2022; Lugilde et al., 2022).

LEAP2 is modulated in a reciprocal fashion relative to AG, as its plasma concentrations are lowered by fasting or weight loss and elevated by food intake in both humans and murine models, and in obesity, and so the plasma AG/LEAP2 ratio may be a critical determinant of human eating behaviour and metabolic processes (Mani et al., 2019; Bhargava et al., 2023; Emini et al ., 2024). There is a suggestion that post-prandial increases in plasma LEAP2 may be greater with increasing pre-load meal size, but this has only been analysed across multiple human studies with single size pre-loads, with differing macronutrient compositions, post-prandial blood time points, and mixture of BMI status (Bhargava et al., 2023; Emini et al., 2024).

LEAP2 administration blocks the ability of exogenous AG to stimulate food intake in pre-clinical models, and acute administration of LEAP2 alone to rodents (Ge et al., 2018; Cornejo et al., 2019; M’Kadmi et al., 2019; Islam et al., 2020; Hagemann et al., 2022; Hola et al., 2022; Islam et al., 2022; Shankar et al., 2024), while *Leap2* gene knockout mice have increased food intake (Shankar et al., 2021). LEAP2 infusions in men without and with obesity achieving supraphysiological plasma LEAP2 concentrations decreased *ad libitum* meal food intake (Hagemann et al., 2022; Englund et al., 2026), confirming for the first time a physiological role for GHSR signalling to increase human eating behaviour.

Correlational studies of plasma LEAP2 or AG/LEAP2 ratio with appetite ratings and *ad libitum* food intake have often been supportive of physiological roles for plasma LEAP2 in suppressing human eating behaviour but results have been varied, depending on whether examined in fasting or post-prandial state (though only studied after single size pre-loads), the nature of the pre-load and BMI status (Bhargava et al., 2023; Holm et al., 2023; Andreoli et al., 2024a; Andreoli et al., 2024b; Emini et al., 2024).

Furthermore, quantifying actual energy intake within controlled laboratory environments may present significant challenges due to factors such as bias, psychological stress, and the restriction of food options during *ad libitum* test meals. An alternative methodology involves assessing desired food consumption through a computer-mediated virtual portion creation task (VPCT), which has been empirically validated among individuals of normal weight as well as those who have undergone bariatric surgical interventions (Hamm et al., 2020; Hamm et al., 2021), and also examined in context of hyperinsulinaemic euglycaemic clamps (Gielkens et al., 1998). Such VPCT tasks allow participants to choose their preferred portion sizes of a wider variety of foods, thereby enabling a broader evaluation of appetite.

### 1.6. Hypotheses and Aims

This study hypothesised that increasing meal size consumed at a preload results in proportional post-prandial: (i) increases in plasma glucose, serum insulin, plasma LEAP2, and decreases in plasma AG and AG/LEAP2 ratio; (ii) decreases in appetite ratings; (iii) decreases in desired and actual food intake; (iv) post-prandial changes in glucose and hormones are related to the post-prandial attenuation of eating behaviour; and as an exploratory analysis (v) investigate within each outcome which post-prandial calculations give the strongest correlations.

The aims of this study were therefore to examine in a single-blinded cross-over study of healthy adults without obesity, correlations (using linear mixed model analysis) between: (a) pre-load meal size (0, 600, 900, 1200 kcal in a fixed volume), (b) post-prandial plasma glucose, serum insulin, plasma LEAP2, AG and AG/LEAP2 ratio, (c) post-prandial hunger, composite appetite, food craving and fullness visual analogue scale (VAS) ratings, (d) desired food intake using a VPCT at two hours after pre-load, and (e) actual food intake using *ad libitum* meal at three hours after pre-load. Food intake measures were examined as both absolute kcal intake and relative intake as % of estimated resting energy expenditure. Furthermore, exploratory analyses investigated differences in these correlations when using absolute, delta, and absolute area-under-curve (AUC) values for blood measures and appetite ratings in addition to the pre-registered incremental AUC (iAUC) measure.

## 2. METHODS

### 2.1. Ethics

Ethics approval for the participants was obtained from West London & GTAC Research Ethics Committee (23/LO/0510). This study was operated in accordance with the principles of the Declaration of Helsinki, and all participants provided informed consent. The study was pre-registered https://clinicaltrials.gov/study/NCT06013592.

### 2.2. Participant Recruitment and Screening Visit

Participants recruited were healthy adults between the ages of 18 and 60 years without obesity (i.e. body mass index (BMI) 18.0-29.9 kg/m^2^). Recruitment was facilitated through public advertisements, social media, Imperial College and hospital websites and noticeboards. Upon completion of an online brief questionnaire to confirm preliminary eligibility, potential participants had a telephone screening to inform them about the study and to verify their eligibility.

Eligible participants then underwent an in-person health assessment screening visit to confirm eligibility and practised the computer-based tasks (virtual portion size creation task, food spatial memory task) before participating in the four experimental study visits. Inclusion and exclusion criteria are given in Supplementary Table S1. Screening blood measurements included full blood count, renal function, electrolytes, liver function tests, thyroid function test, HbA1c, iron studies and urinary pregnancy test for females. Questionnaires about mood, eating behaviour and alcohol use were completed (see Supplementary Methods).

### 2.3. Study Design and Procedure

This study was a single-blind, randomised, within-participant, experimental medicine study conducted at Imperial College Research Facility, Imperial College London. After a screening visit, participants attended four more study visits lasting 4.5 hours, at least one day apart, in a randomised (using a computer-generated algorithm) cross-over, placebo-controlled design after an overnight fast. Participants were told to avoid strenuous exercise and alcohol, and refrain from eating or drinking from 8pm on the day before the visit. Participants received either a 0, 600, 900, 1200 or 1800 kcal liquid fixed preload breakfast (730 kcal, 55% carbohydrate, 31% fat, 14% protein) at t=0 min, with a virtual portion size creation task (VPCT) to measure *desired* food intake at t=120min, and an *ad libitum* meal to measure actual food intake at t=180min (Figure 1). At t=125 min, participants completed a virtual spatial memory task for high-energy foods (different from foods presented at VPCT and *ad libitum* meal) and objects lasting 30 minutes (de Vries et al., 2020), the results of which will be published elsewhere.

**Figure 1.**
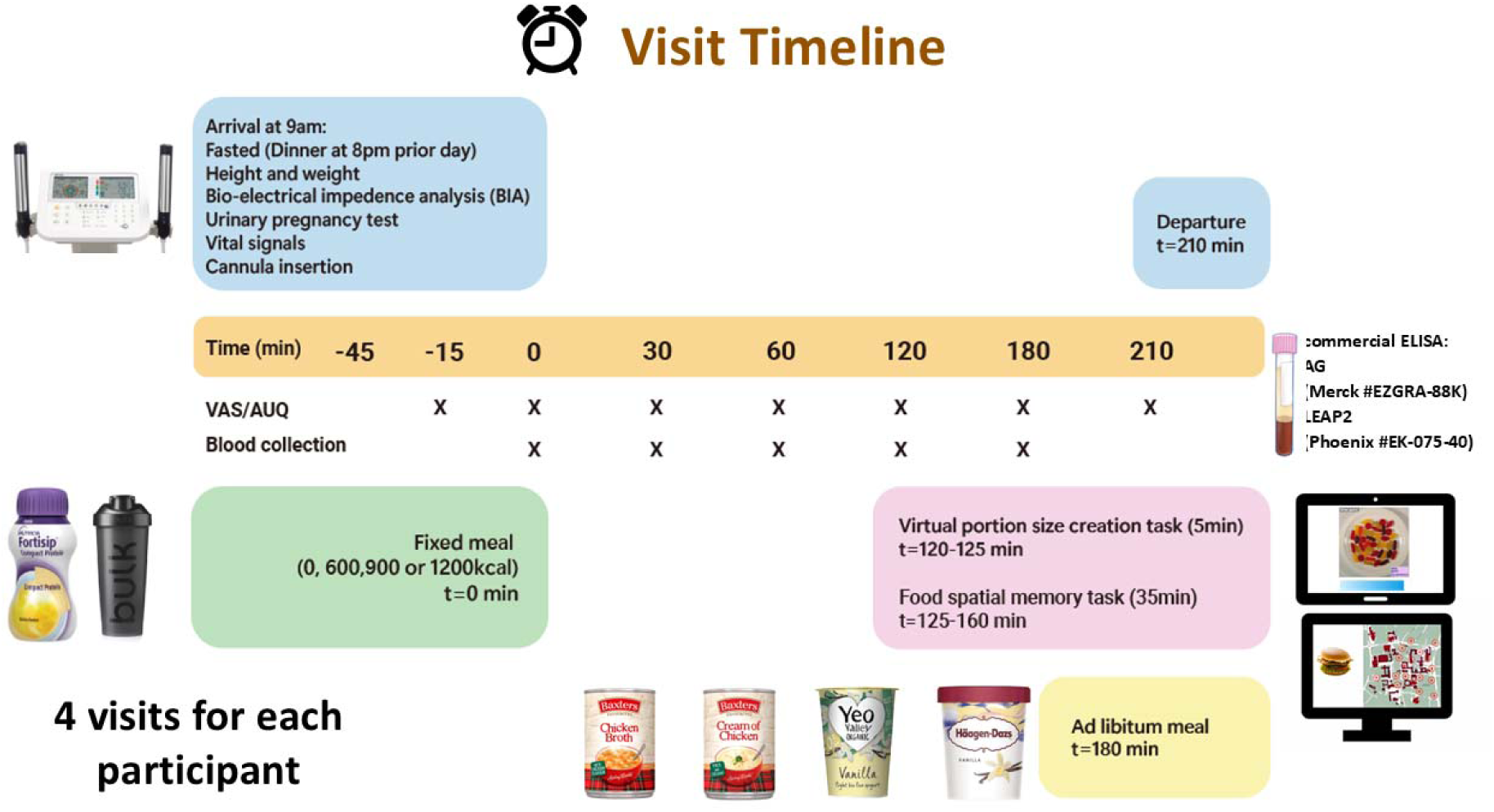
Schematic diagram of study visit timeline. After cannulation at t=+0min, participants received a fixed preload drink (750 mL) with 0, 600, 900 or 1200 kcal. Venous blood samples were collected through the cannula at t=0, 30, 60, 120 and 180 min for analysing fasting and post-prandial glucose and hormones, and visual analogue scale (VAS) ratings of hunger, composite appetite, food craving and fullness taken at -15, 0, 30, 60, 120, 180, 210 min. A five-minute virtual portion creation task (VPCT) was given at t=+120 min, followed by a food spatial memory task at t=+125min and *ad libitum* test meal at t=+180 min.

#### 2.3.1. Anthropometry

At each visit, height and weight were measured to determine body mass index (BMI) and bio-electrical impedance analysis (BIA) (MC-780MA, Tanita, Tokyo, Japan) used for determining % body fat and fat free mass (FFM). Resting energy expenditure (REE) for each participant was calculated by the Cunningham equation = 500 + 22*lean body mass (LBM), where LBM was equated to FFM.

#### 2.3.2. Fixed preload meal

Participants were blinded to the energy content of the preload liquid meal with different calorific values. To standardise effects of volume, all meals were offered in 750 mL portions in an opaque cup with a lid. The size of each preload meal was characterised both in terms of its absolute (kcal) and relative (kcal as % estimated REE) caloric value. After receipt, the meal was consumed within 10-15min.

#### 2.3.3. Blood tests

At t=0, 30, 60, 120 and 180min, 10mL blood samples were collected and assayed (Department of Clinical Biochemistry, Imperial College Healthcare NHS Trust) for plasma glucose and serum insulin (Alinity, Abbott GmbH, Wiesbaden, Germany). Homeostasis model assessment of insulin resistance (HOMA-IR) was calculated using the formula: fasting insulin (mU/L) x fasting glucose (mmol/L)/22.5 (Matthews et al., 1985). Blood was collected in lithium heparin tubes containing protease inhibitors aprotinin (Trasylol, Nordic Pharma Ltd., UK) and AEBSF (4-(2-aminoethyl) benzenesulfonyl fluoride hydrochloride) (A8456; Sigma-Aldrich, Dorset, UK), as previously described (Mani et al., 2019; Bhargava et al., 2023). Plasma samples were stored at -80°C for future measurement of gut hormones.

Plasma LEAP2 concentrations were measured in duplicate using a commercial enzyme-linked immunoassay (EK-075-40, Phoenix Pharmaceuticals Inc., CA, USA) according to the manufacturer’s instructions with plasma samples diluted 10 times in assay buffer (Mani et al., 2019; Bhargava et al., 2023), with inter-assay and intra-assay coefficients of variation 5.4% and 5.2% respectively for this study.

Plasma AG concentrations were measured in duplicate using a commercial enzyme-linked immunoassay (EZGRA-88K, Merck Millipore, Gillingham, Dorset, UK) according to the manufacturer’s instruction (Mani et al., 2019), with inter-assay and intra-assay coefficients of variation 9.5% and 7.2% respectively for this study.

#### 2.3.4. Visual analogue scale ratings

VAS ratings (0-100mm) were used to measure hunger, fullness, pleasantness to eat, and prospective food consumption (volume would eat), as well as aversive symptoms: sickness (nausea) and discomfort, at that moment (Goldstone et al., 2014). Composite appetite was calculated as [(hunger + pleasantness + volume + (100 - fullness) / 4] for data reduction and to minimise issues with multiple comparisons. Food craving (score 1-7) was measured by averaging two 7-point Likert scales asking participants how much they agree or disagreed with the following statements between “strongly disagree” to “strongly agree”: “All I want right now is something to eat”, “Nothing would be better than eating something right now”. These questions were adapted from overlapping questions assessing alcohol and cigarette craving using the Alcohol Urge Questionnaire (AUQ) and Brief Questionnaire of Smoking Urges (QSU-brief) (Tiffany & Drobes, 1991; Bohn et al., 1995). Within-participant post-prandial changes of composite appetite, hunger, food craving, fullness, nausea and discomfort were calculated using data from t=0, 30, 60, 120, 180 min at each visit to give incremental area under curve iAUC_0-2/3h_ using the trapezoid rule.

#### 2.3.5. Virtual portion creation task

At t=120min, participants performed a 5-minute virtual portion creation task (VPCT) to measure desired food intake for 16 dishes (savoury/sweet, low/high fat) modified from a previous study (Hamm et al., 2020; Hamm et al., 2021). Food pictures selected for this VPCT represented savoury or sweet foods with high-fat (energy from fat >33%, >4.0kcal/g) or low-fat (energy from fat <33%, <3.5kcal/g (**Supplementary Figure 1**). Desired food intake (absolute/relative) was averaged across the dishes for all foods. Desired food intake was examined as both *absolute* kcal and as *relative* kcal, expressed as % of estimated REE at that visit.

#### 2.3.6. Ad libitum test meal

At t=180min, participants received an excess amount of four dishes of different categories: low-fat savoury, high-fat savoury, low-fat sweet, high-fat sweet. Participants were blinded to energy content of these foods and instructed to “eat as much as you want”. All foods were individually presented in designated transparent glass bowls and weighed before and after ingestion to calculate actual food intake (absolute/relative), for total amount and by food category low-/high-fat, sweet/savoury. If they did not like tomato soup, chicken was provided as an alternative option. The nutritional composition of the dishes served is given in **Supplementary Table S2**. Total actual food intake was examined as both *absolute* kcal and as *relative* kcal, expressed as % of estimated REE at that visit.

### 2.4. Statistics

All statistical analyses were performed using IBM SPSS Statistics software version 29 (SPSS Inc., IBM, Chicago, USA) and GraphPad Prism v9.1.1 (GraphPad Software, San Diego, USA), with effects considered significant at P<0.05, with small effect sizes at pseudo-r^2^ values ≥0.04 and correlation coefficients r ≥0.20, and moderate effect sizes at pseudo-r^2^ values ≥0.25 and r ≥0.50. Demographic data were checked for normality using the Shapiro-Wilk test. For normally distributed data, mean ± standard deviation (SD) was reported, and otherwise, median and interquartile range (IQR).

#### 2.4.1. Power calculations

Assuming four study visits for each participant, a repeated measures ANOVA analysis using incremental AUC (iAUC_0-3hr_) for plasma hormone concentrations, n=15 has 80% power at P<0.05, to detect an effect size of 0.32 (moderate) for overall difference between visits, assuming correlation among repeated measures 0.5.

#### 2.4.2. Mixed model analysis

Correlations were examined using linear mixed model analysis of meal caloric size (absolute/relative), with post-prandial blood measures, and with eating behaviour from visual analogue scale (VAS) ratings, desired and actual food intake (using iAUC or iAUC) to determine the marginal value for pseudo-r^2^ as an estimate of the variance of the dependent variable explained by the covariate, along with mean ± SD for β parameter estimate.

A participant’s data was excluded from analysis of all visits for either desired or actual food intake respectively i f their average desired food intake or total actual food intake at 0 kcal pre-load visit was <20% estimated REE, since this likely indicated a dislike for the foods presented, to avoid floor effects. All corelations with pre-load meal size, and desired and actual food intake, were performed using both absolute intake (kcal) and relative intake (kcal as % of estimated REE).

For VPCT and *ad libitum* meal data, the impact of sex, and any moderating impact of sex on effect of preload meal size (sex x preload meal size interaction), was examined for desired and actual food intake, using either absolute or relative food intake data.

To examine changes in appetite or aversive symptom ratings and blood measures over time, linear mixed model analysis used time after pre-load and visit as within participant factors to report time x visit interaction, and if non-significant main effects of time and visit, with Greenhouse-Geisser correction. Post-hoc comparisons were reported (a) at each post-prandial time point relative to baseline 0 h within each visit, and (b) at each post-prandial time point relative to the 0 kcal visit between visits, using Dunnet’s correction for multiple comparisons.

Heatmaps summarise the results of correlations between outcome variables. The stronger the red the greater the positive correlation, and the stronger the blue, the greater the negative correlation, with the r value and direction given in the cell, and the superscript letters giving the P value. Benjamini-Hochberg false discovery rate (FDR) correction was also applied to correct for multiple comparisons.

#### 2.4.3. Exploratory analysis of alternative post-prandial outcomes

The preregistered analysis used iAUC_0-2/3h_ for post-prandial blood measures and appetite ratings in the correlations. To explore the potential stronger results with other post-prandial measures, correlations between pre-load meals size, post-prandial blood measures and appetite ratings were also performed using values from: 0h (as control to ensure no difference at baseline by chance), absolute value at 2/3h, change from baseline to 2/3h (Δ0-3h), absolute AUC (AUC_0-2/3h_), incremental AUC (iAUC_0-2/3h_), absolute minimum (hunger, composite appetite, food craving, plasma AG and AG/LEAP2 ratio) or maximum (fullness, plasma glucose, serum insulin, plasma LEAP2) value over 2/3 h excluding baseline value (min/max 2/3h), and change from baseline to minimum/maximum value (Δ0-min/max 2/3h).

## 3. RESULTS

### 3.1. Participant demographics

The total number of 17 participants had 68 visits, with two participants (N102 and N104, both females) missing their 900 kcal visit, and in addition one participant (N107, male) missing gut hormones from their 600 kcal visit. Detailed demographics including body composition, and average fasting plasma glucose and hormone concentrations are given in **Table 1**, while eating behaviour, mood and alcohol use questionnaires are given in **Supplementary Table S1**. Two participants were excluded from analysis of the desired food intake (VPCT), as average intake <20% estimated REE at 0 kcal visit, leaving 15 participants (8 male, 7 female), 58 visits (including the two participants N102 and N104 missing 900 kcal visits, with one participant N107 missing gut hormones for 600kcal visit). Six participants were excluded from analysis of actual food intake (*ad libitum* meal) as total intake was <20% estimated REE at 0 kcal visit, leaving 11 participants (7 male, 4 female), 43 visits (including one participant N104 missing 900 kcal visit, with one participant N107 missing gut hormones for 600 kcal visit).

**Table 1.** Participant characteristics. Categorical variables are presented as numbers and percentage of the cohort. Normally distributed data are shown as mean ± SD, otherwise as median [IQR], along with range (minimum to maximum). Abbreviations: BMI, body mass index; HOMA-IR, homeostatic model assessment for insulin resistance; IQR, interquartile range. ^a^ averaged across all study visits. ^b^ to convert glucose from mmol/L to mg/dL multiple by 18.02. ^c^ to convert insulin from mU/L to pmol/L multiple by 6.94. ^d^ to convert LEAP2 from ng/mL to pmol/L multiple by 218.3. ^e^ to convert acyl ghrelin from pg/mL to pmol/L multiply by 0.297.

|  |  |  |
| --- | --- | --- |
| N | 17 |  |
| Sex, n (%) |  |  |
| Male | 9 (53.0%) |  |
| Ethnicity, n (%) |  |  |
| Caucasian | 11 (64.7%) |  |
| Age (years) |  |  |
| Mean ± SD | 32.1 ± 10.8 | |
| Range | 20 - 55 |  |
| Height (m) <sup>a</sup> |  |  |
| Mean ± SD | 1.72 ± 0.1 | |
| Range | 1.6 - 1.9 |  |
| Weight (kg) <sup>a</sup> |  |  |
| Mean ± SD | 65.8 ± 9.0 | |
| Range | 48.0 - 83.1 |  |
| BMI (kg/m<sup>2</sup>) <sup>a</sup> |  |  |
| Median [IQR] | 21.2 [20.9, 23.4] |  |
| Range | 18.9 - 27.1 |  |
| Overweight status, n (%) |  |  |
| BMI ≥18.0, <25.0 kg/m<sup>2</sup> | 14 (82.4%) | |
| BMI ≥25.0 kg/m<sup>2</sup> | 3 (17.7%) | |
| Body fat (%) <sup>a</sup> | Female | Male |
| Mean ± SD | 25.9 ± 7.3 | 15.0 ± 5.0 |
| Range | 16.2 – 34.6 | 9.3 – 22.1 |
| HOMA-IR <sup>a</sup> |  |  |
| Median [IQR] | 0.7 [0.6, 0.9] |  |
| Range | 0.5 – 1.2 |  |
| Fasting plasma glucose (mmol/L) <sup>a,b</sup> |  |  |
| Mean ± SD | 4.7 ± 0.4 | |
| Range | 4.2 – 5.3 |  |
| Fasting serum insulin (mU/L) <sup>a,c</sup> |  |  |
| Median [IQR] | 3.5 [3.0, 4.0] |  |
| Range | 2.0 – 5.4 |  |
| Fasting plasma LEAP2 (ng/mL) <sup>a,d</sup> |  |  |
| Mean ± SD | 11.6 ± 2.1 | |
| Range | 7.4 – 16.1 |  |
| Fasting plasma acyl ghrelin (pg/mL) <sup>a,e</sup> |  |  |
| Mean ± SD | 368.0 ± 191.4 | |
| Range | 77.6 – 719.5 |  |
| Plasma acyl ghrelin/LEAP2 molar ratio <sup>a</sup> |  |  |
| Mean ± SD | 0.0463 ± 0.0290 | |
| Range | 0.0101 – 0.1252 |  |

### 3.2. Effects of pre-load meal size on post-prandial bloods

As expected, after consumption of a caloric pre-load (600-1200 kcal), there were significant *increases* in plasma glucose (**Figure 2A** for change from baseline, **Supplementary Figure S2A** for absolute), serum insulin (**Figure 2B, Supplementary Figure S2B**), and plasma LEAP2, though only significant after 1200 kcal meal (**Figure 2C, Supplementary Figure S2C**), and significant *decreases* in plasma AG (**Figure 2D, Supplementary Figure S2D**), and plasma AG/LEAP2 ratio (**Figure 2E, Supplementary Figure S2E**).

**Figure 2.**
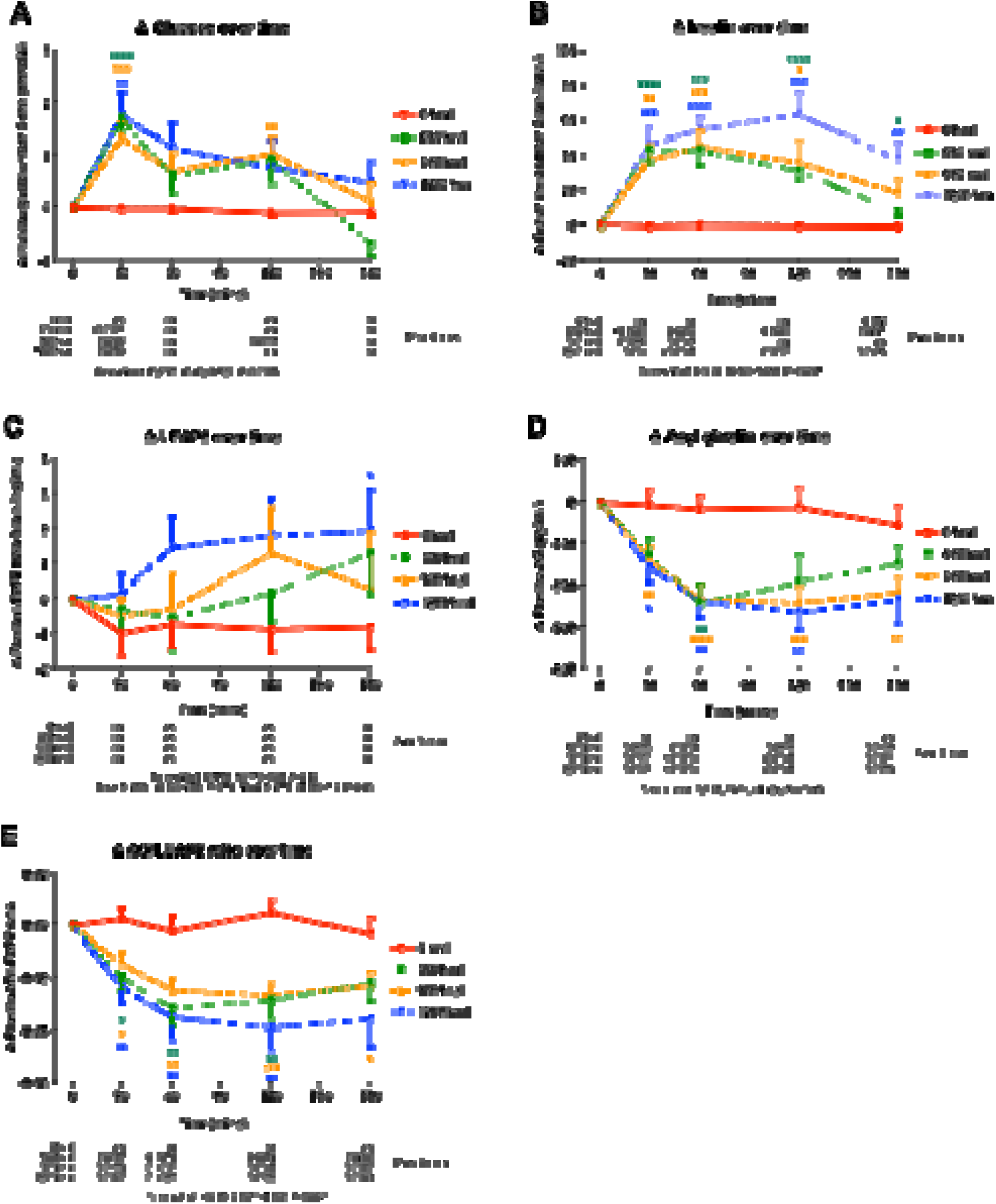
Changes in blood glucose and hormone concentrations over time at study visits. Change from baseline (0 min) in (A) plasma glucose, (B) serum insulin, plasma (C) LEAP2, (D) AG and (E) AG/LEAP2 molar ratio, at each of four study visits, with pre-load given at 0 min: 0 kcal (red circle, solid line), 600 kcal (green square, dotted line), 900 kcal (yellow triangle, dashed line), 1200 kcal (blue inverted triangle dashed-dotted line. Data expressed as mean ± SEM, n=17, n=66 visits (missing two 900 kcal visits), and for (C-E) n=65 visits (missing one 600 kcal visit). Statistical results from general linear model for interaction of time x visit given beneath graph. Coloured symbols within graph indicate P value for post-hoc Dunnet’s test vs. 0 kcal at each time point: *P<0.05, **P<0.01, ***P<0.005, ****P<0.001; numbers immediately below graph indicate P value for post-hoc Dunnet’s test vs. 0 min for each visit (with Greenhouse-Geisser correction). Abbreviations: ns, non-significant (P>0.05). To convert glucose from mmol/L to mg/dL multiple by 18.02; to convert insulin from mU/L to pmol/L multiple by 6.94.

When correlating iAUC_0-3h_ for glucose and hormones with absolute pre-load meal size (kcal) (**Figure 4**), there were significant *positive* correlations with plasma glucose (**Figure 3A**), serum insulin (**Figure 3B**), and plasma LEAP2 (**Figure 3C**) (i.e. the greater the pre-load meal size, the greater their post-prandial *increase*), and *negative* correlations with plasma AG (**Figure 3D**) and plasma AG/LEAP2 ratio (**Figure 3E**) (i.e. the greater the pre-load meal size, the greater their post-prandial *decrease*). Similar results were seen when correlating with relative pre-load meal size (kcal as % REE) (**Figure 4, Supplementary Figures S3A-E**).

**Figure 3.**
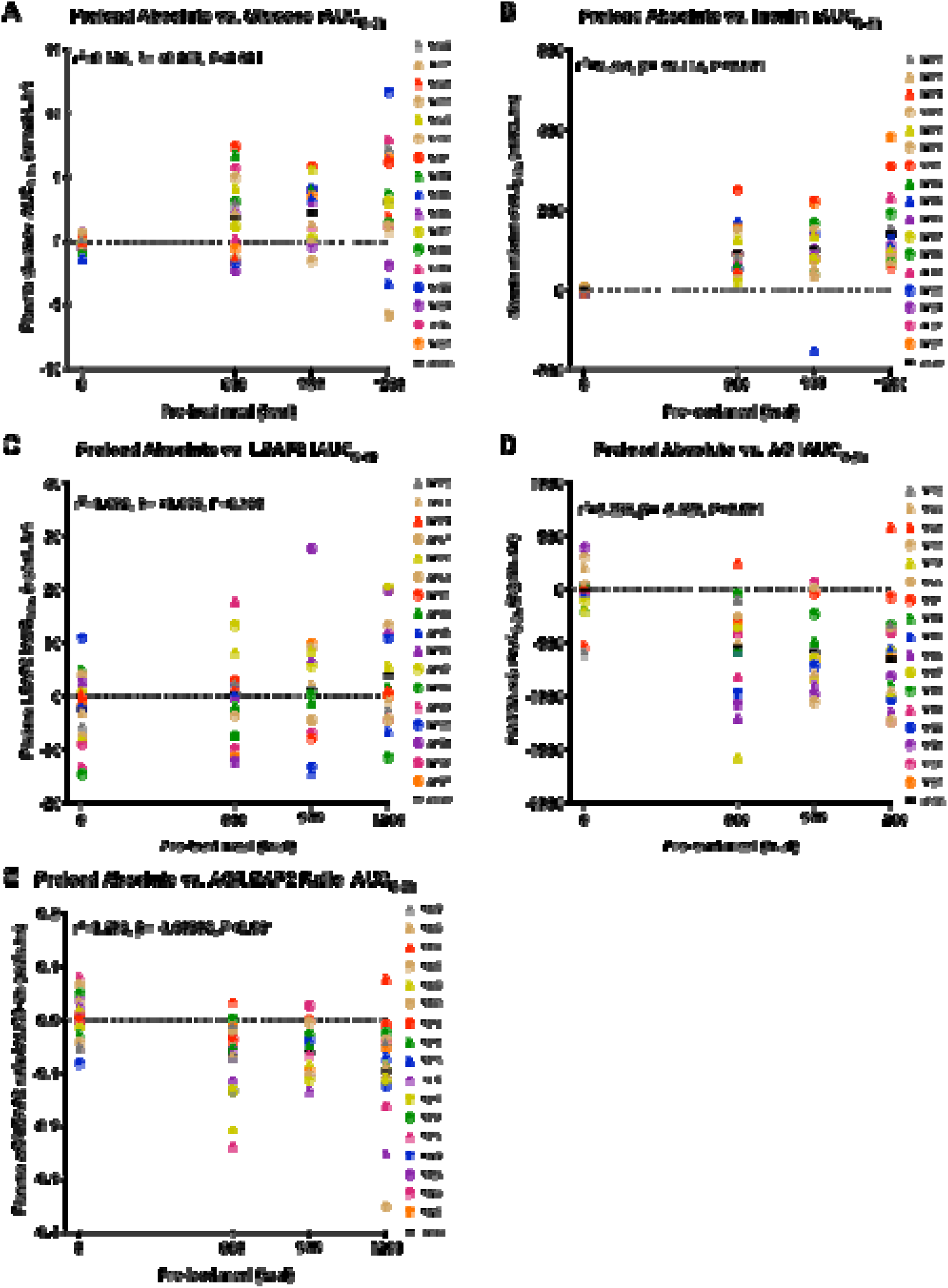
Correlations of absolute preload meal size with blood glucose and hormones. Correlations of increasing pre-load meal size over 4 study visits (0, 600, 900 and 1200 kcal) with (A) plasma glucose, (B) serum insulin, plasma (C) LEAP2, (D) AG and (E) AG/LEAP2 molar ratio as total iAUC_0-3h_, with marginal value of pseudo-R squared (r^2^), P value, and estimated mean of ß parameter from linear mixed model analysis. n=17 participants, n=66 visits (missing two 900 kcal visits), except for (C-E) n=65 visits (missing one 600 kcal visit). Males are represented as circles and females as triangles. Abbreviations: iAUC, incremental area under the curve. To convert glucose from mmol/L to mg/dL multiple by 18.02; to convert insulin from mU/L to pmol/L multiple by 6.94.

**Figure 4.**
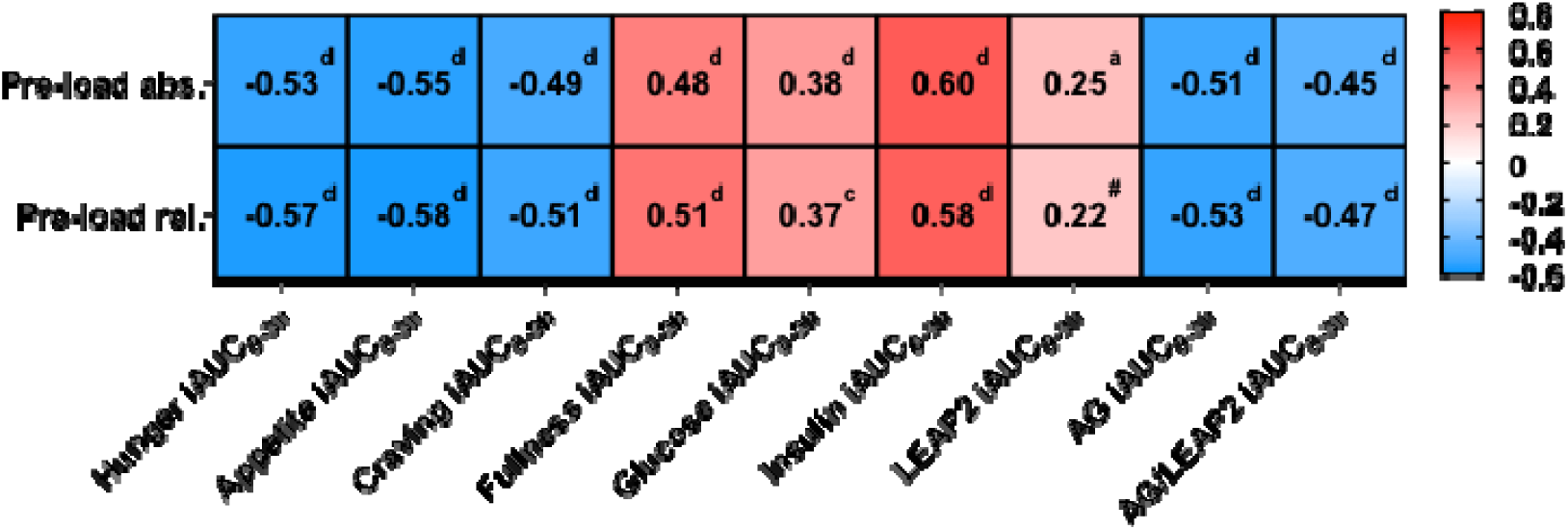
Summary heatmap for correlations of pre-load meal size with appetite ratings, blood glucose and hormones. The stronger the red, the greater the positive correlation, and the stronger the blue, the greater the negative correlation, with r value and direction given in the cell, with superscript letters giving P-value: ^#^ P<0.1, ^a^ P<0.05, ^b^ P<0.01, ^c^ P<0.005, ^d^ P<0.001; all letters P<0.05 using Benjamini-Hochberg false discovery rate (FDR) correction. Abbreviations: abs., absolute kcal; i AUC, incremental area under the curve; rel., relative kcal as % estimated resting energy expenditure. n=17 participants, n=66 visits (missing two 900 kcal visits), except LEAP2, AG and AG/LEAP2 ratio: n=17 participants, n=65 visits (missing one 600 kcal visit).

Effect sizes for correlations of blood measures with pre-load meal size were moderate for insulin and AG, and small for AG/LEAP2 ratio, glucose, LEAP2 (in descending order) (**Figure 4**).

### 3.3. Effects of pre-load meal size on post-prandial appetite ratings

As expected, after consumption of a caloric pre-load (600-1200 kcal), there were significant *decreases* in VAS ratings for hunger (**Figure 5A**), composite appetite (**Figure 5B**), and food craving (**Figure 5C**), and *increases* in fullness (**Figure 5D**).

**Figure 5.**
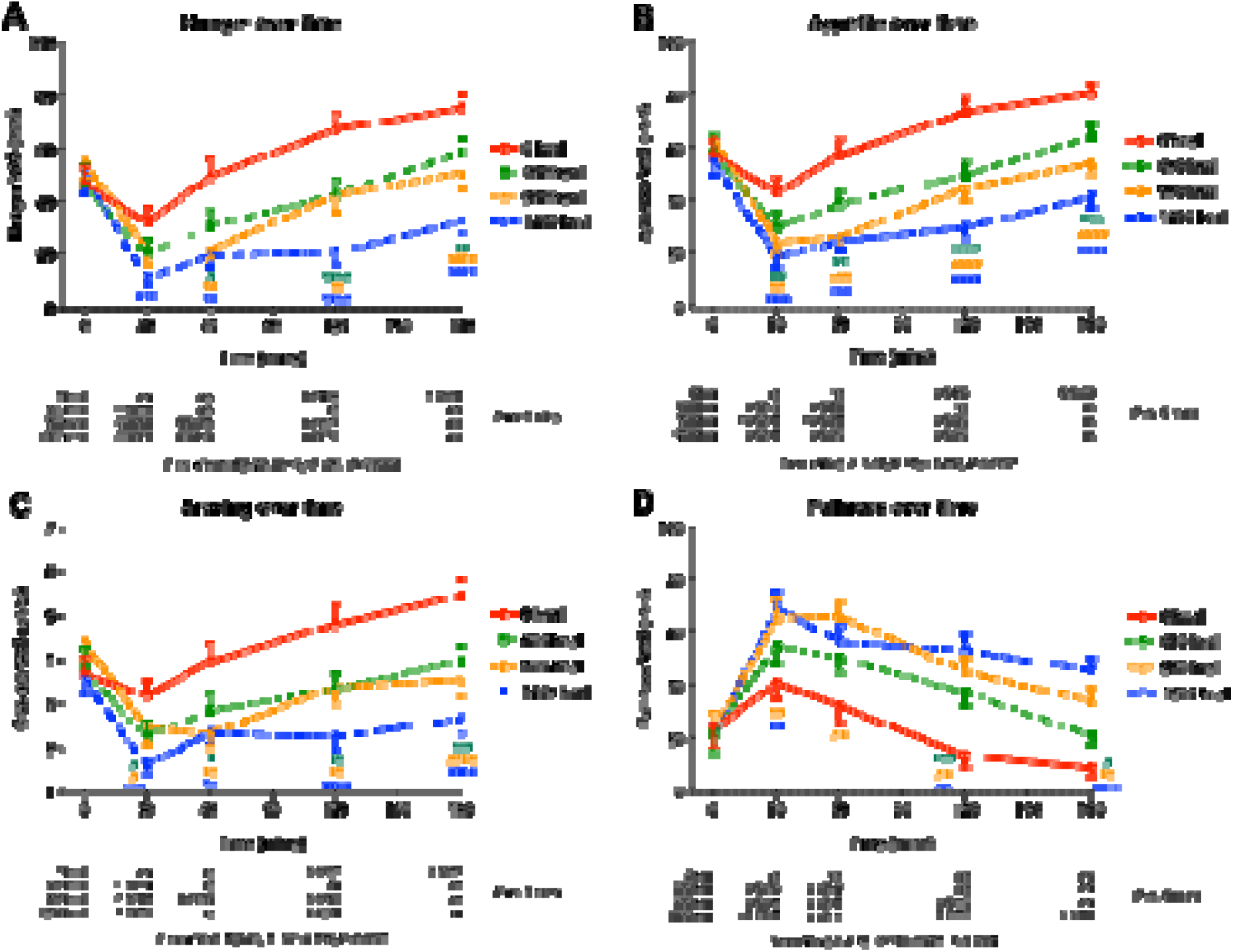
Visual analogue scale ratings of hunger, composite appetite, food craving and fullness. Absolute *v*isual analogue scale (VAS) ratings of (A) hunger (0-100mm), (B) composite appetite (0-100mm), (C) food craving (1-7 units), and (D) fullness (0-100 mm) at each of four study visits, with pre-load given at 0 min: 0 kcal (red circle, solid line), 600 kcal (green square, dotted line), 900 kcal (yellow triangle, dashed line), 1200 kcal (blue inverted triangle dashed-dotted line. Data expressed as mean ± SEM, n=17, n=66 visits (missing two 900 kcal visits), except for (C-E) n=65 visits (missing one 600 kcal visit). Statistical results from general linear model for interaction of time x visit given beneath graph. Coloured symbols within graph indicate P value for post-hoc Dunnet’s test vs. 0 kcal at each time point: *P<0.05, **P<0.01, ***P<0.005, ****P<0.001; numbers immediately below graph indicate P value for post-hoc Dunnet’s test vs. 0 min for each visit (with Greenhouse-Gei sser correction). Abbreviations: ns, non-significant (P>0.05).

When correlating iAUC_0-3h_ for VAS ratings with absolute pre-load meal size (kcal) (**Figure 4**), there were significant *negative* correlations with hunger (**Figure 6A**), composite appetite (**Figure 6B**), and food craving (**Figure 6C**) (i.e. the greater the pre-load meal size, the greater their post-prandial *decrease*), and *positive* correlations with fullness (**Figure 6D**) (i.e. the greater the pre-load meal size, the greater their post-prandial *increase*). Similar results were seen when correlating with relative pre-load meal size (kcal as % REE) (**Figure 4, Supplementary Figure S4A-D**). For both absolute and relative pre-load meal size, the order of correlation strength (based on r values) was appetite = hunger > food craving = fullness, but values were similar and generally of moderate effect size (**Figure 4**).

**Figure 6.**
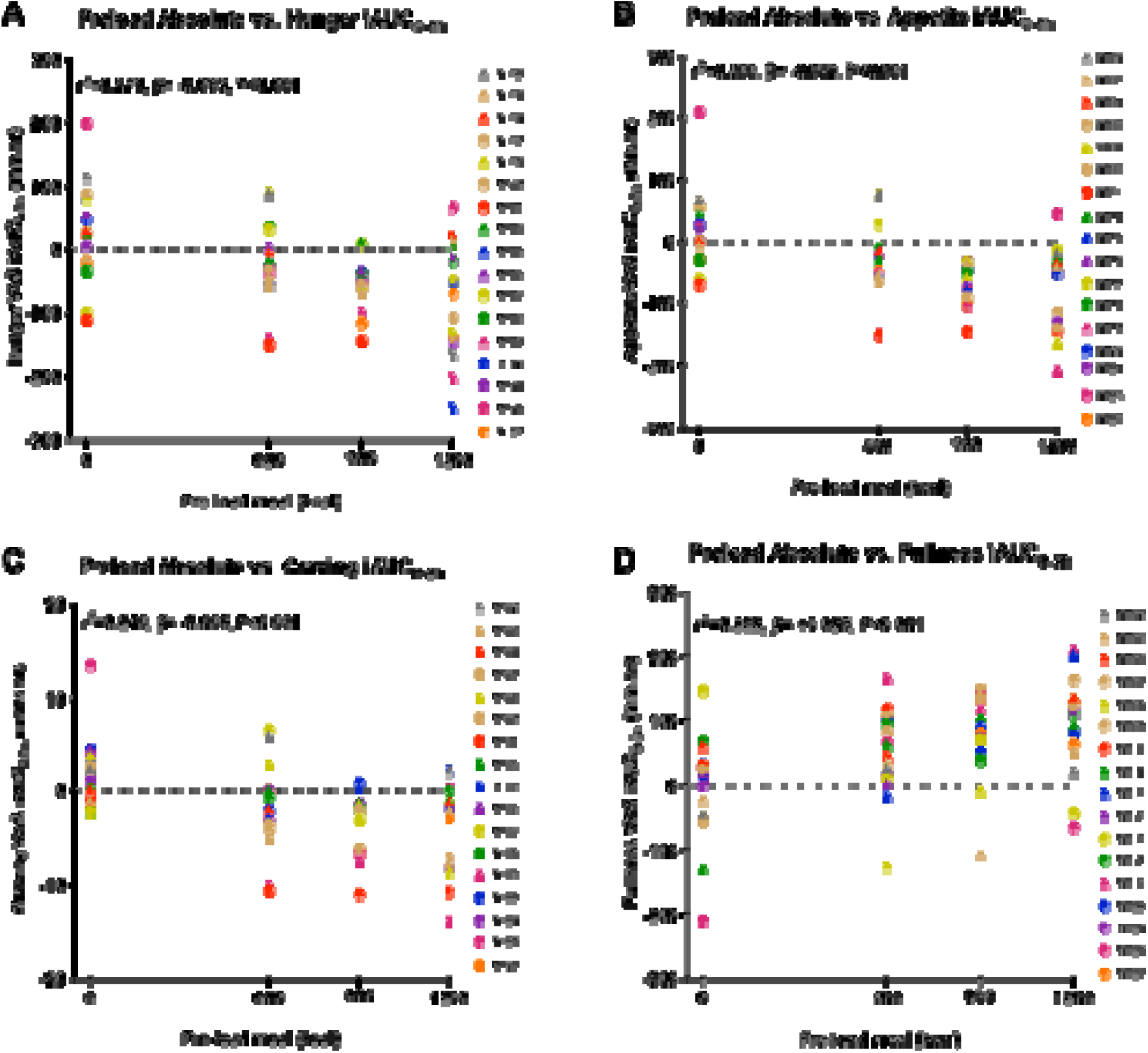
Correlations between absolute preload meal size and VAS appetite ratings. Correlations of absolute pre-load meal size over four study visits (0, 600, 900, 1200 kcal) with (A) hunger, (B) composite appetite, (C) food craving, and (D) fullness as total iAUC_0-3h_, with marginal value of pseudo-R squared (r^2^), P value, and estimated mean of parameter from linear mixed model analysis. n=17 participants, n=66 visits (missing two 900 kcal visits). Males are represented as circles and females as triangles. Abbreviations: iAUC, incremental area under the curve.

### 3.4. Effects of pre-load meal size on desired food intake

As expected, there were significant *negative* correlations between average *absolute* desired food intake from VPCT and *absolute* pre-load meal size (kcal) (**Figures 7A and 8**), i.e. the greater the pre-load meal size, the lower the desired food intake two hours later, with small effect size. A similar negative correlation was seen when using *relative* pre-load meal size and desired food intake (kcal as % REE) (**Figure 8, Supplementary Figure S4E**).

**Figure 7.**
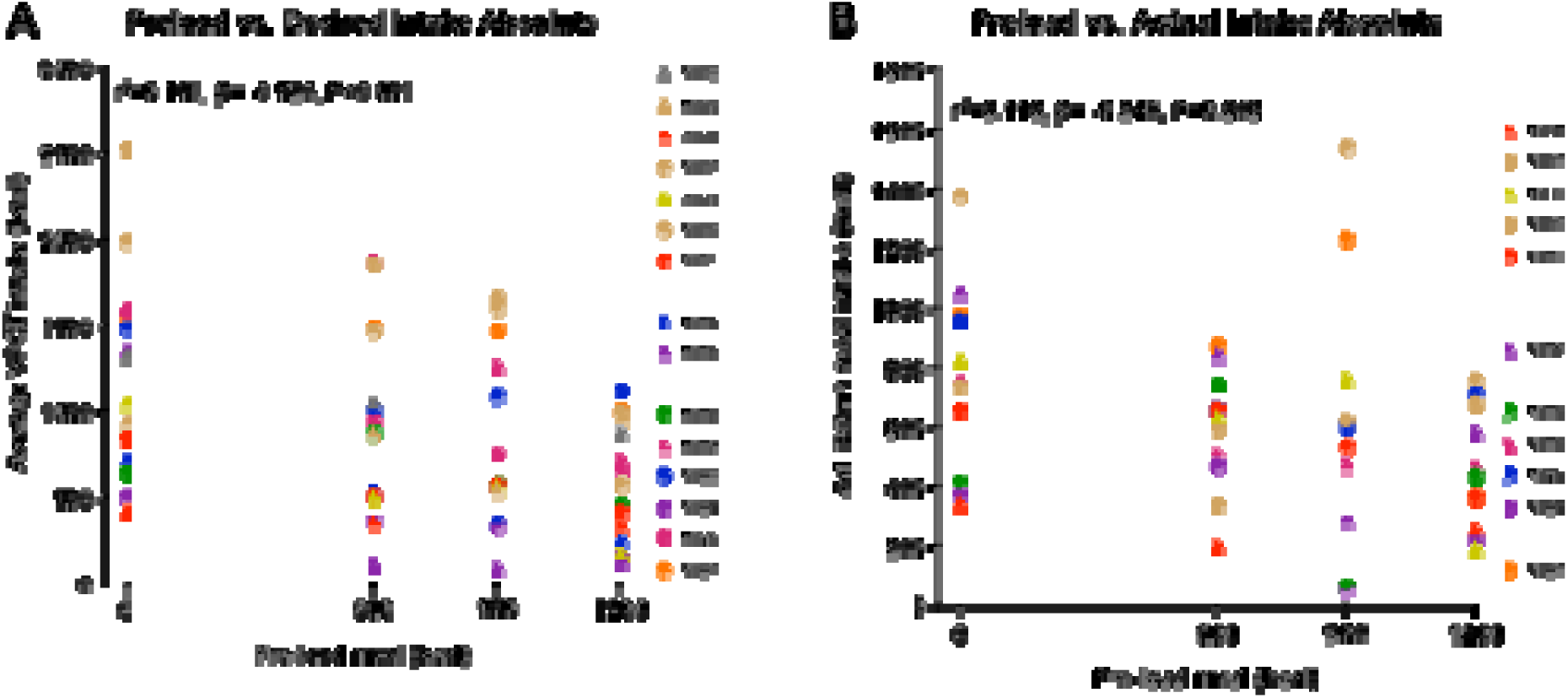
Correlations between absolute preload meal size and food intake. Correlations of absolute pre-load meal size over four study visits (0, 600, 900, 1200 kcal) with absolute (A) desired food intake from VPCT, and (B) actual food intake from *ad libitum* meal, with marginal value of pseudo-R squared (r^2^), P value, and estimated mean of parameter from linear mixed model analysis. (A) n=15 participants, n=58 visits (B) n=11 participants, n=43 visits. Males are represented as circles and females as triangles. Abbreviations: iAUC, incremental area under the curve.

**Figure 8.**
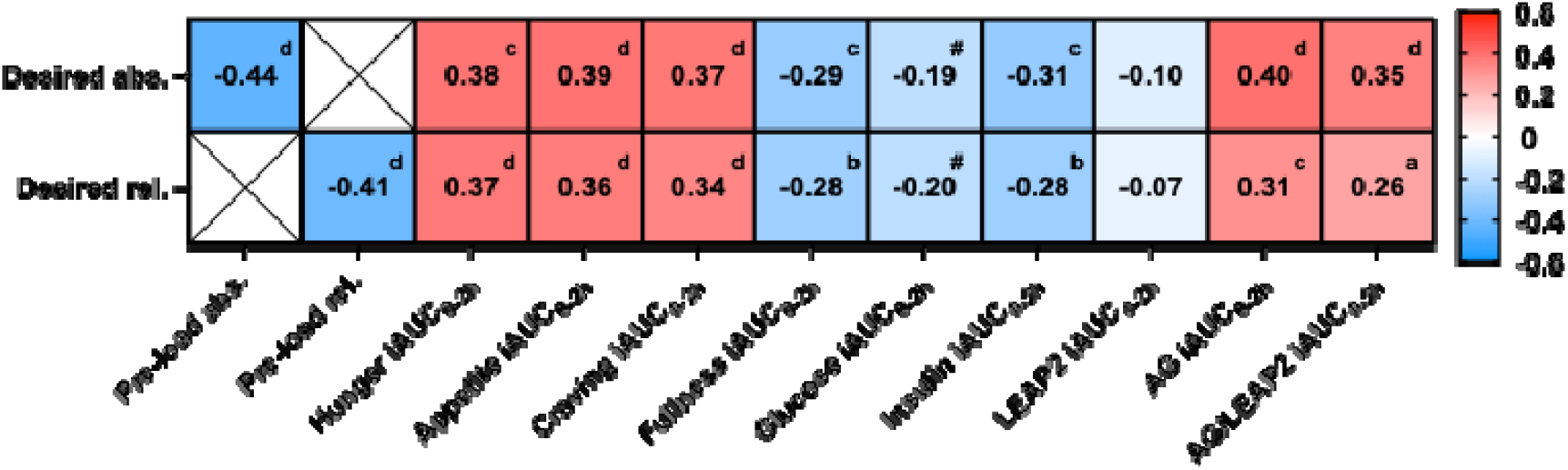
Summary heatmap for correlations of desired food intake with pre-load meal size, appetite ratings, glucose and hormones. The stronger the red, the greater the positive correlation, and the stronger the blue, the greater the negative correlation, with r value and direction given in the cell, with superscript letters indicating P-value: ^#^ P<0.1, ^a^ P<0.05, ^b^ P<0.01, ^c^ P<0.005, ^d^ P<0.001; all letters P<0.05 using Benjamini-Hochberg false discovery rate (FDR) correction. Using data from VPCT, n=15 participants, n=58 visits, except n=57 visits for LEAP2, AG and AG/LEAP2 ratio. Abbreviations: abs., absolute kcal; iAUC, incremental area under the curve; rel., relative kcal as % estimated resting energy expenditure.

### 3.5. Effects of pre-load meal size on actual food intake

As expected, there were significant *negative* correlations between total *absolute* actual food intake from the *ad libitum* meal and *absolute* pre-load meal size (kcal) (**Figures 7B and 9**), i.e. the greater the pre-load meal size, the lower their actual intake three hours later, with small effect size. A similar negative correlation was seen when using *relative* pre-load meal size and actual food intake (kcal as % REE) (**Figure 9, Supplementary Figure S4F**).

**Figure 9.**
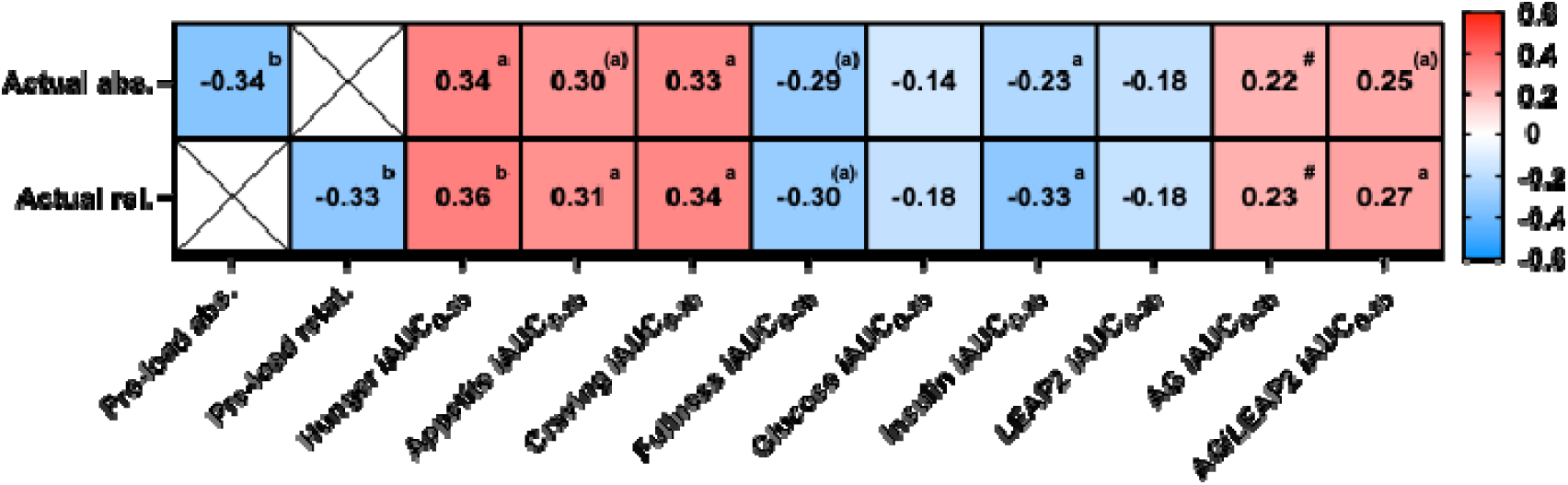
Summary heatmap for correlations of actual food intake with pre-load meal size, appetite ratings, glucose and hormones. The stronger the red, the greater the positive correlation, and the stronger the blue, the greater the negative correlation, with r value and direction given in the cell, with superscript letters indicating P-value: ^#^ P<0.1, ^a^ P<0.05, ^b^ P<0.01, ^c^ P<0.005, ^d^ P<0.001; letters in brackets indicate P>0.05 using Benjamini-Hochberg false discovery rate (FDR) correction. Using data from *ad libitum* meal, n=11 participants, 43 visits, except n=42 visits for LEAP2, AG and AG/LEAP2 ratio. Abbreviations: abs., absolute kcal; iAUC, incremental area under the curve; rel., relative kcal as % estimated resting energy expenditure.

*Desired* food intake at 2h was positively correlated with *actual* food intake at 3h whether expressed as absolute (kcal) or relative (kcal as % REE) intake with small-moderate effect size: r^2^=0.22 and 0.25 respectively, both P<0.001 (11 participants, 43 visits).

### 3.6. Effects of pre-load meal size on aversive symptoms

After consumption of any pre-load (0-1200 kcal), there were small *increases* in nausea and discomfort (though none reached statistical significance), but there were no significant differences between visits, indicating that these mild aversive symptoms were related to the volume consumed rather than the caloric content (**Supplementary Figure S5**). Furthermore, there were no significant correlations of pre-load meal size with nausea nor discomfort iAUC_0-3h_ (**Supplementary Figure S6**).

### 3.7. Relationships of post-prandial appetite ratings and bloods with desired food intake

When correlating iAUC_0-2h_ for VAS ratings with absolute desired intake (kcal) (**Figure 8**), there were significant *positive* correlations with hunger (**Supplementary Figure S7A**), composite appetite (**Supplementary Figure S7C**) and food craving (**Supplementary Figure S7E**) (i.e. the greater the post-prandial *decrease* in ratings, the *lower* their desired food intake), and *negative* correlations with fullness (i.e. the greater the post-prandial *increase* in fullness, the *lower* their desired food intake) (**Supplementary Figure S7F**). Similar results were seen when correlating with relative desired intake (kcal as % REE) (**Figure 8, Supplementary Figures S7B,D,F,H**). For both absolute and relative desired food intake, the order of correlation strength (based on r values) was: appetite = hunger > food craving > fullness, and only small effect size (**Figure 8**).

There were no significant correlations of iAUC_0-2h_ sickness nor discomfort with desired food intake (absolute or relative) (**Supplementary Figures S6 and S8A-D**).

When correlating iAUC_0-2h_ for glucose and hormones with absolute desired intake (kcal), there were significant *negative* correlations with plasma glucose (**Supplementary Figure S9A**) and serum insulin (**Supplementary Figure S9B**) (i.e. the greater the post-prandial *increase* in glucose and insulin, the *lower* their desired intake), but no significant correlation with plasma LEAP2 (**Supplementary Figure S9C**), and significant *positive* correlations with plasma AG (**Supplementary Figure S9D**) and plasma AG/LEAP2 ratio (Supplementary Figure S9E) (i.e. the greater the post-prandial *decrease* in hormones, the *lower* their desired intake). Similar results were seen when correlating with relative desired intake (kcal as % REE) (**Supplementary Figure S10**).

Across absolute and relative desired food intake, the order of correlation strength (based on r values) was: AG > AG/LEAP2 ratio > insulin > glucose > LEAP2, at best reaching only small effect sizes (**Figure 8**).

### 3.8. Relationships of post-prandial appetite ratings and bloods with actual food intake

When correlating iAUC_0-3h_ for VAS ratings with absolute actual intake (kcal) (**Figure 9**), there were significant *positive* correlations with hunger (**Supplementary Figure S11A**), composite appetite (**Supplementary Figure S11C**) and food craving (**Supplementary Figure S11E**) (i.e. the greater the post-prandial *decrease* in ratings, the *lower* their actual food intake), and *negative* correlations with fullness (i.e. the greater the post-prandial *increase* in fullness, the *lower* their actual food intake) (Supplementary Figure S11F). Similar results were seen when correlating with relative actual food intake (kcal as % REE) (**Figure 9, Supplementary Figures S11B,D,F,H**).Across absolute and relative actual food intake, the order of strength of correlations (based on r values) was: hunger > food craving > appetite = fullness, only reaching small effect sizes (**Figure 9**).

When correlating iAUC_0-3h_ aversive symptoms with actual food intake (absolute or relative), there were significant *negative* correlations with sickness (**Supplementary Figures S6 and S8E-F**) (i.e. the greater the post-prandial *increase* in sickness, the *lower* their actual intake), but not with discomfort (**Supplementary Figures S6 and S8G-H**). These negative correlations of actual food intake with sickness appeared to be driven by a few outliers (**Supplementary Figure 89E-F**).

When correlating iAUC_0-3h_ for glucose and hormones with absolute actual intake (kcal) (**Figure 9**), there was no significant *negative* correlation with plasma glucose (**Supplementary Figure S12A**), but a significant *negative* correlation with serum insulin (**Supplementary Figure S12B**) (i.e. the greater the post-prandial *increase* in insulin, the *lower* their actual intake), no significant correlation with plasma LEAP2 (**Supplementary Figure S12C**), trend for a significant *positive* correlation with plasma AG (**Supplementary Figure S12D**), and significant *positive* correlation plasma AG/LEAP2 ratio (**Supplementary Figure S12E**) (i.e. the greater the post-prandial *decrease* in AG and AG/LEAP2 ratio, the *lower* their actual intake). Similar results were seen when correlating with relative desired intake (kcal as % REE) (**Figure 9, Supplementary Figure S13**). For both absolute and relative desired food intake, the order of correlation strength (based on r values) was: AG/LEAP2 ratio > insulin = AG > LEAP2 > glucose, though at best reaching only small effect sizes (**Figure 9**).

### 3.9. Impact of sex on desired and food intake

There was no significant impact of sex on the relationship with pre-load meal size, or main effects of sex, for actual (kcal) or relative (kcal % REE) desired or actual food intake at the VPCT or *ad libitum* test meal (**Supplementary Table S3**).

### 3.10. Relationships of post-prandial bloods with appetite ratings

There were significant *negative* correlations of iAUC_0-3h_ hunger, composite appetite and food craving, with iAUC_0-3h_ plasma glucose (trend only for hunger) and serum insulin (**Figure 10, Supplementary Figures S14-S16A-B**), i.e. the greater the post-prandial *increase* in glucose and insulin, the greater the post-prandial *decrease* in appetite ratings. There was a significant *positive* correlation of fullness with serum insulin. i.e. the greater the post-prandial *increase* in insulin, the greater the post-prandial *increase* in fullness, but not with plasma glucose (**Figure 10, Supplementary Figures S17A-B**). These reached only small effects sizes.

**Figure 10.**
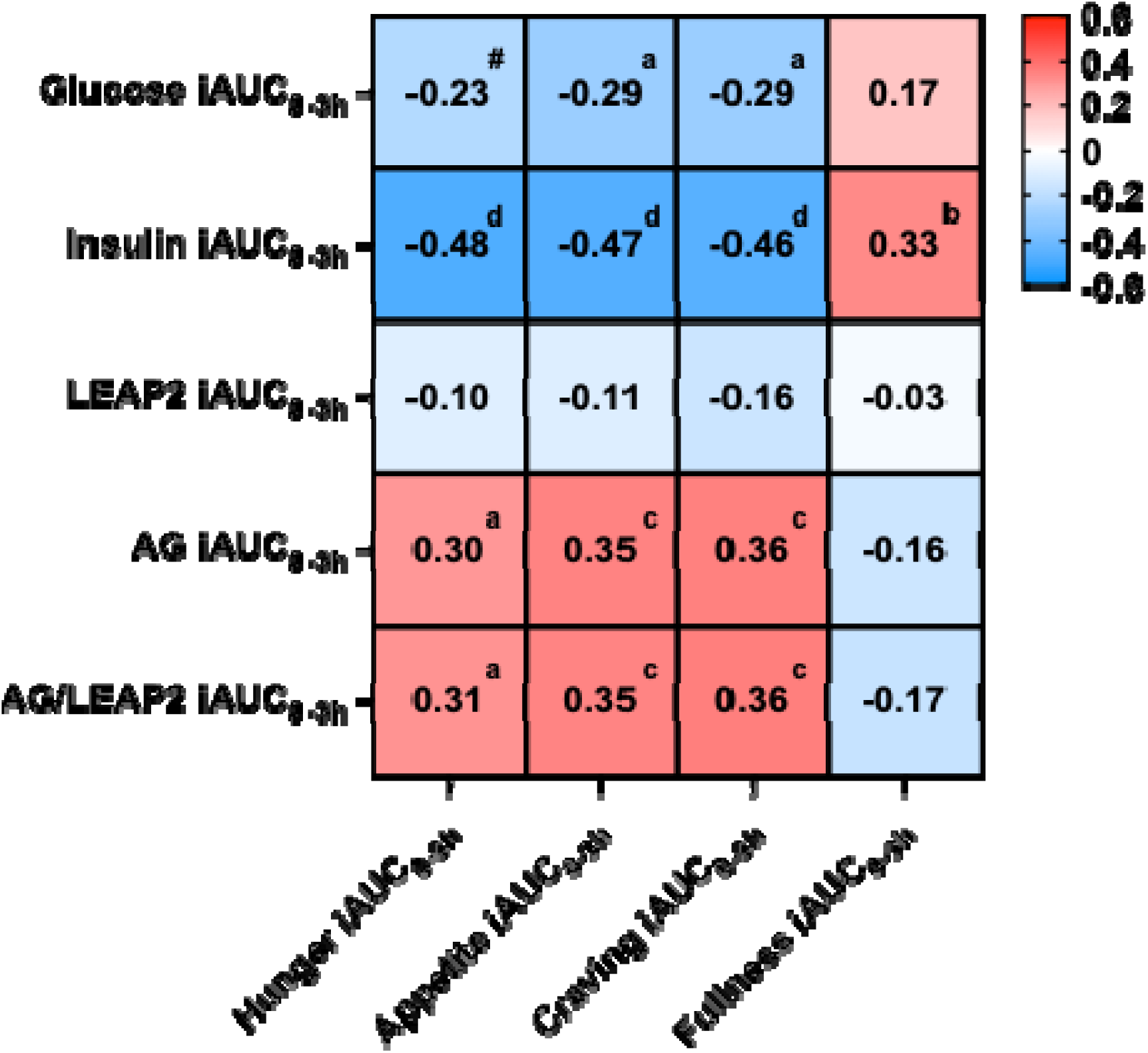
Summary heatmap for correlations of appetite ratings with glucose and hormones. The stronger the red, the greater the positive correlation, and the stronger the blue, the greater the negative correlation, with r value and direction given in the cell, with superscript letters indicating P-value: ^#^ P<0.1, ^a^ P<0.05, ^b^ P<0.01, ^c^ P<0.005, ^d^ P<0.001; all letters P<0.05 using Benjamini-Hochberg false discovery rate (FDR) correction. n=17 participants, n=66 visits, except n=65 visits for LEAP2, AG, AG/LEAP2 ratio. Abbreviations: abs., absolute kcal; iAUC, incremental area under the curve; rel., relative kcal as % estimated resting energy expenditure.

There were no significant correlations of iAUC_0-3h_ hunger, composite appetite, food craving, or fullness, with iAUC_0-3h_ plasma LEAP2 (**Figure 10, Supplementary Figures S14C, 15C, S16C, S17C**). However, there were significant *positive* correlations of iAUC_0-3h_ hunger, composite appetite and food craving with iAUC_0-3h_ plasma AG and AG/LEAP2 ratio (**Figure 10, Supplementary Figures S14D-E, S15D-E, S16D-E**), i.e. the greater the post-prandial *decrease* in AG and AG/LEAP2 ratio, the greater the post-prandial *decrease* in appetite ratings, that reached small effect sizes. However, there were no significant negative correlations of plasma AG nor AG/LEAP2 ratio with fullness (**Figure 10, Supplementary Figures S17D-E**).

### 3.11. Relationships between pre-load meal size and outcome measures using exploratory post-prandial calculations

Results are summarised below, with further details given in **Supplementary Results.**

Using exploratory post-prandial calculations for the outcome variables (namely 3h, Δ0-3h, AUC_0-3h_, iAUC_0-3h_, minimum/maximum between 0.5-3h, Δ 0-minimum/maximum 0.5-3h, or over 2 hours for desired food intake), the two strongest correlations rarely included iAUC_0-3h_. This was only found for correlation of *absolute* pre-load meal size with plasma LEAP2, and *relative* pre-load meal size with plasma AG and LEAP2 (**Supplementary Figure S18**).

For influence of pre-load meal size on VAS ratings and blood measures, the strongest correlations were generally with absolute value at 3h, AUC_0-3h_, and minimum/maximum 0.5-3h (**Supplementary Figure S18**).

Similarly, for relationships of *desired* food intake with VAS ratings and blood measures, the strongest correlations were generally with absolute value 2h, AUC_0-2h_, and minimum/maximum 0.5-2h (**Supplementary Figure S19**).

Furthermore, for relationships of *actual* food intake with VAS ratings and blood measures, the strongest correlations were generally with absolute value 3h, AUC_0-3h_, and minimum/maximum 0.5-3h (**Supplementary Figure S20**).

For correlations between VAS ratings and blood measures, the correlations for hunger were generally strongest using AUC_0-3h_, iAUC_0-3h_ and Δ 0-minimum/maximum 0.5-3h, for appetite absolute value 2h and AUC_0-3h_, for food craving absolute value 3h, AUC_0-3h_, iAUC_0-3h_ and Δ 0-minimum/maximum 0.5-3h, and for fullness absolute value 3h, AUC_0-3h_, minimum/maximum 0.5-3h and Δ 0-minimum/maximum 0.5-3h (**Supplementary Figure S21**).

There were no significant correlations between the change in plasma glucose from maximum value between 0.5-3h to value at 3h (to assess dynamic changes in glucose from peak) and *actual* food intake whether expressed as absolute or relative kcal (r=-0.21, P=0.14; r=-0.23, P=0.10 respectively).

## 4. DISCUSSION

In summary, this within-participant, randomised, single-blinded study, in adults without obesity consuming varying pre-load meal sizes (0-1200 kcal), found that:

i. greater pre-load meal size led to (a) as expected, proportionally greater post-prandial decreases in composite appetite, hunger, food craving, and increase in fullness; (b) as hypothesised, greater decreases in both *desired* (VPCT at 2h), and *actual* (*ad libitum* meal at 3h) food intake; (c) as hypothesised, proportionally greater post-prandial decreases in plasma AG and AG/LEAP2 ratio, and greater post-prandial increases in blood glucose and insulin, and LEAP2, though the latter was a weak correlation; with (d) similar findings whether pre-load and desired/actual food intake were expressed as actual or relative calorie intake; (e) with at best small effect sizes, apart from moderate effect sizes for pre-load meal size against insulin, AG and appetite ratings.
ii. for correlations of appetite with blood measures, (a) as hypothesised, greater post-prandial increases in plasma glucose and serum insulin, and decreases in plasma AG and AG/LEAP2 ratio, were correlated with greater post-prandial decreases in composite appetite, hunger and food craving; though (b) correlations of post-prandial increases in fullness were only seen for insulin; while (c) contrary to that hypothesised, post-prandial correlations of plasma LEAP2 did not correlate with post-prandial changes in appetite measures; (d) correlations were strongest with post-prandial increase in serum insulin, followed by post-prandial decreases in plasma AG and AG/LEAP2 ratio; (e) with overall small effect sizes.
iii. for *desired* food intake (VPCT) at 2h after the pre-load, there were (a) as expected, positive correlations with composite appetite, hunger, and food craving, and negative correlations with fullness; (b) as hypothesised, negative correlations with serum insulin, and positive correlations with plasma AG and AG/LEAP2 ratio, but (c) contrary to hypotheses, no significant correlations with plasma glucose or LEAP2; with (d) strongest correlations seen with pre-load meal size, followed by composite appetite, hunger, food craving and plasma AG; with (e) similar findings when desired food intake was expressed as actual or relative calorie intake, and (e) overall small effect sizes.
iv. similarly for *actual* food intake (*ad libitum* meal) at 3h after the pre-load, there were (a) as expected, positive correlations with composite appetite, hunger, and food craving, and negative correlations with fullness; (b) as hypothesised, negative correlations with serum insulin, and positive correlations with plasma AG/LEAP2 ratio with a trend for AG; but (c) contrary to hypotheses, no significant correlations with plasma glucose or LEAP; with (d) strongest correlations seen with pre-load meal size, followed by composite appetite, hunger, and food craving, followed by fullness, and then plasma AG/LEAP2 ratio, followed by serum insulin and AG; with (e) similar findings when actual food intake was expressed as actual or relative calorie intake, and (e) overall small effect sizes..
v. similar results were seen throughout for both actual (absolute kcal) or relative (kcal %REE) food intake.
vi. in exploratory analyses of alternative post-prandial measures for appetite ratings and bloods, absolute values at 2-3h, absolute AUC, and minimum/maximum values over 2-3h usually gave stronger correlations with pre-load meal size, desired and actual food intake than the pre-registered iAUC value.

As a manipulation check, the relationships between greater pre-load meal size and proportional changes in post-prandial appetite ratings in adults without obesity were as expected and consistent with previous literature of fullness in adults without/with obesity, and hunger, desire to eat, prospective food consumption in adults without obesity (Blom et al., 2005; le Roux et al., 2005), and in this analysis are expanded to ratings of food craving. Similarly, the negative correlations of pre-load meal size with both desired and actual food intake are as expected from previous pre-load meal studies and subsequent *ad libitum* meals from several different studies, though there is usually a lack of energy compensation, i.e. overconsumption when taking into account calories consumed at the preload (Rolls et al., 2025). Indeed, in the correlation of pre-load meal size with actual food intake, the slopes were only -0.243, indicating inadequate energy compensation, since for every 100 kcal eaten at the pre-load, the actual food intake three hours later only fell on average by 24.3 kcal.

The finding that pre-load meal size lowered *desired* food intake, and that *desired* intake correlated with *actual* intake, in adults without obesity, adds to the within-participant validity and physiological utility of the VPCT. Previous studies using the VPCT (Brunstrom & Rogers, 2009) have also confirmed that *desired* intake: (i) did not change over time in adults with obesity, and *desired* intake at repeated visits was correlated in adults with normal weight (Hamm et al., 2020), (ii) decreased after gastric bypass and sleeve gastrectomy surgery for obesity that correlated with weight loss (Hamm et al., 2020), (iii) in adults without obesity, under recall or actual stress correlated with *actual* intake of similar foods under stress (Hamm et al., 2021), (iv) in adults with normal weight was lower when they considered eating for health or pleasure (Hege et al., 2018; Veit et al., 2020), and (v) was higher in adults with overweight/obesity than normal wight (Veit et al., 2020). The use of computer-based VPCT to measure *desired* food intake has advantages over *ad libitum* meals measuring *actual* intake, as the VPCT can more easily present many different foods (improving examination of interaction with different categories of food e.g. sweet vs. savoury, high- vs low-fat), is cheaper and simpler to perform, and can be utilised in functional neuroimaging (Hege et al., 2018; Veit et al., 2020).

### 4.1. Glucose

The finding that post-prandial increases in plasma glucose with increasing meal size were negatively correlated with post-prandial composite appetite and food craving, with a similar trend for hunger, but not significantly with desired and actual food intake, nor positively with post-prandial fullness, suggests that post-prandial glucose changes may not be a vital mediator of post-prandial reductions in eating behaviour. Previous studies have not examined such correlations in the context of different pre-load meal sizes given orally with the same macronutrient composition in a within-participant design. However, in 10 men with normal weight and overweight, *intraduodenal* glucose infusions at different rates (1, 2 or 4 kcal/min) achieving peak plasma glucose of 7.5-9.1 mmol/L (vs. 5.5 mmol/L during saline infusion) (135-164 vs. 99 mg/dL) dose responsively reduced *ad libitum* energy intake at 120-150 min, but there was no correlation between AUC_0-120min_ or 120 min plasma glucose and actual energy intake (Pilichiewicz et al., 2007). Furthermore, a meta-analysis examined correlations of plasma glucose with appetite ratings and actual food intake across seven post-prandial studies (Flint et al., 1998; Flint et al., 2001; Verdich et al., 2001; Bakhoj et al., 2003; Raben et al ., 2003; Flint et al., 2004a; Flint et al., 2004b), comprising 136 adults without/with obesity, using either single pre-load meals between 549 and 836 kcal, or 4-10 different pre-loads varying in macronutrient content or amount of fibre with energy content between 263 and 717 kcal, over post-prandial periods between 180 and 315 minutes (Flint et al., 2007). There were no significant correlations of post-prandial iAUC plasma glucose with post-prandial iAUC satiety or hunger, or absolute *ad libitum* energy intake in all participants, or those with normal weight, overweight or males only, including age, duration of post-prandial period and energy content of pre-load meal as covariates (Flint et al., 2007). Furthermore, in 28 men with normal weight, postprandial iAUC_0-3h_ plasma glucose across 10 different 270-715 kcal breakfast meals containing 50g carbohydrate, varying in glycaemic index and other macronutrients, did not correlate with iAUC_0-3h_ ratings of hunger, prospective food consumption, fullness nor satiety, and in fact *positively* correlated with *ad libitum* energy intake at a savoury meal at 3 hours (Flint et al., 2006).

However, negative correlations of post-prandial glucose and eating behaviour have been seen in a few studies. Although in 40 adults with type 1 diabetes mellitus, post-prandial fullness (30 mins after ∼270 kcal meal) was *positively* correlated with plasma glucose at 30 mins (with no significant negative correlation with hunger), this was only with supraphysiological glucose concentrations with majority 10-30 mmol/L (180-540 mg/dL) (Jones et al., 1997). In 24 adults with normal weight, post-prandial AUC_0-1h_ plasma glucose across five 300 kcal meals differing in type of carbohydrate, was *negatively* correlated with AUC_0-1h_ composite appetite rating, and *ad libitum* energy intake at savoury meal at 1 hour (Anderson et al., 2002). In 38 adults without obesity, post-prandial plasma glucose over 2 hours post-prandially after an *ad libitum* mixed lunch eaten over 0.5 hour (given 4 hours after a 72 kcal fixed breakfast), was *positively* correlated with fullness ratings over 2 hours (Lemmens et al., 2011).

Interestingly, later post-prandial decreases in plasma glucose after initial increases have been linked to meal initiation, suggesting that *dynamic* changes in plasma glucose may be more important than absolute concentrations. In 18 adults with normal weight 2-6 hours after a fixed meal, increases in hunger ratings and meal requests, were both preceded by and correlated with transient reductions in blood glucose (Campfield et al., 1996). In 10 men with normal weight without dietary restraint, after a 239 kcal high-carbohydrate preload, dynamic and transient declines in blood glucose correlated with meal requests, and the duration of post-prandial blood glucose response until returning to baseline was positively correlated with duration of between-meal interval and satiety ratings (Melanson et al., 1999). In a large study of 1,070 adults with normal weight, overweight and obesity, using continuous glucose monitoring at home, post-prandial glucose dips 2-3h after 5-6 meals at different times of day (fixed preload 890 kcal meals, 300 kcal oral glucose tolerance test, 500-534 kcal muffins and milkshake) differing in energy content and macronutrient composition, were a better predictor of post-prandial changes in hunger ratings at 2-3 hours and subsequent energy intake at 3-4 hours and over the subsequent 24h, than rise in plasma glucose from 0-2 hours and plasma glucose iAUC_0-2h_ (Wyatt et al., 2021).

Similarly, in association studies, using different non-oral methods of increasing plasma glucose, there are often no changes in eating behaviour. For example, in eight men with normal weight, intravenous glucose infusion rates to achieve plasma glucose of 8 mmol/L compared to 5 mmol/L (144 vs. 90 mg/dL) did not differ in ratings of hunger or desire to eat or prospective food consumption, though they did have greater fullness (Andrews et al., 1998). In adults without obesity, there were no changes in ratings of hunger, prospective food consumption, composite appetite, fullness or satiety after oral or intravenous glucose administration despite post-prandial glucose reaching ∼9 mmol/L (∼162 mg/dL) (Lauritsen et al., 2023). In 12-14 adults with normal weight, intravenous glucose infusion (vs. saline) achieving mean plasma glucose of 9.3 mmol/L mmol/L (vs. 5.3 mmol/L, 168 vs. 95 mg/dL), did not alter hunger or fullness ratings, or duration of eating, but did lower energy intake at an *ad libitum* meal by ∼15% (Chapman et al., 1998). In 15 men with normal weight, intravenous infusions of glucose (vs. saline) combined with a 218 kcal snack, achieving peak plasma glucose 9.7 mmol/L (vs. 4.3 mmol/L, 175 vs. 77 mg/dL) did not differ in ratings of hunger, appetite, fullness, or satiety (Schultes et al., 2016). In seven men with normal weight, *intraduodenal*, but not *intravenous*, infusion of glucose delivering 288 vs. 152 kcal supressed hunger, increased fullness and satiety ratings, and energy intake at *ad libitum* meal was lower with intraduodenal than Intravenous glucose infusion, despite achieving similar blood glucose concentrations of ∼9-9.5 mmol/L (∼162-171 mg/dL) prior to the meal at 2 hours, implicating other mediators than glucose related to the route of administration i.e. the incretin effect (Lavin et al., 1996).

This may be different with supraphysiological plasma glucose concentrations, for example, in six adults with normal weight, hyperglycaemic clamps without insulin, achieving plasma glucose 15 mmol/L (270 mg/dL), reduced ratings of hunger and prospective food intake, with a similar trend for wish to eat, but neither a change in fullness, nor desired food intake from a variety of food pictures, compared to control saline infusion with plasma glucose 5 mmol/L (90 mg/dL) (Gielkens et al., 1998). Interestingly infusion of 2-deoxy-D-glucose competitively inhibiting intracellular glucose utilisation did reduce hunger in five men with normal weight (Thompson & Campbell, 1977), potentially supporting a physiological role.

Thus overall, glucose appears to more consistently influence eating behaviour primarily under extreme conditions such as hypoglycaemia or hyperglycaemia, or when there are dynamic declines in plasma glucose in the later post-prandial period (Horner et al., 2020), although the latter was not seen in the current study.

### 4.2. Insulin

The present finding that larger meal size-induced increases in post-prandial insulin were associated with lower composite appetite, hunger, food craving, and desired food intake is consistent with previous literature and supports a role for insulin as a physiological signal contributing to post-prandial satiety. In the meta-analysis of seven studies (Flint et al., 1998; Flint et al., 2001; Verdich et al., 2001; Bakhoj et al., 2003; Raben et al., 2003; Flint et al., 2004a; Flint et al., 2004b), comprising 136 adults without/with overweight/obesity mentioned above (Flint et al., 2007), post-prandial iAUC plasma insulin was positively correlated with post-prandial increases in satiety in normal weight but not overweight/obesity, positively correlated with post-prandial decreases in hunger in normal weight and overweight/obesity, and negative correlations with absolute *ad libitum* energy intake in overweight/obesity but not normal weight, including age, duration of post-prandial period and energy content of pre-load meal as covariates (Flint et al., 2007). Across 41 adults without obesity, with each group given one of six different food categories as 240 kcal pre-loads, AUC_0-2h_ serum insulin was negatively correlated with *ad libitum* food intake at 2 hour, but not satiety ratings (Holt et al., 1996). Similarly, in 10 men with normal weight and overweight, *intraduodenal* glucose infusions at different rates (1, 2 or 4 kcal/min) achieving peak plasma glucose of 7.5-9.1 mmol/L (vs. 5.5 mmol/L during saline infusion, 135-164 vs. 99 mg/dL), dose responsively increased plasma insulin and reduced *ad libitum* energy intake at 2-2.5 hours, that was negatively correlated with AUC_0-2h_ and 2 hour plasma insulin (Pilichiewicz et al., 2007). However, in 15 men with normal weight, intravenous infusions of glucose (vs. saline) combined with a 218 kcal snack, achieving peak insulin of 61.7 mU/L (vs. 18.2 mU/L, 428 vs. 126 pmol/L) did not differ in ratings of hunger, appetite, fullness nor satiety (Schultes et al., 2016).

Interestingly, after low-fat 432 kcal and high-fat 1188 kcal pre-loads, there was a negative correlation between pre-meal serum insulin with subsequent *ad libitum* energy intake 5.5h later in six men with normal weight, but not in six men with obesity (Speechly & Buffenstein, 2000), while after a 598 kcal pre-load, a negative correlation was found between 3 hour, iAUC_0-3h_ and AUC_0-3h_ serum insulin and energy intake at an *ad libitum* meal at 3.2 hours in 12 men with normal weight, but not in 19 men with obesity (Verdich et al., 2001). This suggests that obesity-associated insulin resistance might attenuate the satiety effect of post-prandial insulin. Indeed, neuroimaging studies have found that peripheral and brain insulin resistance are related to each other using systemic and intranasal insulin administration (Heni et al., 2015; Kullmann et al., 2020).

In seven men with normal weight, *intraduodenal*, but not *intravenous*, infusion of glucose delivering 288 vs. 152 kcal supressed hunger, increased fullness and satiety ratings, and energy intake at *ad libitum* meal was lower with intraduodenal than intravenous glucose infusion associated with greater plasma insulin prior to the meal at 2 hours (Lavin et al., 1996). Furthermore, administration of the somatostatin analogue octreotide lowered plasma insulin and blocked the attenuation of eating behaviour by intraduodenal glucose, suggesting a satiety effect of insulin, though other hormonal incretin mediators may be involved such as GLP-1 since they will also be suppressed by octreotide (Lavin et al., 1996).

However, evidence for satiating effects of insulin have not been seen in hyperinsulinaemic clamps studies in adults with normal weight. In 20 adults with normal weight, hyperinsulinaemic infusions (achieving plasma insulin 100-150 mU/L compared to 20 mU/L during saline infusion, 694-1042 vs. 139 pmol/L) in fact *increased* hunger ratings under both hypoglycaemic (3.3 mmol/L, 59 mg/dL) and mildly hyperglycaemic (6.9 mmol/L, 124 mg/dL) conditions, and enhanced palatability of sweet solutions (Rodin et al., 1985). In eight adults with normal weight, a hyperinsulinaemic euglycaemic clamp achieving a 3-4-fold increase in serum insulin and maintaining plasma glucose at ∼5.0 mmol/L (compared to ∼4.2 mmol/L during saline infusion, ∼90 vs. ∼76 mg/dL), had no effect on an *ad libitum* intake of a mixed liquid meal (Woo et al., 1984). In 14 adults with normal weight, low- and high-dose hyperinsulinaemic euglycaemic clamps (achieving plasma insulin 47-95 mU/L vs. 6 mU/L at saline infusion (326-660 vs. 42 pmol/L, maintaining plasma glucose at ∼5 mmol/L, ∼90 mg/dL) had no effect on hunger and fullness ratings, nor *ad libitum* food intake at a mixed meal (Chapman et al.). In six adults with normal weight, hyperinsulinaemic euglycaemic clamps (achieving plasma insulin 85-105 mU/L, compared to 5-7 mU/L with saline infusion, 590-729 vs. 35-49 pmol/L, both with plasma glucose 4-5 mmol/L, 72-90 mg/dL), had no effects on ratings of hunger, wish to eat, prospective food intake, fullness, nor desired food intake from a variety of food pictures (Gielkens et al., 1998). In two groups of adults without obesity (n=16-18, matched for BMI and age), receiving either a hyperinsulinaemic euglycaemic clamp or saline (achieving plasma insulin ∼151-168 vs. ∼11-13 mU/L, ∼1049-1167 vs. ∼76-90 pmol/L, with plasma glucose 4.9-5.2 mmol/L, 88-94 mg/dL), there was no between group difference in hunger ratings, nor wanting or liking ratings or simultaneous brain responses using functional MRI to high-calorie or low-calorie foods (Belfort-DeAguiar et al., 2016).

Mechanistically, insulin receptors are widely expressed within hypothalamic and brainstem nuclei involved in energy homeostasis and within mesolimbic reward pathways, where insulin signalling has been proposed to reduce food motivation and enhance satiety via actions of mesolimbic dopamine circuits (Mebel et al., 2012; Labouebe et al., 2013; Heni et al., 2015; Tiedemann et al., 2017; Kullmann et al., 2020; Mitchell & Begg, 2021).

Although in the present study, there was a significant negative correlation between postprandial increases in serum insulin and *desired* food intake, this was not significant for *actual* intake, although there was a similar trend. This discrepancy may reflect the smaller sample size available for analyses of actual food intake, reducing statistical power, as well as methodological differences between measures, such as the limited range of foods offered at the *ad libitum* meal relative to VPCT (4 vs. 16 options), which participants may have found less appealing.

### 4.3. LEAP2

The finding that post-prandial increases in LEAP2 were correlated with post-prandial increases in preload meal size up to 1200 kcal in a within participant design in adults without obesity, confirm the previous findings from analysis across different studies without and with obesity, with pre-load meals of between 337-730 kcal, despite their different macronutrient composition, and time points of blood collection (Emini et al., 2024). However, the current study only found a weak correlation between pre-load meal size and post-prandial increases in plasma LEAP2, though the effects of larger pre-loads above 1200 kcal remain unknown, as does the effects in those with obesity, where post-prandial increases in plasma LEAP2 are greater, especially in women (Mani et al., 2019; Andreoli et al ., 2024a).

The lack of any significant correlations of post-prandial plasma LEAP2 after differing meal size in adults without obesity with hunger, fullness, food craving and composite appetite ratings, nor desired or actual food intake, was contrary to our hypothesis and not in agreement with the few previous studies using single meal sizes. In 22 adults without obesity, post-prandial increases in plasma LEAP2 correlated positively with post-prandial decreases in composite appetite (averaged across 70 and 150 mins, vs. another visit where they remained fasted), and post-prandial decreases in cue reactivity to high-energy/low-energy or high-energy food using functional MRI (fMRI, at 70 mins), with a similar trend for *ad libitum* food intake (at 150 minutes), though not with post-prandial increases in fullness, nor post-prandial decreases in appeal rating of high-energy or high-energy vs. low-energy foods, though this was only examined after a single 730 kcal meal (Bhargava et al ., 2023). However, in adults with obesity with poorly controlled T2DM, receiving a low-calorie diet over 12 months without (n=9-13) or with (n=11-23) insertion of a duodenal-jejunal bypass liner device, there were no correlations across multiple visits of post-prandial iAUC_0-2h_ LEAP2 and post-prandial iAUC_0-2h_ of composite appetite nor fullness (after 600 kcal pre-load) (Emini et al., 2024). Similarly, in 44 adults with normal weight/overweight/obesity, post-prandial plasma LEAP2 at 1 hour after 400-600 kcal pre-load (20% daily energy requirement) did not correlate with ratings of hunger, desire to eat, prospective food consumption, nor fullness (adjusting for BMI and sex) (Andreoli et al., 2024a). Furthermore, in 123 adults with normal-weight, overweight and obesity, across fasting and post-prandial time points (120 mins after a high protein (35% total kcal) *ad libitum* meal), plasma LEAP2 did not correlate with hunger, desired food intake, fullness, explicit liking or wanting of low-fat savoury, high-fat savoury, low-fat sweet or high-fat sweet foods (including sex, age, and BMI category as covariates), though surprisingly plasma LEAP2 fell after the *ad libitum* meal, which might be due to the high protein content (Andreoli et al., 2024b).

These discrepancies may be related to single versus multiple meal sizes, variations in post-prandial periods and sample sizes, BMI category and presence of T2DM. However, experimental infusion studies do provide evidence that LEAP2 suppresses eating behaviour. In both men without and with obesity, intravenous LEAP2 infusions, producing supraphysiological plasma LEAP2 concentrations, reduced *ad libitum* food intake at a savoury meal by ∼12%, though without any significant changes in hunger, prospective food consumption, fullness or satiety ratings (Hagemann et al., 2022; Englund et al., 2026). These findings do support a stimulatory role for GHSR signalling on human eating behaviour, but evidence for the physiological increases in plasma LEAP2 after food intake attenuating human eating behaviour needs further confirmation.

### 4.4. Ghrelin and ghrelin/LEAP2 ratio

The present study demonstrated that larger preload meal sizes were associated with greater post-prandial reductions in plasma AG concentrations and the AG/LEAP2 molar ratio, indicating a progressive attenuation of ghrelin receptor signalling with increasing energy intake. This finding extends previous observations that post-prandial ghrelin suppression is dependent on meal energy content. In adults without obesity, increasing pre-load meals size between 250-3,000 kcal led to proportional reductions in plasma total ghrelin AUC_0-3h_ despite slight variations in macronutrient content (le Roux et al., 2005), while in men, there was greater suppression of plasma AG over 4 hours after consuming an 800 kcal than 200–400 kcal meal of identical macronutrient composition (Ishii et al., 2016).

Analysis of correlations between plasma ghrelin and appetite ratings are complicated by large variability between study designs including sample, size, sex, BMI status of participants, whether measuring total, acyl or desacyl ghrelin, use of protease inhibitors and acidification of plasma to prevent degradation of AG to DAG (Liu et al., 2008; Blatnik & Soderstrom, 2011), nutritional state (fasted, fed or pre-meal), pre-load meal size and macronutrient content, whether analysing post-prandial changes in ghrelin and ratings in terms of absolute concentrations, changes from baseline or absolute or incremental AUC, post-prandial time points, analysis of single of multiple meals, and whether studied after a weight loss intervention such as low-calorie or very low-calorie diet.

When restricting analysis to correlations with plasma AG when fed (as in the current study), a few studies have found *positive* correlations of post-prandial plasma AG with post-prandial hunger in adults with normal weight/overweight/obesity (Mackelvie et al ., 2007), overweight/obesity (Gibbons et al., 2013) and desire to eat in normal weight/overweight/obesity (Mackelvie et al., 2007), normal weight (Martens et al., 2012), and obesity (Andarini et al., 2017). However, most studies have found no significant correlations of post-prandial AG with composite appetite in normal weight/overweight (Deighton et al., 2014; Bhargava et al., 2023); hunger in normal weight (Andarini et al., 2017), overweight (Unick et al., 2010), normal weight/overweight/obesity (Liu et al., 2015; Andreoli et al., 2024a), and obesity (Andarini et al., 2017; Nymo et al., 2018); desire to eat in normal weight (Andarini et al., 2017), normal weight/overweight/obesity (Andreoli et al., 2024a), and obesity (Nymo et al., 2018); prospective food consumption in normal weight/overweight/obesity (Mackelvie et al., 2007; Liu et al., 2015; Andreoli et al., 2024a), and obesity (Nymo et al., 2018); fullness in normal weight/overweight (Bhargava et al., 2023), normal weight/overweight/obesity (Mackelvie et al., 2007; Liu et al., 2015; Andreoli et al., 2024a), overweight/obesity (Gibbons et al., 2013), and obesity (Nymo et al., 2018); nor satiety in normal weight/overweight/obesity (Liu et al., 2015).

Furthermore, only one study has found positive corelation of post-prandial AG with *ad libitum* energy intake of savoury/sweet foods in adults with overweight/obesity (Gibbons et al., 2013), with no correlation found with *ad libitum* energy intake in adults with normal weight (Roth et al., 2022), overweight (Unick et al., 2010; St-Onge et al., 2014), normal weight/overweight (even when adjusting for estimated resting energy expenditure) (Bhargava et al., 2023), nor obesity (Roth et al., 2022).

However, none of the above studies examining correlations of post-prandial AG with appetite ratings or *ad libitum* food intake have used pre-loads of increasing caloric size with identical macronutrient composition as in the current study. The previous studies demonstrating suppressive effects of preload meal size on plasma total ghrelin and AG did not examine appetite ratings nor *ad libitum* energy intake (le Roux et al., 2005; Ishii et al., 2016).

Importantly, the AG/LEAP2 ratio may provide a more physiologically relevant index of ghrelin system activity than AG concentrations alone. LEAP2 acts as both an endogenous antagonist (with similar potency to AG but present at higher concentrations in plasma) and inverse agonist of the constitutively active GHSR, such that the balance between circulating AG and LEAP2 determines the net orexigenic drive mediated through ghrelin receptor signalling. Recent evidence suggests that the AG/LEAP2 ratio responds dynamically to nutritional status, increasing during fasting and energy restriction and decreasing following nutrient ingestion, reflecting coordinated regulation of ghrelin receptor activation by both ligands (Ge et al., 2018; Mani et al., 2019). The present finding that larger meals produced greater reductions in the AG/LEAP2 ratio therefore suggests that meal size influences not only circulating ghrelin concentrations but also the overall functional activity of the ghrelin signalling system.

Indeed, in the present study plasma AG/LEAP2 ratio was negatively associated with subjective appetite ratings, desired and actual food intake measures, with slightly stronger correlations for actual intake than plasma AG alone. Supportive findings have been seen in other post-prandial studies. In 22 adults without obesity, post-prandial decreases in plasma AG/LEAP2 tended to correlate with post-prandial increases in fullness (averaged across 70 and 150 mins, vs. visit where remained fasted), and post-prandial decreases in appeal rating of high-energy vs. low energy foods (at 70 mins), but did not correlate with post-prandial decreases in composite appetite, cue reactivity to high-energy/low-energy or high-energy food using fMRI (at 70 mins), nor *ad libitum* food intake (at 150 mins), though this was only examined after a single 730 kcal meal (Bhargava et al., 2023). In 44 adults with normal weight/overweight/obesity, post-prandial plasma AG/LEAP2 at 1 hour after a 400-600 kcal pre-load (20% daily energy requirement) did not correlate with ratings of hunger, desire to eat, prospective food consumption, nor fullness (adjusting for BMI and sex) (Andreoli et al., 2024a). In 10 men with normal weight men, across 3 visits at which they consumed oral ketone, isocaloric isovolumetric glucose (259 kcal) or control 0 kcal drinks, total ghrelin/LEAP2 ratio, across baseline, 60, 180 and 300 mins post-prandial time points was positively correlated with hunger and prospective food consumption, and negatively correlated with satiety and fullness, however the analysis was not performed using a repeated measures model (Holm et al., 2023).

Interestingly, in the current study, correlations between plasma AG and AG/LEAP2 ratio were only significant for hunger, composite appetite and food craving, but not fullness (which was only correlated positively with serum insulin). This could be because of inter-individual differences in interpretation of the interoceptive nature of feeling ‘full’, which might be experienced as the opposite of hunger, discomfort, bloating or satisfaction (satiety) etc. (Cheon et al., 2019), or because ‘fullness’ is also mediated via non-blood mediators such as vagus nerve signalling from gastric stretch receptors. However, negative correlations of fullness with plasma AG and AG/LEAP2 ratio did reach significant small effect sizes when using absolute values at 2h, absolute AUC _0-3h_ and minimum value over 3h, instead of iAUC _0-3h_, which may reflect variability in baseline ratings (see Section 4.5).

### 4.5. Exploratory post-prandial end points

Interestingly, in exploratory analyses of different post-prandial measures for appetite ratings and bloods, absolute values at 2-3h, absolute AUC, and minimum/maximum values over 2-3h gave stronger correlations with pre-load meal size, desired and actual food intake than iAUC value (**Supplementary Results** and **Supplementary Figures S18-S21**). This may be because the latter is susceptible to variation in baseline values, as has previously been suggested for interpretation of continuous glucose monitoring data (Thomas et al ., 2026), although there was no systematic variation in baseline fasting values for any measurement in the current study by subsequent pre-load meal size. The appetite literature is indeed highly variable as to which exact post-prandial measures are assessed, but these results suggest that absolute AUC or absolute values at relevant time points may be preferable to iAUC in future correlational studies. Indeed, correlations of desired food intake at VPCT with ratings of fullness, hunger, composite appetite and food craving did reached moderate effect size for absolute values at 2h and absolute AUC_0-2h_ (**Supplementary Figure S19**), as did correlations of plasma AG and AG/LEAP2 ratio with hunger, composite appetite and food craving for absolute values at 2-3h and absolute AUC_0-3h_ (**Supplementary Figure S21**).

### 4.6. Overall strengths and limitations

This study used a single-blinded, crossover, within-participant design, using standardised volume of macronutrient content and flavour of liquid pre-loads, and measuring energy intake by different means of both absolute kcal and relative to estimated %REE, and both desired and actual food intake, using a variety of foods, especially for the former.

Although there was a good overall number of visits, the participant sample size was limited, with loss of participants with low desired or actual food intake at 0 kcal visit, especially for the latter, with multiple correlations between related variables, leading to risk of type I and II errors. The pre-load meal sizes were not blinded to the investigators. Furthermore, this study was only in participants without obesity.

The correlational nature of the statistical analysis does not allow determination of causality. Correlational effect sizes of both appetite ratings and blood measures with desired and actual food intake were at best small, giving limited individual validity as predictors of next-meal intake, but also reflecting the numerous potential mediators of satiety. Future analyses should include multivariate approaches. Incremental AUC values were used for correlations of appetite and blood measures with food intake to provide consistency with the statistical analyses of the post-prandial impact of pre-load meals sizes on eating behaviour, and avoid problems of multiple comparisons when using different post-prandial time points. However, as revealed in the exploratory analyses, individual time points or absolute AUC may give stronger correlations than iAUC values, which did enable more correlations to reach moderate effect size. It i s unclear what are the time delays between blood measurements and hormonal actions in the brain on human eating behaviour, and so consideration should be made for analyses of multiple and even staggered time points in future correlational analyses. Surprisingly though, in a prior study of adults without/with obesity, correlations of plasma total ghrelin with hunger were strongest when hunger measurement was 0-30 min *behind* the time of ghrelin measurement (rather than in advance) (Frecka & Mattes, 2008).

### 4.7. Future work

Ongoing analyses from the study are examining correlations with other potential post-prandial plasma hormone mediators of satiety including PYY, GLP-1, PP and GIP; impacts of pre-load meals size, appetite and blood measures on desired and actual food intake classified by savoury/sweet and low/high fat food categories, as well as hedonic taste ratings at the *ad libitum* meal, to see if there is any preferential impact on food type. Direct causative relationships between variables will also be investigated using repeated measures Bayesian mediation analysis to determine the role of blood measures in mediating influence of preload meal size on eating behaviour, as well as consideration of multivariate correlational approaches, though collinearity of potential mediators may be an issue

### 4.8. Conclusion

In conclusion, in adults without obesity, the greater the pre-load meal size the greater the post-prandial increase in plasma glucose and serum insulin, and the greater fall in plasma AG and AG/LEAP2 ratio, which was associated with a greater attenuation of eating behaviour, as assessed by appetite ratings and desired and actual food intake. The strongest relationships with blood measures were seen for serum insulin, plasma AG and AG/LEAP2 ratio. However, as correlations of food intake with preload meal size and appetite ratings appeared stronger than with hormone measures this suggests that additional factors, such as vagal innervation, nutrients and interaction with other appetitive hormones will be contributing to the overall satiety cascade. Although there was a weak effect of increasing pre-load meal size to proportionately increase plasma LEAP2, this was not related to post-prandial attenuation of eating behaviour, and this warrants further study in obesity.

## Data Availability

All data produced in the present study are available upon reasonable request to the authors

## ACKNOWLEDGEMENTS

This work was supported by a UK Research Institute (UKRI) Medical Research Council (MRC) Population and Systems Medicine Board (PSMB) project grant (MR/T017279/1) and RIPEN Innovation Hub Progression Award, Biotechnology and Biological Sciences Research Council (BBSRC) Diet and Health Open Innovation Research Club grant (PA6449) to APG, and Indonesia Endowment Fund for Education (LPDP PTUD) Scholarship to TP. Infrastructure support was provided by the NIHR Imperial Biomedical Research Centre and the NIHR Imperial Clinical Research Facility, London. The views expressed are those of the authors and not necessarily those of the National Health Service (NHS), the NIHR, or the Department of Health and Social Care (DHSC). We thank staff at the NIHR Imperial Clinical Research Facility, Imperial College Healthcare NHS Trust, Hammersmith Hospital, London, UK; Departments of Clinical Biochemistry at Imperial College Healthcare NHS Trust, London, for performing glucose and insulin assays; Phoenix Pharmaceuticals Inc., CA, USA, and Synnovis Ltd, Viapath, Kings College Hospital, London, UK, for performing plasma LEAP2 and AG assays; and the participants for taking part in the study.

For the purpose of open access, the authors have applied a Creative Commons Attribution (CC-BY) licence to any author accepted manuscript version arising.

## DISCLOSURES

APG is a consultant for Rhythm Pharmaceuticals; has been a clinical investigator for clinical trials sponsored by Rhythm Pharmaceuticals, Millendo Therapeutics and Soleno Therapeutics; has been a consultant for Eli Lilly, Evidera, Helsinn Healthcare S.A, Idera Pharmaceuticals, Novo Nordisk, Radius Health, Soleno Therapeutics, Tonix Pharmaceuticals and Vida Ventures; has been on Advisory Boards for Radius Health and Millendo Therapeutics; has been on Data Safety Monitoring Committee for Novo Nordisk; has received speaker fees from Eli Lilly, Novo Nordisk, Soleno Therapeutics and Rhythm Pharmaceuticals, and research support from Novo Nordisk. None of the other authors have any relevant disclosures.

## CONTRIBUTIONS

Conceptualisation: APG; Data curation: APG; Formal analysis: TP, JL, RB, WIN, JG, MRF, XZ, ME, WL, SL, APG; Funding acquisition: APG; Investigation: TP, JL, RB, WIN, JG, MRF, XZ, ME, WL, SL, ZB, APG; Methodology: RB, JB, APG; Project administration: RB, MRF, APG; Resources: JB, APG; Software: JB, APG; Supervision: APG; Visualisation: TP, JL, APG; Writing - original draft preparation: TP, JL, APG; Writing - review & editing: all authors.

## Abbreviations

AG: acyl ghrelin
AUC: area under curve
BIA: bioimpedance analysis
BMI: body mass index
DAG: desacyl ghrelin
GHSR: GH secretagogue receptor
GOAT: ghrelin-O-acyl transferase
h: hours
HE: high-energy
iAUC: incremental AUC
LE: low-energy
LEAP2: liver-expressed antimicrobial peptide-2
REE: resting energy expenditure
SEM: standard error of mean
VAS: visual analogue scale
VPCT: virtual portion size creation task.

## Notes

### Clinical Trial

NCT06013592

### Author Declarations

Ethics Committee of West London & GTAC gave ethical approval for this work

### Summary of Updates

Reference to sex of participant symbols in correlational graphs and description of demographics. Inclusion of sex as factor in analysis of relationship between preload meal size with desired and actual food intake. Correction of some errors in r, r2 and beta-values throughout Figures in Main and Supplemental Results. Mention of effect sizes when reporting and discussing results. Inclusion of FDR correction for correlation summary heat map results. Inclusion of additional limitations to Discussion. Addition/correction of some references in Introduction.

## REFERENCES

Andarini, S., Kangsaputra, F. B., & Handayani, D. (2017). Pre- and postprandial acylated ghrelin in obese and normal weight men. Asia Pac J Clin Nutr. 26(Suppl 1), S85–S91. 10.6133/apjcn.062017.s5

Anderson, G. H., Catherine, N. L., Woodend, D. M., & Wolever, T. M. (2002). Inverse association between the effect of carbohydrates on blood glucose and subsequent short-term food intake in young men. Am J Clin Nutr. 76(5), 1023–30. 10.1093/ajcn/76.5.1023

Anderson, K. C., Hasan, F., Grammer, E. E., & Kranz, S. (2023). Endogenous Ghrelin Levels and Perception of Hunger: A Systematic Review and Meta-Analysis Adv Nutr. 14(5), 1226–1236.

Andreoli, M. F., Fittipaldi, A. S., Castrogiovanni, D., De Francesco, P. N., Valdivia, S., Heredia, F., Ribet-Travers, C., Mendez, I., Fasano, M. V., Schioth, H. B., Doi, S. A., Habib, A. M., & Perello, M. (2024a). Pre-prandial plasma liver-expressed antimicrobial peptide 2 (LEAP2) concentration in humans is inversely associated with hunger sensation in a ghrelin independent manner. Eur J Nutr. 63(3), 751–762. 10.1007/s00394-023-03304-8

Andreoli, M. F., Kruger, A. L., Sokolov, A. V., Rukh, G., De Francesco, P. N., Perello, M., & Schioth, H. B. (2024b). LEAP2 is associated with impulsivity and reward sensitivity depending on the nutritional status and decreases with protein intake in humans. Diabetes Obes Metab. 26(10), 4734–4743. 10.1111/dom.15850

Andrews, J. M., Rayner, C. K., Doran, S., Hebbard, G. S., & Horowitz, M. (1998). Physiological changes in blood glucose affect appetite and pyloric motility during intraduodenal lipid infusion. Am J Physiol. 275(4), G797–804. 10.1152/ajpgi.1998.275.4.G797

Asmar, M., Tangaa, W., Madsbad, S., Hare, K., Astrup, A., Flint, A., Bulow, J., & Holst, J. J. (2010). On the role of glucose-dependent insulintropic polypeptide in postprandial metabolism in humans. Am J Physiol Endocrinol Metab. 298(3), E614–21. 10.1152/ajpendo.00639.2009

Bakhoj, S., Flint, A., Holst, J. J., & Tetens, I. (2003). Lower glucose-dependent insulinotropic polypeptide (GIP) response but similar glucagon-like peptide 1 (GLP-1), glycaemic, and insulinaemic response to ancient wheat compared to modern wheat depends on processing. Eur J Clin Nutr. 57(10), 1254–61. 10.1038/sj.ejcn.1601680

Belfort-DeAguiar, R., Seo, D., Naik, S., Hwang, J., Lacadie, C., Schmidt, C., Constable, R. T., Sinha, R., & Sherwin, R. (2016). Food image-induced brain activation is not diminished by insulin infusion. Int J Obes (Lond). 40(11), 1679–1686. 10.1038/ijo.2016.152

Bhargava, R., Luur, S., Rodriguez Flores, M., Emini, M., Prechtl, C. G., & Goldstone, A. P. (2023). Post-prandial increases in liver-gut hormone LEAP2 correlate with attenuated eating behaviour in adults without obesity. J Endocr Soc. 7(7), bvad061. 10.1210/jendso/bvad061

Blatnik, M., & Soderstrom, C. I. (2011). A practical guide for the stabilization of acyl ghrelin in human blood collections. Clin Endocrinol (Oxf). 74(3), 325–31. 10.1111/j.1365-2265.2010.03916.x

Blom, W. A., Stafleu, A., de Graaf, C., Kok, F. J., Schaafsma, G., & Hendriks, H. F. (2005). Ghrelin response to carbohydrate-enriched breakfast is related to insulin. Am J Clin Nutr. 81(2), 367–75. 10.1093/ajcn.81.2.367

Blundell, J. (1991). Pharmacological approaches to appetite suppression. Trends Pharmacol Sci. 12(4), 147–57. 10.1016/0165-6147(91)90532-w

Blundell, J., de, G. C., Hulshof, T., Jebb, S., Livingstone, B., Lluch, A., Mela, D., Salah, S., Schuring, E., van der Knaap, H., & Westerterp, M. (2010). Appetite control: methodological aspects of the evaluation of foods. Obes. Rev. 11(3), 251–270.

Bohn, M. J., Krahn, D. D., & Staehler, B. A. (1995). Development and initial validation of a measure of drinking urges in abstinent alcoholics. Alcohol Clin Exp Res. 19(3), 600–6. 10.1111/j.1530-0277.1995.tb01554.x

Briggs, D. I., Lockie, S. H., Wu, Q., Lemus, M. B., Stark, R., & Andrews, Z. B. (2013). Calorie-restricted weight loss reverses high-fat diet-induced ghrelin resistance, which contributes to rebound weight gain in a ghrelin-dependent manner. Endocrinology. 154(2), 709–17. 10.1210/en.2012-1421

Brunstrom, J. M., & Rogers, P. J. (2009). How many calories are on our plate? Expected fullness, not liking, determines meal-size selection. Obesity (Silver Spring). 17(10), 1884–90. 10.1038/oby.2009.201

Cameron, J. D., Goldfield, G. S., Finlayson, G., Blundell, J. E., & Doucet, E. (2014). Fasting for 24 hours heightens reward from food and food-related cues. PLoS One. 9(1), e85970. 10.1371/journal.pone.0085970

Campfield, L. A., Smith, F. J., Rosenbaum, M., & Hirsch, J. (1996). Human eating: evidence for a physiological basis using a modified paradigm. Neurosci Biobehav Rev. 20(1), 133–7. 10.1016/0149-7634(95)00043-e

Chapman, I. M., Goble, E. A., Wittert, G. A., Morley, J. E., & Horowitz, M. (1998). Effect of intravenous glucose and euglycemic insulin infusions on short-term appetite and food intake. Am J Physiol. 274(3 Pt 2), R596–R603.

Cheon, B. K., Sim, A. Y., Lee, L., & Forde, C. G. (2019). Avoiding hunger or attaining fullness? Implicit goals of satiety guide portion selection and food intake patterns. Appetite. 138(10-16. 10.1016/j.appet.2019.03.003

Cornejo, M. P., Castrogiovanni, D., Schioth, H. B., Reynaldo, M., Marie, J., Fehrentz, J. A., & Perello, M. (2019). Growth hormone secretagogue receptor signalling affects high-fat intake independently of plasma levels of ghrelin and LEAP2, in a 4-day binge eating model. J Neuroendocrinol. 31(10), e12785. 10.1111/jne.12785

Coutinho, S. R., Halset, E. H., Gasbakk, S., Rehfeld, J. F., Kulseng, B., Truby, H., & Martins, C. (2018). Compensatory mechanisms activated with intermittent energy restriction: A randomized control trial. Clin Nutr. 37(3), 815–823. 10.1016/j.clnu.2017.04.002

de Graaf, C., Blom, W. A., Smeets, P. A., Stafleu, A., & Hendriks, H. F. (2004). Biomarkers of satiation and satiety. Am J Clin Nutr. 79(6), 946–61. 10.1093/ajcn/79.6.946

de Vries, R., Morquecho-Campos, P., de Vet, E., de Rijk, M., Postma, E., de Graaf, K., Engel, B., & Boesveldt, S. (2020). Human spatial memory implicitly prioritizes high-calorie foods. Sci Rep. 10(1), 15174. 10.1038/s41598-020-72570-x

Deighton, K., Batterham, R. L., & Stensel, D. J. (2014). Appetite and gut peptide responses to exercise and calorie restriction. The effect of modest energy deficits. Appetite. 81(52-9. 10.1016/j.appet.2014.06.003

Druce, M. R., Wren, A. M., Park, A. J., Milton, J. E., Patterson, M., Frost, G., Ghatei, M. A., Small, C., & Bloom, S. R. (2005). Ghrelin increases food intake in obese as well as lean subjects. Int J Obes. 29(9), 1130–1136.

Ebert, R., & Creutzfeldt, W. (1989). Gastric inhibitory polypeptide (GIP) hypersecretion in obesity depends on meal size and is not related to hyperinsulinemia. Acta Diabetol Lat. 26(1), 1–15. 10.1007/BF02581191

Emini, M., Bhargava, R., Aldhwayan, M., Chhina, N., Rodriguez Flores, M., Aldubaikhi, G., Al Lababidi, M., Al-Najim, W., Miras, A. D., Ruban, A., Glaysher, M. A., Prechtl, C. G., Byrne, J. P., Teare, J. P., & Goldstone, A. P. (2024). Satiety hormone LEAP2 after low-calorie diet with/without Endobarrier insertion in obesity and type 2 diabetes mellitus. J Endocr Soc. 9(1), bvae214. 10.1210/jendso/bvae214

Englund, A., Lange, A. H., Hagemann, C. A., Kizilkaya, H. S., Rosenkilde, M. M., Hartmann, B., Holst, J. J., Dela, F., Lund, A. B., Gasbjerg, L. S., & Knop, F. K. (2026). LEAP2 reduces ad libitum food intake and attenuates postprandial glucose excursions in men with obesity. Diabetes. 75(925-937. 10.2337/db25-1132

Fernandez, G., Cabral, A., De Francesco, P. N., Uriarte, M., Reynaldo, M., Castrogiovanni, D., Zubiria, G., Giovambattista, A., Cantel, S., Denoyelle, S., Fehrentz, J. A., Tolle, V., Schioth, H. B., & Perello, M. (2022). GHSR control s food deprivation-induced activation of CRF neurons of the hypothalamic paraventricular nucleus in a LEAP2-dependent manner. Cell Mol Life Sci. 79(5), 277. 10.1007/s00018-022-04302-5

Flint, A., Gregersen, N. T., Gluud, L. L., Moller, B. K., Raben, A., Tetens, I., Verdich, C., & Astrup, A. (2007). Associations between postprandial insulin and blood glucose responses, appetite sensations and energy intake in normal weight and overweight individuals: a meta-analysis of test meal studies. Br J Nutr. 98(1), 17–25. 10.1017/S000711450768297X

Flint, A., Moller, B. K., Raben, A., Pedersen, D., Tetens, I., Holst, J. J., & Astrup, A. (2004a). The use of glycaemic index tables to predict glycaemic index of composite breakfast meals. Br J Nutr. 91(6), 979–89. 10.1079/bjn20041124

Flint, A., Moller, B. K., Raben, A., Sloth, B., Pedersen, D., Tetens, I., Holst, J. J., & Astrup, A. (2006). Glycemic and insulinemic responses as determinants of appetite in humans. Am J Clin Nutr. 84(6), 1365–73. 10.1093/ajcn/84.6.1365

Flint, A., Raben, A., Astrup, A., & Holst, J. J. (1998). Glucagon-like peptide 1 promotes satiety and suppresses energy intake in humans. J Clin Invest. 101(3), 515–520.

Flint, A., Raben, A., Ersboll, A. K., Holst, J. J., & Astrup, A. (2001). The effect of physiological levels of glucagon-like peptide-1 on appetite, gastric emptying, energy and substrate metabolism in obesity. Int J Obes Relat Metab Disord. 25(6), 781–92. 10.1038/sj.ijo.0801627

Flint, A., Tangaa, W., Rugbjerg, K., Raben, A., Astrup, A., & Holst, J. J. (2004b). The effect of GIP on postprandial appetite and energy expenditure. Int J Obes. 28(Suppl 1), 168.

Flint, D. J. (1998). Effects of antibodies to adipocytes on body weight, food intake, and adipose tissue cellularity in obese rats. Biochemical and Biophysical Research Communications. 252(1), 263–268.

Frecka, J. M., & Mattes, R. D. (2008). Possible entrainment of ghrelin to habitual meal patterns in humans. Am J Physiol Gastrointest Liver Physiol. 294(3), G699–707. 10.1152/ajpgi.00448.2007

Fuhrer, D., Zysset, S., & Stumvoll, M. (2008). Brain activity in hunger and satiety: an exploratory visually stimulated fMRI study. Obesity. 16(945–950. 10.1038/oby.2008.33

Garutti, M., Sirico, M., Noto, C., Foffano, L., Hopkins, M., & Puglisi, F. (2025). Hallmarks of Appetite: A Comprehensive Review of Hunger, Appetite, Satiation, and Satiety. Curr Obes Rep. 14(1), 12. 10.1007/s13679-024-00604-w

Gasbjerg, L. S., Bari, E. J., Christensen, M., & Knop, F. K. (2021). Exendin(9-39)NH(2) : Recommendations for clinical use based on a systematic literature review. Diabetes Obes Metab. 23(11), 2419–2436. 10.1111/dom.14507

Ge, X., Yang, H., Bednarek, M. A., Galon-Tilleman, H., Chen, P., Chen, M., Lichtman, J. S., Wang, Y., Dalmas, O., Yin, Y., Tian, H., Jermutus, L., Grimsby, J., Rondinone, C. M., Konkar, A., & Kaplan, D. D. (2018). LEAP2 is an endogenous antagonist of the ghrelin receptor. Cell Metab. 27(2), 461–469 e6. 10.1016/j.cmet.2017.10.016

Gibbons, C., Caudwell, P., Finlayson, G., Webb, D. L., Hellstrom, P. M., Naslund, E., & Blundell, J. E. (2013). Comparison of postprandial profiles of ghrelin, active GLP-1, and total PYY to meals varying in fat and carbohydrate and their association with hunger and the phases of satiety. J Clin Endocrinol Metab. 98(5), E847–55. 10.1210/jc.2012-3835

Gielkens, H. A., Verkijk, M., Lam, W. F., Lamers, C. B., & Masclee, A. A. (1998). Effects of hyperglycemia and hyperinsulinemia on satiety in humans. Metabolism. 47(3), 321–4. 10.1016/s0026-0495(98)90264-5

Goldstone, A. P., Prechtl, C. G., Scholtz, S., Miras, A. D., Chhina, N., Durighel, G., Deliran, S. S., Beckmann, C., Ghatei, M. A., Ashby, D. R., Waldman, A. D., Gaylinn, B. D., Thorner, M. O., Frost, G. S., Bloom, S. R., & Bell, J. D. (2014). Ghrelin mimics fasting to enhance human hedonic, orbitofrontal cortex, and hippocampal responses to food. Am J Clin Nutr. 99(6), 1319–30. 10.3945/ajcn.113.075291

Goldstone, A. P., Prechtl de Hernandez, C. G., Beaver, J. D., Muhammed, K., Croese, C., Bell, G., Durighel, G., Hughes, E., Waldman, A. D., Frost, G., & Bell, J. D. (2009). Fasting biases brain reward systems towards high-calorie foods. Eur J Neurosci. 30(8), 1625–35. 10.1111/j.1460-9568.2009.06949.x

Hagemann, C. A., Jensen, M. S., Holm, S., Gasbjerg, L. S., Byberg, S., Skov-Jeppesen, K., Hartmann, B., Holst, J. J., Dela, F., Vilsboll, T., Christensen, M. B., Holst, B., & Knop, F. K. (2022). LEAP2 reduces postprandial glucose excursions and ad libitum food intake in healthy men. Cell Rep Med. 3(4), 100582. 10.1016/j.xcrm.2022.100582

Hamm, J. D., Dotel, J., Tamura, S., Shechter, A., Herzog, M., Brunstrom, J. M., Albu, J., Pi-Sunyer, F. X., Laferrere, B., & Kissileff, H. R. (2020). Reliability and responsiveness of virtual portion size creation tasks: Influences of context, foods, and a bariatric surgical procedure. Physiol Behav. 223(113001. 10.1016/j.physbeh.2020.113001

Hamm, J. D., Klatzkin, R. R., Herzog, M., Tamura, S., Brunstrom, J. M., & Kissileff, H. R. (2021). Recalled and momentary virtual portions created of snacks predict actual intake under laboratory stress condition. Physiol Behav. 238(113479. 10.1016/j.physbeh.2021.113479

Han, J. E., Frasnelli, J., Zeighami, Y., Larcher, K., Boyle, J., McConnell, T., Malik, S., Jones-Gotman, M., & Dagher, A. (2018). Ghrelin Enhances Food Odor Conditioning in Healthy Humans: An fMRI Study. Cell Rep. 25(10), 2643–2652 e4. 10.1016/j.celrep.2018.11.026

Hege, M. A., Veit, R., Krumsiek, J., Kullmann, S., Heni, M., Rogers, P. J., Brunstrom, J. M., Fritsche, A., & Preissl, H. (2018). Eating less or more - Mindset induced changes in neural correlates of pre-meal planning. Appetite. 125(492–501. 10.1016/j.appet.2018.03.006

Hengist, A., Sciarrillo, C. M., Guo, J., Walter, M., & Hall, K. D. (2024). Gut-derived appetite hormones do not explain energy intake differences in humans following low-carbohydrate versus low-fat diets. Obesity. 32(9), 1689–1698. 10.1002/oby.24104

Heni, M., Kullmann, S., Preissl, H., Fritsche, A., & Haring, H. U. (2015). Impaired insulin action in the human brain: causes and metabolic consequences. Nat Rev Endocrinol. 11(12), 701–11. 10.1038/nrendo.2015.173

Hola, L., Zelezna, B., Karnosova, A., Kunes, J., Fehrentz, J. A., Denoyelle, S., Cantel, S., Blechova, M., Sykora, D., Myskova, A., & Maletinska, L. (2022). A novel truncated liver enriched antimicrobial peptide-2 palmitoylated at its N-terminal antagonizes effects of ghrelin. J Pharmacol Exp Ther. 383(2), 129–136. 10.1124/jpet.122.001322

Holm, S. K., Vestergaard, E. T., Zubanovic, N. B., Byberg, S., Clemmensen, C., Holst, B., & Thomsen, H. H. (2023). Ketone monoester increases circulating levels of LEAP2 and decreases appetite in healthy men. Diabetes Obes Metab. 25(7), 2023–2027. 10.1111/dom.15044

Holt, S. H., Brand Miller, J. C., & Petocz, P. (1996). Interrelationships among postprandial satiety, glucose and insulin responses and changes in subsequent food intake. Eur J Clin Nutr. 50(12), 788–97.

Horner, K., Hopkins, M., Finlayson, G., Gibbons, C., & Brennan, L. (2020). Biomarkers of appetite: is there a potential role for metabolomics? Nutr Res Rev. 33(2), 271–286. 10.1017/S0954422420000062

Ishii, S., Osaki, N., & Shimotoyodome, A. (2016). The Effects of a Hypocaloric Diet on Diet-Induced Thermogenesis and Blood Hormone Response in Healthy Male Adults: A Pilot Study. J Nutr Sci Vitaminol (Tokyo). 62(1), 40–6. 10.3177/jnsv.62.40

Islam, M. N., Mita, Y., Maruyama, K., Tanida, R., Zhang, W., Sakoda, H., & Nakazato, M. (2020). Liver-expressed antimicrobial peptide 2 antagonizes the effect of ghrelin in rodents. J Endocrinol. 244(1), 13–23. 10.1530/JOE-19-0102

Islam, M. N., Zhang, W., Sakai, K., Nakazato, Y., Tanida, R., Sakoda, H., Takei, T., Takao, T., & Nakazato, M. (2022). Liver-expressed antimicrobial peptide 2 functions independently of growth hormone secretagogue receptor in calorie-restricted mice. Peptides. 151(170763. 10.1016/j.peptides.2022.170763

Jones, K. L., Horowitz, M., Berry, M., Wishart, J. M., & Guha, S. (1997). Blood glucose concentration influences postprandial fullness in IDDM. Diabetes Care. 20(7), 1141–6. 10.2337/diacare.20.7.1141

Kojima, M., & Kangawa, K. (2006). Drug insight: The functions of ghrelin and its potential as a multitherapeutic hormone. Nat Clin Pract Endocrinol Metab. 2(2), 80–88.

Krause, A., Sillard, R., Kleemeier, B., Kluver, E., Maronde, E., Conejo-Garcia, J. R., Forssmann, W. G., Schulz-Knappe, P., Nehls, M. C., Wattler, F., Wattler, S., & Adermann, K. (2003). Isolation and biochemical characterization of LEAP-2, a novel blood peptide expressed in the liver. Protein Sci. 12(1), 143–52. 10.1110/ps.0213603

Kullmann, S., Kleinridders, A., Small, D. M., Fritsche, A., Häring, H.-U., Preissl, H., & Heni, M. (2020). Central nervous pathways of insulin action in the control of metabolism and food intake. The Lancet Diabetes & Endocrinology. 8(6), 524–534. 10.1016/S2213-8587(20)30113-3

Labouebe, G., Liu, S., Dias, C., Zou, H., Wong, J. C., Karunakaran, S., Clee, S. M., Phillips, A. G., Boutrel, B., & Borgland, S. L. (2013). Insulin induces long-term depression of ventral tegmental area dopamine neurons via endocannabinoids. Nat Neurosci. 16(3), 300–8. 10.1038/nn.3321

Lauritsen, J. V., Bergmann, N., Junker, A. E., Gyldenlove, M., Skov, L., Gluud, L. L., Hartmann, B., Holst, J. J., Vilsboll, T., & Knop, F. K. (2023). Oral glucose has little or no effect on appetite and satiety sensations despite a significant gastrointestinal response. Eur J Endocrinol. 189(6), 619–626. 10.1093/ejendo/lvad161

Lavin, J. H., Wittert, G., Sun, W. M., Horowitz, M., Morley, J. E., & Read, N. W. (1996). Appetite regulation by carbohydrate: role of blood glucose and gastrointestinal hormones. Am J Physiol. 271(2 Pt 1), E209–14. 10.1152/ajpendo.1996.271.2.E209

le Roux, C. W., Batterham, R. L., Aylwin, S. J., Patterson, M., Borg, C. M., Wynne, K. J., Kent, A., Vincent, R. P., Gardiner, J., Ghatei, M. A., & Bloom, S. R. (2006). Attenuated peptide YY release in obese subjects is associated with reduced satiety. Endocrinology. 147(3-8.

le Roux, C. W., Patterson, M., Vincent, R. P., Hunt, C., Ghatei, M. A., & Bloom, S. R. (2005). Postprandial plasma ghrelin is suppressed proportional to meal calorie content in normal weight but not obese subjects. J Clin Endo Metab. 90(2), 1068–1071.

Lemmens, S. G., Martens, E. A., Kester, A. D., & Westerterp-Plantenga, M. S. (2011). Changes in gut hormone and glucose concentrations in relation to hunger and fullness. Am J Clin Nutr. 94(3), 717–25. 10.3945/ajcn.110.008631

Lim, J. J., Liu, Y., Lu, L. W., Sequeira, I. R., & Poppitt, S. D. (2023). No Evidence That Circulating GLP-1 or PYY Are Associated with Increased Satiety during Low Energy Diet-Induced Weight Loss: Modelling Biomarkers of Appetite. Nutrients. 15(10), 10.3390/nu15102399

Lim, J. J., & Poppitt, S. D. (2019). How Satiating Are the ’Satiety’ Peptides: A Problem of Pharmacology versus Physiology in the Development of Novel Foods for Regulation of Food Intake. Nutrients. 11(7), 10.3390/nu11071517

Lippl, F., Erdmann, J., Steiger, A., Lichter, N., Czogalla-Peter, C., Bidlingmaier, M., Tholl, S., & Schusdziarra, V. (2012). Low-dose ghrelin infusion - evidence against a hormonal role in food intake. Regul Pept. 174(1-3), 26–31. 10.1016/j.regpep.2011.11.005

Liu, A. G., Puyau, R. S., Han, H., Johnson, W. D., Greenway, F. L., & Dhurandhar, N. V. (2015). The effect of an egg breakfast on satiety in children and adolescents: a randomized crossover trial. J Am Coll Nutr. 34(3), 185–90. 10.1080/07315724.2014.942471

Liu, J., Prudom, C. E., Nass, R., Pezzoli, S. S., Oliveri, M. C., Johnson, M. L., Veldhuis, P., Gordon, D. A., Howard, A. D., Witcher, D. R., Geysen, H. M., Gaylinn, B. D., & Thorner, M. O. (2008). Novel ghrelin assays provide evidence for independent regulation of ghrelin acylation and secretion in healthy young men. J Clin Endocrinol Metab. 93(5), 1980–1987.

Lugilde, J., Casado, S., Beiroa, D., Cunarro, J., Garcia-Lavandeira, M., Alvarez, C. V., Nogueiras, R., Dieguez, C., & Tovar, S. (2022). LEAP-2 Counteracts Ghrelin-Induced Food Intake in a Nutrient, Growth Hormone and Age Independent Manner. Cells. 11(3), 324. 10.3390/cells11030324

Lyngstad, A., Nymo, S., Coutinho, S. R., Rehfeld, J. F., Truby, H., Kulseng, B., & Martins, C. (2019). Investigating the effect of sex and ketosis on weight-loss-induced changes in appetite. Am J Clin Nutr. 109(6), 1511–1518. 10.1093/ajcn/nqz002

M’Kadmi, C., Cabral, A., Barrile, F., Giribaldi, J., Cantel, S., Damian, M., Mary, S., Denoyelle, S., Dutertre, S., Peraldi-Roux, S., Neasta, J., Oiry, C., Baneres, J. L., Marie, J., Perello, M., & Fehrentz, J. A. (2019). N-Terminal Liver-Expressed Antimicrobial Peptide 2 (LEAP2) Region Exhibits Inverse Agonist Activity toward the Ghrelin Receptor. J Med Chem. 62(2), 965–973. 10.1021/acs.jmedchem.8b01644

Mackelvie, K. J., Meneilly, G. S., Elahi, D., Wong, A. C., Barr, S. I., & Chanoine, J. P. (2007). Regulation of appetite in lean and obese adolescents after exercise: role of acylated and desacyl ghrelin. J Clin Endocrinol Metab. 92(2), 648–54. 10.1210/jc.2006-1028

Malik, S., McGlone, F., Bedrossian, D., & Dagher, A. (2008). Ghrelin modulates brain activity in areas that control appetitive behavior. Cell Metab. 7(5), 400–409. 10.1016/j.cmet.2008.03.007

Mani, B. K., Puzziferri, N., He, Z., Rodriguez, J. A., Osborne-Lawrence, S., Metzger, N. P., Chhina, N., Gaylinn, B., Thorner, M. O., Thomas, E. L., Bell, J. D., Williams, K. W., Goldstone, A. P., & Zigman, J. M. (2019). LEAP2 changes with body mass and food intake in humans and mice. J Clin Invest. 129(9), 3909–3923. 10.1172/JCI125332

Martens, M. J., Lemmens, S. G., Born, J. M., & Westerterp-Plantenga, M. S. (2012). Satiating capacity and post-prandial relationships between appetite parameters and gut-peptide concentrations with solid and liquefied carbohydrate. PLoS One. 7(7), e42110. 10.1371/journal.pone.0042110

Matthews, D. R., Hosker, J. P., Rudenski, A. S., Naylor, B. A., Treacher, D. F., & Turner, R. C. (1985). Homeostasis model assessment: insulin resistance and beta-cell function from fasting plasma glucose and insulin concentrations in man. Diabetologia. 28(7), 412–419.

Mebel, D. M., Wong, J. C., Dong, Y. J., & Borgland, S. L. (2012). Insulin in the ventral tegmental area reduces hedonic feeding and suppresses dopamine concentration via increased reuptake. Eur J Neurosci. 36(3), 2336–46. 10.1111/j.1460-9568.2012.08168.x

Melanson, K. J., Westerterp-Plantenga, M. S., Campfield, L. A., & Saris, W. H. (1999). Blood glucose and meal patterns in time-blinded males, after aspartame, carbohydrate, and fat consumption, in relation to sweetness perception. Br J Nutr. 82(6), 437–46.

Mitchell, C. S., & Begg, D. P. (2021). The regulation of food intake by insulin in the central nervous system. J Neuroendocrinol. 33(4), e12952. 10.1111/jne.12952

Morris, L. S., Voon, V., & Leggio, L. (2018). Stress, Motivation, and the Gut-Brain Axis: A Focus on the Ghrelin System and Alcohol Use Disorder. Alcohol Clin Exp Res. 10.1111/acer.13781

Muller, T. D., Nogueiras, R., Andermann, M. L., Andrews, Z. B., Anker, S. D., Argente, J., Batterham, R. L., Benoit, S. C., Bowers, C. Y., Broglio, F., Casanueva, F. F., D’Alessio, D., Depoortere, I., Geliebter, A., Ghigo, E., Cole, P. A., Cowley, M., Cummings, D. E., Dagher, A., Diano, S., Dickson, S. L., Dieguez, C., Granata, R., Grill, H. J., Grove, K., Habegger, K. M., Heppner, K., Heiman, M. L., Holsen, L., Holst, B., Inui, A., Jansson, J. O., Kirchner, H., Korbonits, M., Laferrere, B., LeRoux, C. W., Lopez, M., Morin, S., Nakazato, M., Nass, R., Perez-Tilve, D., Pfluger, P. T., Schwartz, T. W., Seeley, R. J., Sleeman, M., Sun, Y., Sussel, L., Tong, J., Thorner, M. O., van der Lely, A. J., van der Ploeg, L. H., Zigman, J. M., Kojima, M., Kangawa, K., Smith, R. G., Horvath, T., & Tschop, M. H. (2015). Ghrelin. Mol Metab. 4(6), 437–60. 10.1016/j.molmet.2015.03.005

Nymo, S., Coutinho, S. R., Eknes, P. H., Vestbostad, I., Rehfeld, J. F., Truby, H., Kulseng, B., & Martins, C. (2018). Investigation of the long-term sustainability of changes in appetite after weight loss. Int J Obes. 42(8), 1489–1499. 10.1038/s41366-018-0119-9

Nymo, S., Coutinho, S. R., Jorgensen, J., Rehfeld, J. F., Truby, H., Kulseng, B., & Martins, C. (2017). Timeline of changes in appetite during weight loss with a ketogenic diet. Int J Obes. 41(8), 1224–1231. 10.1038/ijo.2017.96

Peos, J. J., Helms, E. R., Fournier, P. A., Ong, J., Hall, C., Krieger, J., & Sainsbury, A. (2021). Continuous versus Intermittent Dieting for Fat Loss and Fat-Free Mass Retention in Resistance-trained Adults: The ICECAP Trial. Med Sci Sports Exerc. 53(8), 1685–1698. 10.1249/MSS.0000000000002636

Perello, M., & Dickson, S. L. (2015). Ghrelin signalling on food reward: a salient link between the gut and the mesolimbic system. J Neuroendocrinol. 27(6), 424–34. 10.1111/jne.12236

Pilichiewicz, A. N., Chaikomin, R., Brennan, I. M., Wishart, J. M., Rayner, C. K., Jones, K. L., Smout, A. J., Horowitz, M., & Feinle-Bisset, C. (2007). Load-dependent effects of duodenal glucose on glycemia, gastrointestinal hormones, antropyloroduodenal motility, and energy intake in healthy men. Am J Physiol Endocrinol Metab. 293(3), E743–53. 10.1152/ajpendo.00159.2007

Raben, A., Agerholm-Larsen, L., Flint, A., Holst, J. J., & Astrup, A. (2003). Meals with similar energy densities but rich in protein, fat, carbohydrate, or alcohol have different effects on energy expenditure and substrate metabolism but not on appetite and energy intake. Am J Clin Nutr. 77(1), 91–100. 10.1093/ajcn/77.1.91

Ragland, T. J., & Malin, S. K. (2023). Plasma LEAP-2 Following a Low-Calorie Diet with or without Interval Exercise in Women with Obesity. Nutrients. 15(3), 655. 10.3390/nu15030655

Rodin, J., Wack, J., Ferrannini, E., & DeFronzo, R. A. (1985). Effect of insulin and glucose on feeding behavior. Metabolism. 34(9), 826–31. 10.1016/0026-0495(85)90106-4

Rolls, B. J. (1986). Sensory-specific satiety. Nutr Rev. 44(3), 93–101. 10.1111/j.1753-4887.1986.tb07593.x

Rolls, B. J., Roe, L. S., Cunningham, P. M., Keller, K. L., & Zuraikat, F. M. (2025). High satiety: Evaluating determinants of energy compensation and intake in multiple preloading studies. Appetite. 213(108036. 10.1016/j.appet.2025.108036

Romero, A., Kirchner, H., Heppner, K., Pfluger, P. T., Tschop, M. H., & Nogueiras, R. (2010). GOAT: the master switch for the ghrelin system? Eur J Endocrinol. 163(1), 1–8.

Roth, C. L., Melhorn, S. J., De Leon, M. R. B., Rowland, M. G., Elfers, C. T., Huang, A., Saelens, B. E., & Schur, E. A. (2022). Impaired Brain Satiety Responses After Weight Loss in Children With Obesity. J Clin Endocrinol Metab. 107(8), 2254–2266. 10.1210/clinem/dgac299

Schultes, B., Panknin, A. K., Hallschmid, M., Jauch-Chara, K., Wilms, B., de Courbiere, F., Lehnert, H., & Schmid, S. M. (2016). Glycemic increase induced by intravenous glucose infusion fails to affect hunger, appetite, or satiety following breakfast in healthy men. Appetite. 105(562–6. 10.1016/j.appet.2016.06.032

Schulz, C., Vezzani, C., & Kroemer, N. B. (2023). How gut hormones shape reward: a systematic review of the role of ghrelin and GLP-1 in human fMRI. Physiol Behav. 263(114111. 10.1016/j.physbeh.2023.114111

Service, F. J., Hall, L. D., Westland, R. E., O’Brien, P. C., Go, V. L., Haymond, M. W., & Rizza, R. A. (1983). Effects of size, time of day and sequence of meal ingestion on carbohydrate tolerance in normal subjects. Diabetologia. 25(4), 316–21. 10.1007/BF00253193

Shankar, K., Metzger, N. P., Lawrence, C., Gupta, D., Osborne-Lawrence, S., Varshney, S., Singh, O., Richard, C. P., Zaykov, A. N., Rolfts, R., DuBois, B. N., Perez-Tilve, D., Mani, B. K., Hammer, S. T. G., & Zigman, J. M. (2024). A long-acting LEAP2 analog reduces hepatic steatosis and inflammation and causes marked weight loss in mice. Mol Metab. 84(101950. 10.1016/j.molmet.2024.101950

Shankar, K., Metzger, N. P., Singh, O., Mani, B. K., Osborne-Lawrence, S., Varshney, S., Gupta, D., Ogden, S. B., Takemi, S., Richard, C. P., Nandy, K., Liu, C., & Zigman, J. M. (2021). LEAP2 deletion in mice enhances ghrelin’s actions as an orexigen and growth hormone secretagogue. Mol Metab. 53(101327. 10.1016/j.molmet.2021.101327

Speechly, D. P., & Buffenstein, R. (2000). Appetite dysfunction in obese males: evidence for role of hyperinsulinaemia in passive overconsumption with a high fat diet. Eur J Clin Nutr. 54(3), 225–33. 10.1038/sj.ejcn.1600924

St-Onge, M. P., Mayrsohn, B., O’Keeffe, M., Kissileff, H. R., Choudhury, A. R., & Laferrere, B. (2014). Impact of medium and long chain triglycerides consumption on appetite and food intake in overweight men. Eur J Clin Nutr. 68(10), 1134–40. 10.1038/ejcn.2014.145

Steinert, R. E., Feinle-Bisset, C., Asarian, L., Horowitz, M., Beglinger, C., & Geary, N. (2017). Ghrelin, CCK, GLP-1, and PYY(3-36): Secretory Controls and Physiological Roles in Eating and Glycemia in Health, Obesity, and After RYGB. Physiol Rev. 97(1), 411–463. 10.1152/physrev.00031.2014

Stengel, A., Wang, L., & Tache, Y. (2011). Stress-related alterations of acyl and desacyl ghrelin circulating levels: mechanisms and functional implications. Peptides. 32(11), 2208–2217.

Suarez, A. N., Noble, E. E., & Kanoski, S. E. (2019). Regulation of Memory Function by Feeding-Relevant Biological Systems: Following the Breadcrumbs to the Hippocampus. Front Mol Neurosci. 12(101. 10.3389/fnmol.2019.00101

Thomas, D. M., Flore, C., & Ludwig, D. S. (2026). Inappropriate analyses make continuous glucose monitoring appear imprecise. Am J Clin Nutr. 10.1016/j.ajcnut.2026.101388

Thompson, D. A., & Campbell, R. G. (1977). Hunger in humans induced by 2-deoxy-D-glucose: glucoprivic control of taste preference and food intake. Science. 198(4321), 1065–8. 10.1126/science.929188

Tiedemann, L. J., Schmid, S. M., Hettel, J., Giesen, K., Francke, P., Buchel, C., & Brassen, S. (2017). Central insulin modulates food valuation via mesolimbic pathways. Nat Commun. 8(16052. 10.1038/ncomms16052

Tiffany, S. T., & Drobes, D. J. (1991). The development and initial validation of a questionnaire on smoking urges. Br J Addict. 86(11), 1467–76.

Tolle, V., Tezenas du Montcel, C., Mattioni, J., Schéle, E., Viltart, O., & Dickson, S. L. (2024). To eat or not to eat: A role for ghrelin and LEAP2 in eating disorders? Neuroscience Applied. 3(104045. 10.1016/j.nsa.2024.104045

Unick, J. L., Otto, A. D., Goodpaster, B. H., Helsel, D. L., Pellegrini, C. A., & Jakicic, J. M. (2010). Acute effect of walking on energy intake in overweight/obese women. Appetite. 55(3), 413–9. 10.1016/j.appet.2010.07.012

Veit, R., Horstman, L. I., Hege, M. A., Heni, M., Rogers, P. J., Brunstrom, J. M., Fritsche, A., Preissl, H., & Kullmann, S. (2020). Health, pleasure, and fullness: changing mindset affects brain responses and portion size selection in adults with overweight and obesity. Int J Obes (Lond). 44(2), 428–437. 10.1038/s41366-019-0400-6

Verdich, C., Toubro, S., Buemann, B., Lysgard, M. J., Juul, H. J., & Astrup, A. (2001). The role of postprandial releases of insulin and incretin hormones in meal-induced satiety--effect of obesity and weight reduction. Int J Obes Relat Metab Disord. 25(8), 1206–1214.

Wang, J. H., Li, H. Z., Shao, X. X., Nie, W. H., Liu, Y. L., Xu, Z. G., & Guo, Z. Y. (2019). Identifying the binding mechanism of LEAP2 to receptor GHSR1a. FEBS J. 286(7), 1332–1345. 10.1111/febs.14763

Wang, Y., Wu, Q., Zhou, Q., Chen, Y., Lei, X., Chen, Y., & Chen, Q. (2022). Circulating acyl and des-acyl ghrelin levels in obese adults: a systematic review and meta-analysis. Sci Rep. 12(1), 2679. 10.1038/s41598-022-06636-3

Woo, R., Kissileff, H. R., & Pi-Sunyer, F. X. (1984). Elevated postprandial insulin levels do not induce satiety in normal-weight humans. Am J Physiol. 247(4 Pt 2), R745–9. 10.1152/ajpregu.1984.247.4.R745

Wren, A. M., Seal, L. J., Cohen, M. A., Brynes, A. E., Frost, G. S., Murphy, K. G., Dhillo, W. S., Ghatei, M. A., & Bloom, S. R. (2001). Ghrelin enhances appetite and increases food intake in humans. J Clin Endo Metab. 86(12), 5992–5995.

Wyatt, P., Berry, S. E., Finlayson, G., O’Driscoll, R., Hadjigeorgiou, G., Drew, D. A., Khatib, H. A., Nguyen, L. H., Linenberg, I ., Chan, A. T., Spector, T. D., Franks, P. W., Wolf, J., Blundell, J., & Valdes, A. M. (2021). Postprandial glycaemic dips predict appetite and energy intake in healthy individuals. Nat Metab. 3(4), 523–529. 10.1038/s42255-021-00383-x

