## Supplementary Material for "Relationships between increasing pre-load meal size, post-prandial acyl ghrelin, LEAP2, insulin, glucose, appetite, and food intake in adults without obesity"

Tasya Parastika [1], Jiaxun Liu [1], Raghav Bhargava [1], Wai In Ng [1], Jialin Guo [1], Marcela Rodriguez Flores [1], Xinyi Zhang [1], Mimoza Emini [1], Wanqian Li [1], Sandra Luur [1], Zoe Barley [1], Jeff Brunstrom [2], Anthony P Goldstone [1]

[1] PsychoNeuroEndocrinology Research Group, Division of Psychiatry, Department of Brain Sciences, Imperial College London, Hammersmith Hospital, London, UK

[2] School of Psychology and Neuroscience, University of Bristol, UK

**SUPPLEMENTARY MATERIAL**

Supplementary Figure S3. Correlations of relative preload meal size with blood glucose and hormones. 13

**SUPPLEMENTARY METHODS**

**Inclusion Criteria**

(i) Male or female between the ages of 18 and 60 years.

(ii) Without obesity with body mass index (BMI) 18.0-29.9 kg/m$^2$ or with obesity with BMI 30.0-50.0 kg/m$^2$.

(iii) Healthy as determined by medical history and vital signs.

(iv) Capable of giving written informed consent, which includes compliance with the requirements and restrictions listed in the consent form.

(v) Participant is able to read, comprehend and record information written in English.

(vi) A signed and dated written informed consent is obtained from the participant.

**Exclusion Criteria**

(i) History of, or current abuse or dependence on alcohol or drugs.

(ii) Current smoker or less than 2 years since quitting (cigarette, cigars, e-cigarettes) or use of nicotine replacement therapy.

(iii) Significant current or past medical or psychiatric history that, in the opinion of the investigators, contraindicates their participation.

(iv) History of type 1 or type 2 diabetes mellitus.

(v) History of ischaemic heart disease, heart failure, cardiac arrhythmia, peripheral vascular, cerebrovascular disease or uncontrolled hypertension.

(vi) Current diagnosis of anaemia or iron deficiency.

(vii) Body weight instability (change in body weight of > 5% over the preceding 3 months).

(viii) Unwillingness or inability to follow the procedures outlined in the protocol.

(ix) Use of current regular prescription or over-the-counter medications that in the opinion of the Investigators may affect participant safety or outcome measures.

(x) Clinically significant abnormalities in screening blood tests abnormalities which in the opinion of the study physician, is clinically significant e.g. diabetes mellitus, hypothyroidism, renal impairment, abnormal liver function tests [bilirubin, alanine transaminase (ALT), aspartate transaminase (AST), gamma-glutamyl transferase (GGT)] >3x upper limit of normal, other than due to fatty liver disease.

(xi) Current pregnancy or breast-feeding in female volunteers (the Investigators will recommend using contraception for the duration of the visits to avoid participant drop-out).

(xii) Pulse rate <40 or >100 beats per minute OR systolic blood pressure >160 and <100 OR diastolic blood pressure >95 and <50 in the semi-supine position.

(xiii) The volunteer has participated in a clinical trial and has received an investigational product within the following time period prior to the first experimental visit in the current study: 90 days, 5 halflives or twice the duration of the biological effect of the investigational product (whichever is longer).

(xiv) Exposure to more than 3 new investigational medicinal products within 12 months prior to the scan.

(xv) Vegan, gluten or lactose-intolerant (as test meals in the paradigms may include animal products, dairy and wheat products).

(xvi) Volunteers who have donated, or intend to donate, blood within three months before the screening visit or following study visit completion.

(xvii) Known history of SARS-CoV-2 infection (Covid-19) in the last 4 weeks.

(xviii) Ongoing symptoms suggestive of complications from previous SARS-CoV-2 infection ('long Covid-19') such as loss or change in sense of smell or taste, shortness of breath, palpitations, lethargy.

(xix) SCOFF questionnaire score >1/5 indicating eating disorders (Luck et al. 2002).

(xx) DSM-V criteria for alcohol use disorder (AUD) >2/11 indicating mild AUD (American-Psychiatric-Association 2013).

(xxi) Participants who have had previous obesity/bariatric surgery or are on medications for obesity.

(xxii) Dutch Eating Behaviour Questionnaire (DEBQ) restraint score >3/5 indicating highly restrained eating behaviour (van Strien et al. 1986).

### SUPPLEMENTARY RESULTS

#### Effects of pre-load meal size on outcomes using exploratory post-prandial calculations

When analysing correlations of *absolute* pre-load meal size with post-prandial appetite ratings, glucose and hormone outcome values other than the pre-registered $iAUC_{0\text{-}3h}$ measure, this measure only gave the strongest correlation for plasma LEAP2, with second strongest Δ0-maximum 3h value (**Supplementary Figure S18A**).

For hunger, composite appetite, food craving and fullness VAS ratings, the strongest correlations were with absolute value at 3h, and second strongest with $AUC_{0\text{-}3h}$ value, except for food craving which was with Δ0-3h value (**Supplementary Figure S18A**). Regarding blood measures, for glucose and insulin the strongest correlations were with maximum 0.5-3h value, and second strongest with Δ0-maximum 0.5-3h; while for AG and AG/LEAP2 ratio, strongest correlations were with $AUC_{0\text{-}3h}$ value, and second strongest with minimum 0.5-3h value (**Supplementary Figure S18A**).

Similar results were seen for correlations with *relative* pre-load meal size (Supplementary Figure S18B), except that for insulin, the second strongest correlation was now with $AUC_{0\text{-}3h}$, and for AG, the strongest correlation was now with $iAUC_{0\text{-}3h}$, and second strongest with $AUC_{0\text{-}3h}$.

As a control check, there were no significant correlations of preload meal size (absolute or relative) with 0 min baseline appetite ratings nor blood measures, indicating that there was no systematic bias by chance in these measures before the pre-load meal was administered, related to the subsequent meal size (**Supplementary Figure S18A,B**).

#### Relationships of desired food intake with outcomes using exploratory post-prandial calculations

When analysing correlations of *absolute* desired food intake with appetite ratings, blood glucose and hormones outcome values, the pre-registered $iAUC_{0\text{-}2h}$ measure did not give the strongest correlation for any of the outcomes (**Supplementary Figure S19A-B**). For hunger, composite appetite, food craving and fullness ratings, the strongest correlations were with absolute value at 2h, and second strongest with $AUC_{0\text{-}2h}$ value (**Supplementary Figure S19A-B**). Regarding blood measures, for glucose and insulin the strongest correlations were with maximum 0.5-2h value, and second strongest with $AUC_{0\text{-}2h}$. For AG, the strongest correlation was with Δ0-2h value, and second strongest with absolute 2h value. By contrast, for AG/LEAP2 ratio, the strongest correlation was with absolute 2h value, and second strongest with maximum 0.5-2h value (**Supplementary Figure S19A**).

Similar results were seen for correlations with *relative* desired food intake (**Supplementary Figure S19B**), except that for insulin, the second strongest correlation was now with Δ0-maximum 2h value, and for both

AG and AG/LEAP2 ratio, the strongest correlations were now with absolute 2h value, and second strongest with maximum 0.5-2h value.

As a control check, there were no significant correlations of desired food intake (absolute or relative) with 0 min baseline appetite ratings nor blood measures, indicating that there was no systematic bias by chance in these measures before the VPCT was tested (**Supplementary Figure S19A-B**).

**Relationships of actual food intake with outcomes using exploratory post-prandial calculations**

When analysing correlations of *absolute* actual food intake with appetite ratings, blood glucose and hormones outcome values, the pre-registered $iAUC_{0\text{-}3h}$ measure did not give the strongest correlation for any of the outcomes (Supplementary Figure S20A). For hunger, composite appetite and food craving ratings, the strongest correlations were with absolute value at 3h, while the second strongest values were maximum 0.5-3h, AUC0-3h and maximum 0.5-3h respectively. For fullness ratings, the strongest correlation was with maximum 0.5-3h value, and second strongest with absolute 2h value (**Supplementary Figure S20A**).

Regarding blood measures, for glucose and insulin, the strongest correlations were with maximum 0.5-2h value, while the second strongest were absolute value 2h and Δ0-maximum 3h value respectively (**Supplementary Figure S20A**). For AG, the strongest correlations were with $AUC_{0\text{-}3h}$ and maximum 0.5-3h. For AG/LEAP2 ratio, the strongest correlation was with $AUC_{0\text{-}3h}$, and second strongest with absolute 2h value (**Supplementary Figure S20A**).

Similar results were seen for correlations with *relative* actual food intake (Supplementary Figure S20B), except that for fullness rating the order was the other way around, with the strongest correlation now with absolute 2h value, and second strongest correlation now with maximum 0.5-3h value. For AG and AG/LEAP2 ratio, the strongest correlation remained with $AUC_{0\text{-}3h}$ value, but the second strongest was now with maximum 0.5-3h value.

As a control check, there were no significant correlations of actual food intake (absolute or relative) with 0 min baseline appetite ratings nor blood measures, indicating that there was no systematic bias by chance in these measures before the *ad libitum* meals was administered (**Supplementary Figure S20A-B**).

**Relationships of appetite ratings with blood outcomes using exploratory post-prandial calculations**

When analysing correlations of blood hormones with appetite ratings of hunger, for glucose the strongest correlation was with Δ0-maximum 0.5-3h value, and second strongest with $iAUC_{0\text{-}3h}$; for insulin the strongest correlation was with $iAUC_{0\text{-}3h}$, and second strongest with $AUC_{0\text{-}3h}$. Results for AG and AG/LEAP2 ratio were similar, whereby the strongest correlations were with $AUC_{0-3h}$ value, and second strongest with absolute 2h value (**Supplementary Figure S21A**).

For correlations of composite appetite, for plasma glucose, the strongest correlation was with $iAUC_{0-3h}$ value, and second strongest with absolute 2h value; for insulin, the strongest correlation was with $AUC_{0-3h}$, and second strongest with absolute 2h value. Whilst for AG and AG/LEAP2 ratio, there were similar results, with the strongest correlation with $AUC_{0-3h}$ value, and second strongest with absolute 2h value (**Supplementary Figure S21B**).

For correlations with food craving, for both glucose and insulin, the strongest correlations were with $iAUC_{0-3h}$ value, and second strongest with Δ0-maximum 0.5-3h value (**Supplementary Figure S21C**). For AG and AG/LEAP2 ratio, the strongest correlations were with $AUC_{0-3h}$ value, and second strongest with absolute 3h and 2h value respectively; while for LEAP2, there was only a trend for a correlation with absolute 3h value (**Supplementary Figure S21C**).

For fullness, for plasma glucose the strongest correlation was with absolute 3h value, and second strongest with Δ0-maximum 0.5-3h value; for insulin, the strongest correlation was with $AUC_{0-3h}$, and second strongest with maximum 0.5-3h value; for AG, the strongest correlation was with maximum 0.5-3h value, and second strongest with $AUC_{0-3h}$ value; whilst for AG/LEAP2 ratio the order of correlations was the other way around, with strongest $AUC_{0-3h}$ value, and second strongest maximum 0.5-3h value (**Supplementary Figure S21D**).

**SUPPLEMENTARY TABLES**

| | | |
|---|---|---|
| Breakfast (days/week) | Median [IQR] | 7 [3.9, 6.3] |
| | Range | 0 – 7 |
| Lunch (days/week) | Median [IQR] | 7 [5.7, 7.1] |
| | Range | 3 – 7 |
| BDI-II (0-63) | Median [IQR] | 1 [1.0, 4.7] |
| | Range | 0 – 10 |
| DEBQ-restraint (0-5) | Mean ± SD | 1.9 ± 0.8 |
| | Range | 0.9 – 3.4 |
| TFEQ-total restraint (0-21) | Mean ± SD | 5.5 ± 2.1 |
| | Range | 2 – 11 |
| TFEQ-flexible restraint (0-7) | Mean ± SD | 1.7 ± 1.2 |
| | Range | 0 – 5 |
| TFEQ-rigid restraint (0-7) | Mean ± SD | 1.5 ± 0.9 |
| | Range | 0 – 4 |
| EDE-Q-restraint (0-5) | Median [IQR] | 0 [0.2, 0.9] |
| | Range | 0 – 2.4 |
| DEBQ-emotional (0-5) | Mean ± SD | 1.6 ± 0.6 |
| | Range | 1 – 2.6 |
| DEBQ-external (0-5) | Mean ± SD | 2.9 ± 0.50 |
| | Range | 2.1 – 3.7 |
| TFEQ-disinhibition (0-16) | Mean ± SD | 4.5 ± 1.6 |
| | Range | 2 – 8 |
| TFEQ-hunger (0-14) | Mean ± SD | 3.9 ± 2.5 |
| | Range | 0 – 10 |
| EDE-Q eating concern (0-5) | Median [IQR] | 0.63 [0.39, 1.44] |
| | Range | 0 – 0.75 |
| EDE-Q weight concern (0-5) | Median [IQR] | 0.63 [0.39, 1.44] |
| | Range | 0 – 3.8 |
| EDE-Q shape concern (0-5) | Median [IQR] | 0.63 [0.39, 1.44] |
| | Range | 0 – 4 |
| EDE-Q global score (0-6) | Median [IQR] | 0.41 [0.27, 0.91] |
| | Range | 0 – 2.29 |
| AUDIT (0-40) | Mean ± SD | 3.8 ± 1.9 |
| | Range | 1 – 8 |

**Supplementary Table S1. Mood and eating behaviour questionnaires**

Normally distributed data are shown as mean ± SD or median [IQR], and range (minimum to maximum). Abbreviations: AUDIT, Alcohol Use Disorders Identification Test (Saunders et al. 1993); BDI-II, Beck Depression Inventory II (Beck et al. 1996); DEBQ, Dutch Eating Behaviour Questionnaire (van Strien et al. 1986); EDE-Q, Eating Disorders Examination Questionnaire (Fairburn and Beglin 1994); IQR, interquartile range; max, maximum; SD, standard deviation; TFEQ, Three Factor Eating Questionnaire (Stunkard and Messick 1985).

| | Savoury Low-fat | | Savoury High-fat | | Sweet Low-fat | Sweet high-fat |
|---|---|---|---|---|---|---|
| | Chicken Broth (Baxters) | Tomato Broth (Baxters) | Cream of Chicken Soup (Baxters) | Cream of Tomato Soup (Baxters) | Vanilla Yoghurt (Yeo Valley) | Vanilla Ice Cream (Haagen Dazs) |
| Energy density (kcal/100g) | 42.67 | 38.22 | 103.12 | 111.70 | 81.00 | 251.00 |
| Fat (g/100g) | 0.62 | 0.80 | 8.05 | 8.22 | 0.00 | 17.00 |
| of which saturated (g/100g) | 0.18 | 0.18 | 4.64 | 4.59 | 0.00 | 10.40 |
| Carbohydrates (g/100g) | 6.76 | 6.22 | 5.26 | 8.21 | 14.10 | 20.00 |
| of which sugars (g/100g) | 1.51 | 2.67 | 1.06 | 5.36 | 14.00 | 18.80 |
| Fibre (g/100g) | 1.16 | 0.44 | 0.09 | 0.18 | 0.00 | 0.00 |
| Protein (g/100g) | 1.87 | 1.42 | 2.12 | 1.06 | 5.80 | 4.30 |
| Salt (g/100g) | 0.56 | 0.77 | 0.57 | 0.60 | 0.17 | 0.18 |
| Amount served (g) | 100 | 100 | 900 | 900 | 900 | 920 |
| Amount served (kcal) | 341.3 | 305.8 | 928 | 1005 | 729 | 2309 |

**Supplementary Table S2. Nutritional information for dishes at *ad libitum* meal.**

Default choices were chicken broth, cream of chicken soup, vanilla yogurt and vanilla ice cream, but if participants disliked chicken broth and soup (determined by ratings at screening visit), tomato broth and soup were given instead (n=1).

| Dependent factor | Independent factor | Absolute or relative kcal | N | Preload x female sex interaction | | | Main effect of female sex | | | | | | |
|---|---|---|---|---|---|---|---|---|---|---|---|---|---|
| | | | | df | F | P | df | F | β | SEM | 95% CI | Cohen's d | P |
| **VPCT average** | **Preload meal size** | **absolute** | 15 | 1, 43.03 | 0.19 | 0.66 | 1, 21.90 | 2.31 | -356.1 | 234.5 | -842.5, 130.3 | -0.39 | 0.14 |
| | | **relative** | 15 | 1, 43.16 | 0.80 | 0.38 | 1, 22.65 | 1.16 | -16.89 | 15.68 | -59.36, 15.59 | -0.28 | 0.29 |
| ***ad libitum* meal total** | **Preload meal size** | **absolute** | 11 | 1, 32.08 | 0.40 | 0.53 | 1,28.48 | 0.05 | -38.91 | 174.98 | -397.05, 319.24 | -0.07 | 0.83 |
| | | **relative** | 11 | 1, 32.23 | 0.52 | 0.48 | 1, 26.83 | 0.09 | 3.57 | 11.73 | -20.50, 27.64 | 0.09 | 0.76 |

**Supplementary Table S3. Impact of sex on desired and acttual food intake.**

Linear mixed model analysis examining relationship between absolute (kcal) or relative (kcal as % of estimated resting energy expenditure) preload meal size, female sex(and preload x female sex interaction on respectively absolute, or relative average desired food intake from virtual portion size creation task (VPCT, 15 participants, 7 females, 58 visits) or total actual food intake (*ad libitum* meal, 11 participants, 4 females, 43 visits).

### SUPPLEMENTARY FIGURES

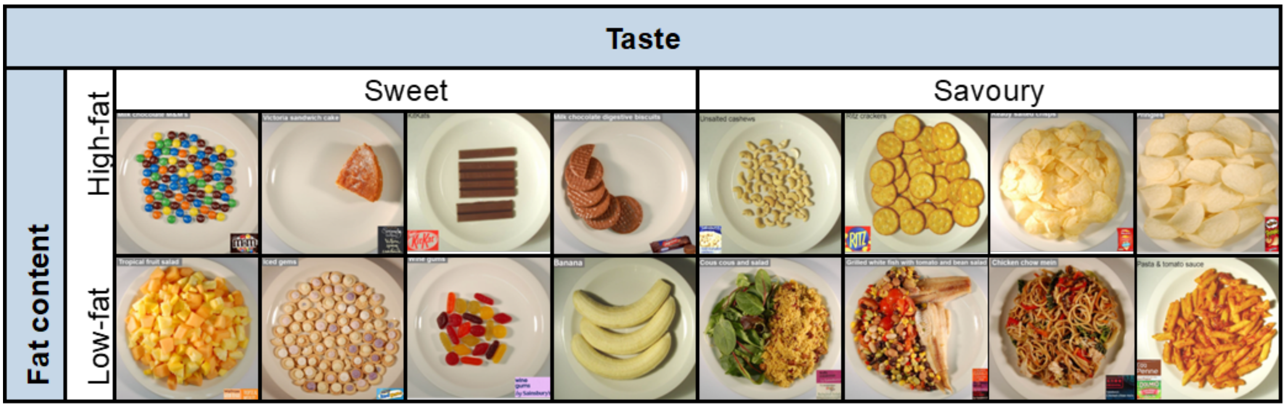

**Supplementary Figure S1. Pictures in virtual portion size creation task.**

Pictures of food presented in the virtual portion creation task (VPCT) by categories of sweetness (sweet or savoury) in columns, and fat content (high-fat or low-fat) in rows.

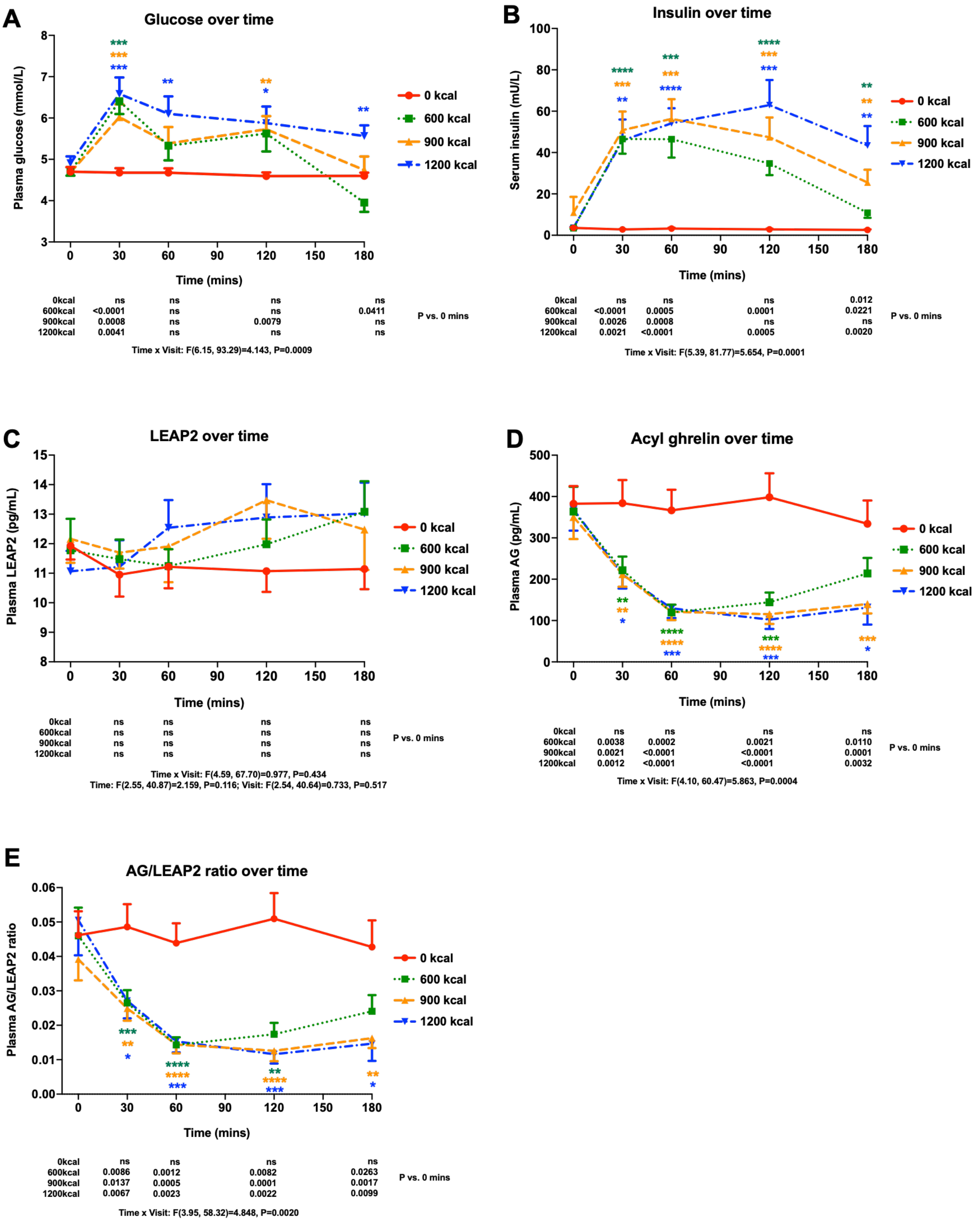

**Supplementary Figure S2. Absolute blood glucose and hormones over time at study visits.** Absolute (A) plasma glucose, (B) serum insulin, and plasma (C) LEAP2, (D) AG and (E) AG/LEAP2 ratio over time at four study visits with different pre-load meal size (0, 600, 900, 1200 kcal), at individual time points over visit. Data expressed as mean ± SEM, n=15-17. Statistical results from general linear model for interaction of time x visit given beneath graph. Coloured symbols within graph indicate P value for post-hoc Dunnet's test vs. 0 kcal visit at each time point: *P<0.05, **P<0.01, ***P<0.0005, ****P<0.0001; numbers immediately below graph indicate P value for post-hoc Dunnet's test vs. 0 min time point for each visit (with Greenhouse-Geisser correction). Abbreviations: ns, non-significant (P>0.05). To convert glucose from mmol/L to mg/dL multiple by 18.02; to convert insulin from mU/L to pmol/L multiple by 6.94.

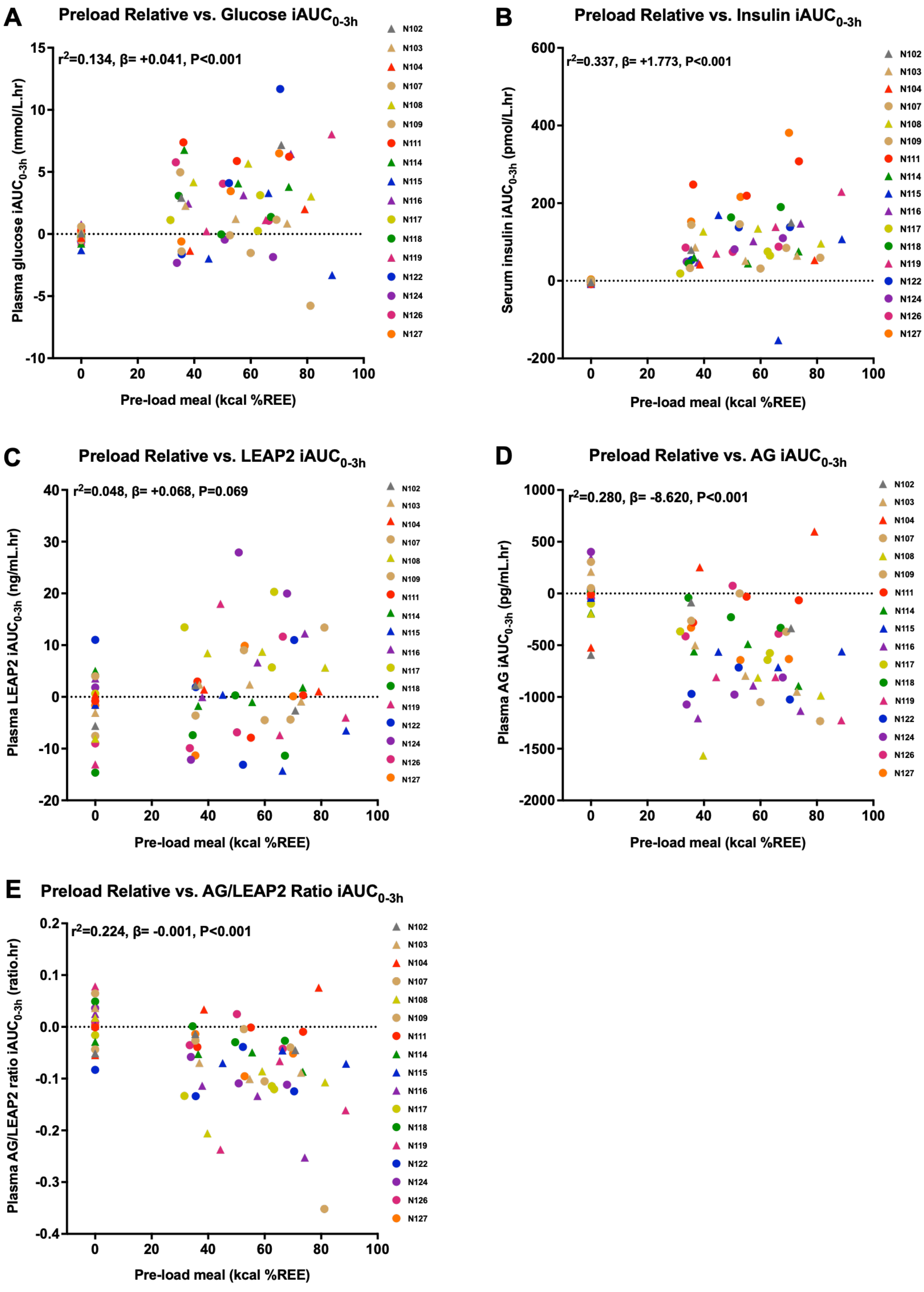

**Supplementary Figure S3. Correlations of relative preload meal size with blood glucose and hormones.** Effects of relative pre-load meal size over four study visits (kcal % relative energy expenditure) with (A) plasma glucose, (B) serum insulin, and plasma (C) LEAP2, (D) AG and (E) AG/LEAP2 molar ratio as total iAUC$_{0\text{-}3h}$, with marginal value of pseudo-R squared ($r^2$), P value, and estimated mean of β parameter from linear mixed model analysis based on data from 17 participants, 66 visits, except for (C-E) 65 visits. Males are represented as circles and females as triangles. Abbreviations: iAUC, incremental area under the curve. To convert glucose from mmol/L to mg/dL multiple by 18.02; to convert insulin from mU/L to pmol/L multiple by 6.94.

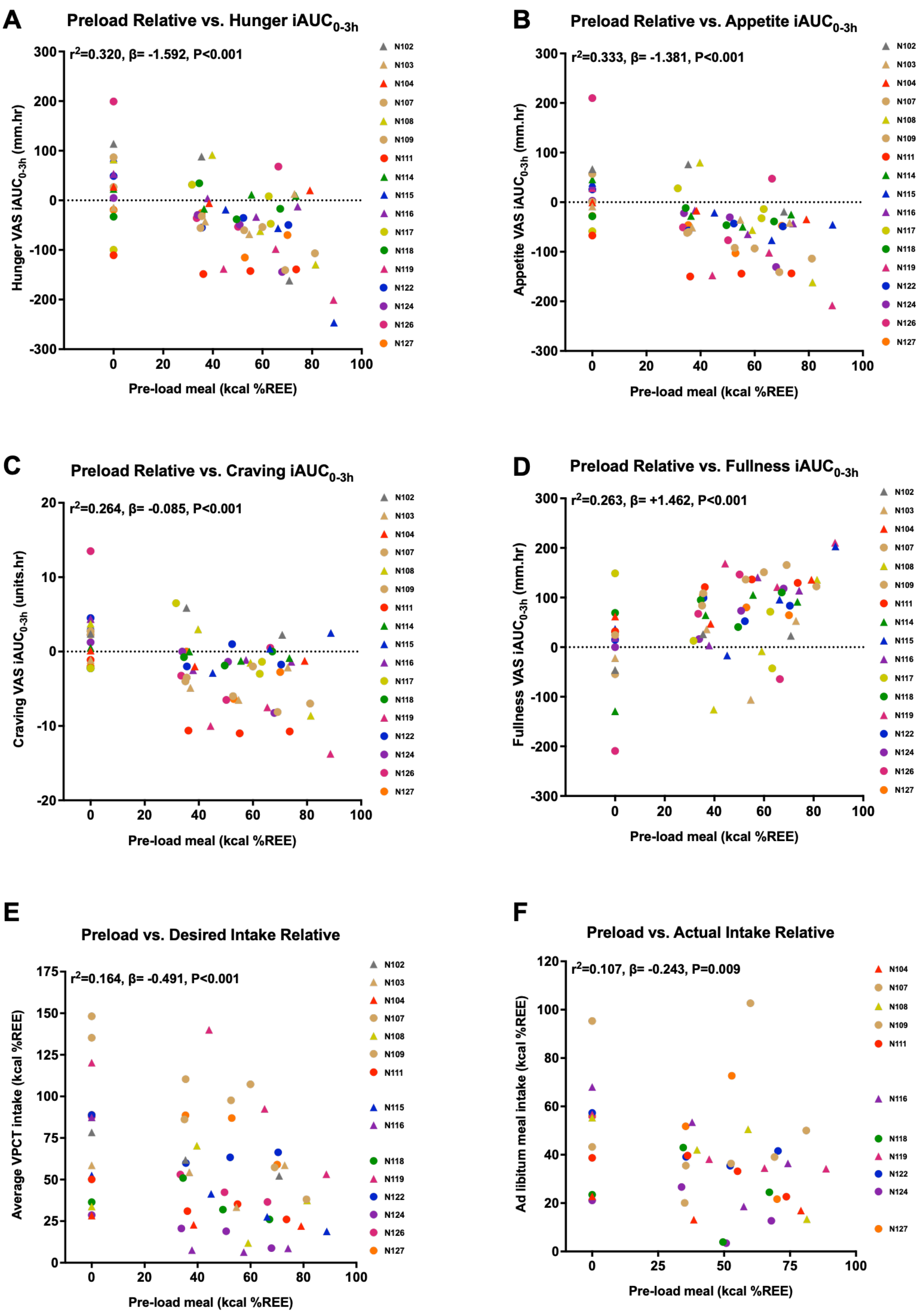

**Supplementary Figure S4. Correlations of relative preload meal size with appetite ratings, desired and actual food intake.**

Correlations of relative pre-load meal size (kcal % relative energy expenditure) over 4 study visits with (A) hunger, (B) composite appetite, (C) food craving, (D) fullness, from VAS ratings as total $iAUC_{0-3h}$, and relative (E) desired food intake from VPCT, (F) actual food intake from *ad libitum* meal, as kcal % relative energy expenditure. Marginal value of pseudo-R squared ($r^2$), P-value, and estimated mean of β parameter from linear mixed model analysis based on data from (A-D) 17 participants, 66 visits, (E) 15 participants, 58 visits, (F) 11 participants, 43 visits. Males are represented as circles and females as triangles. Abbreviations: iAUC, incremental area under the curve; VAS, visual analogue scale; VPCT, virtual portion size creation task.

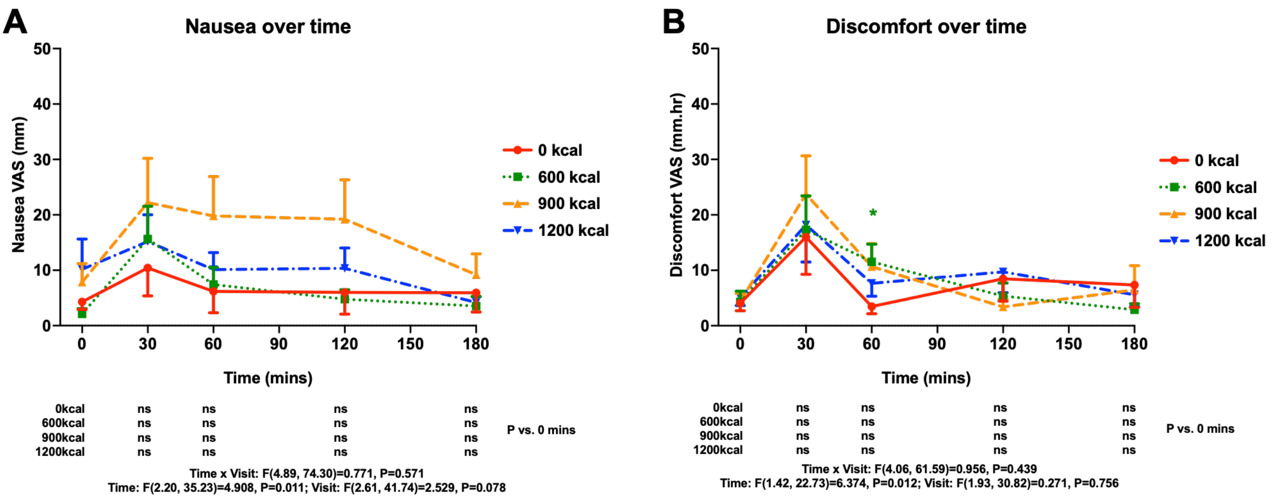

**Supplementary Figure S5. Absolute visual analogue scale ratings of aversive symptoms at visits.** Visual analogue scale (VAS) ratings over time for (A) nausea (0-100mm) and (B) discomfort (0-100mm) at individual time points at each of four study visits with pre-load given at 0 min: 0 kcal (red circle, solid line), 600 kcal (green square, dotted line), 900 kcal (yellow triangle, dashed line), 1200 kcal (blue inverted triangle dashed-dotted line). Data expressed as mean ± SEM, 17 participants, 66 visits. Statistical results from general linear model for interaction of time x visit given beneath graph. Coloured symbols within graph indicate P value for post-hoc Dunnet's test vs. 0 kcal visit at each time point: *P<0.05, **P<0.01, ***P<0.0005, ****P<0.0001; numbers immediately below graph indicate P value for post-hoc Dunnet's test vs. 0 min time point for each visit (with Greenhouse-Geisser correction). Abbreviations: ns, non-significant (P>0.05).

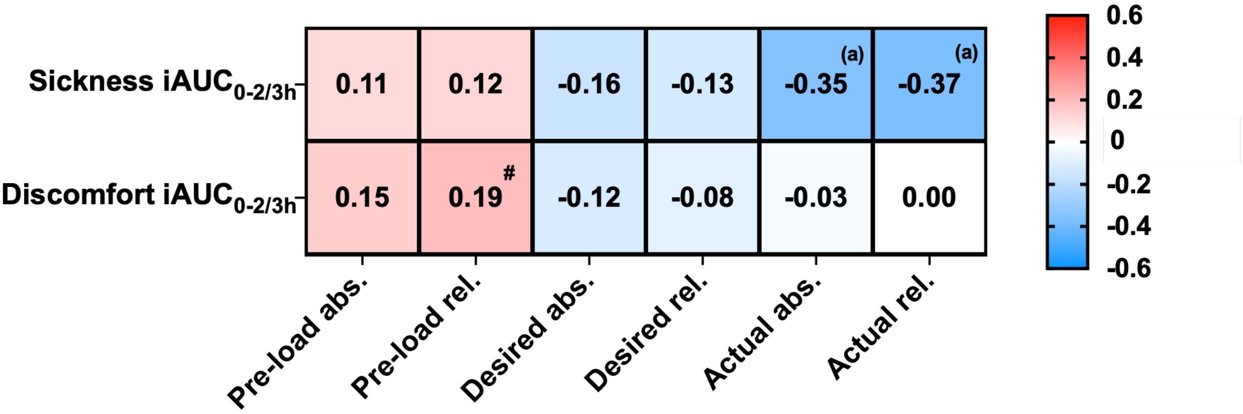

**Supplementary Figure S6. Summary heatmap for correlations of aversive symptoms with outcomes.** The stronger the red, the greater the positive correlation, and the stronger the blue, the greater the negative correlation, with r value and direction given in the cell, with superscript letters giving the P value: [#] P<0.1, [a] P<0.05, [b] P<0.01, [c] P<0.005, [d] P<0.001; letters in brackets indicate P>0.05 using Benjamini-Hochberg false discovery rate (FDR) correction. Abbreviations: abs., absolute kcal; iAUC, incremental area under the curve; rel., relative kcal as % estimated resting energy expenditure. Data from 17 participants, 68 visits for pre-load ($iAUC_{0\text{-}3h}$); 15 participants, 58 visits for desired food intake ($iAUC_{0\text{-}2h}$); 11 participants, 43 visits for actual food intake ($iAUC_{0\text{-}3h}$).

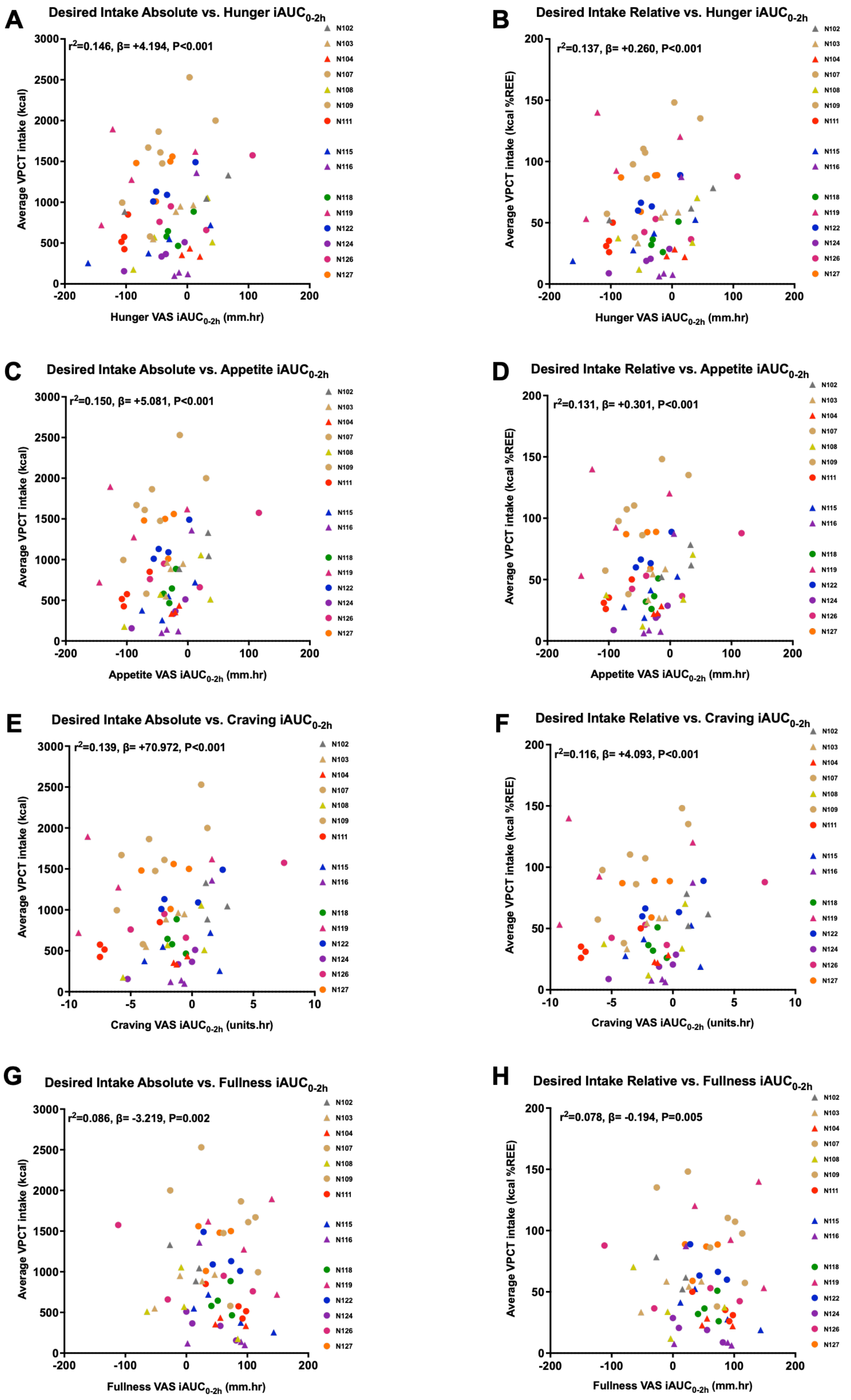

**Supplementary Figure S7. Correlations of appetite ratings with desired food intake.**

Correlations of average (A,C,E,G) absolute (kcal), or (B,D,F,H) relative (kcal %REE) desired food intake from VPCT, with VAS ratings of (A,B) hunger, (C,D) composite appetite, (E,F) food craving, (G,H) fullness, expressed as $iAUC_{0\text{-}2h}$, with marginal value of pseudo-R squared ($r^2$), P value, and estimated mean of β parameter from linear mixed model analysis based on data from 15 participants, 58 visits. Males are represented as circles and females as triangles. Abbreviations: iAUC, incremental area under the curve.

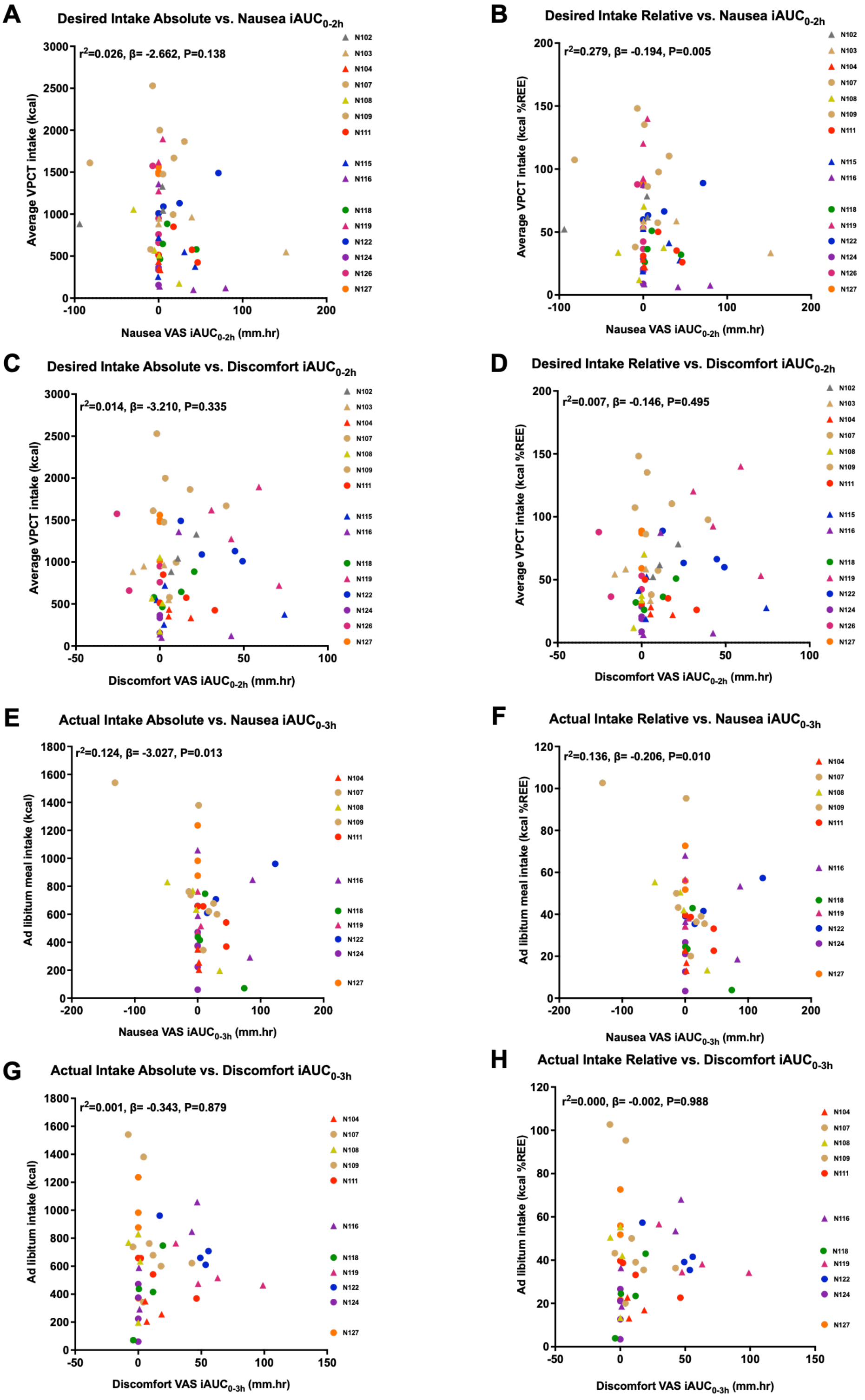

**Supplementary Figure S8. Correlations of aversive symptoms with desired and actual food intake.** Corelations of (A,B,E,F) nausea or (C,D,G,H) discomfort with average (A,C) absolute or (B,D) relative desired food intake at VPCT ($iAUC_{0\text{-}2h}$), or total (E,G) absolute or (F,H) relative actual food intake from *ad libitum* meal ($iAUC_{0\text{-}3h}$), with marginal value of pseudo-R squared ($r^2$), P value, and estimated mean of β parameter from linear mixed model analysis based on data from (A-D) 15 participants, 58 visits and (E-H) 11 participants, 43 visits. Males are represented as circles and females as triangles. Abbreviations: iAUC, incremental area under the curve.

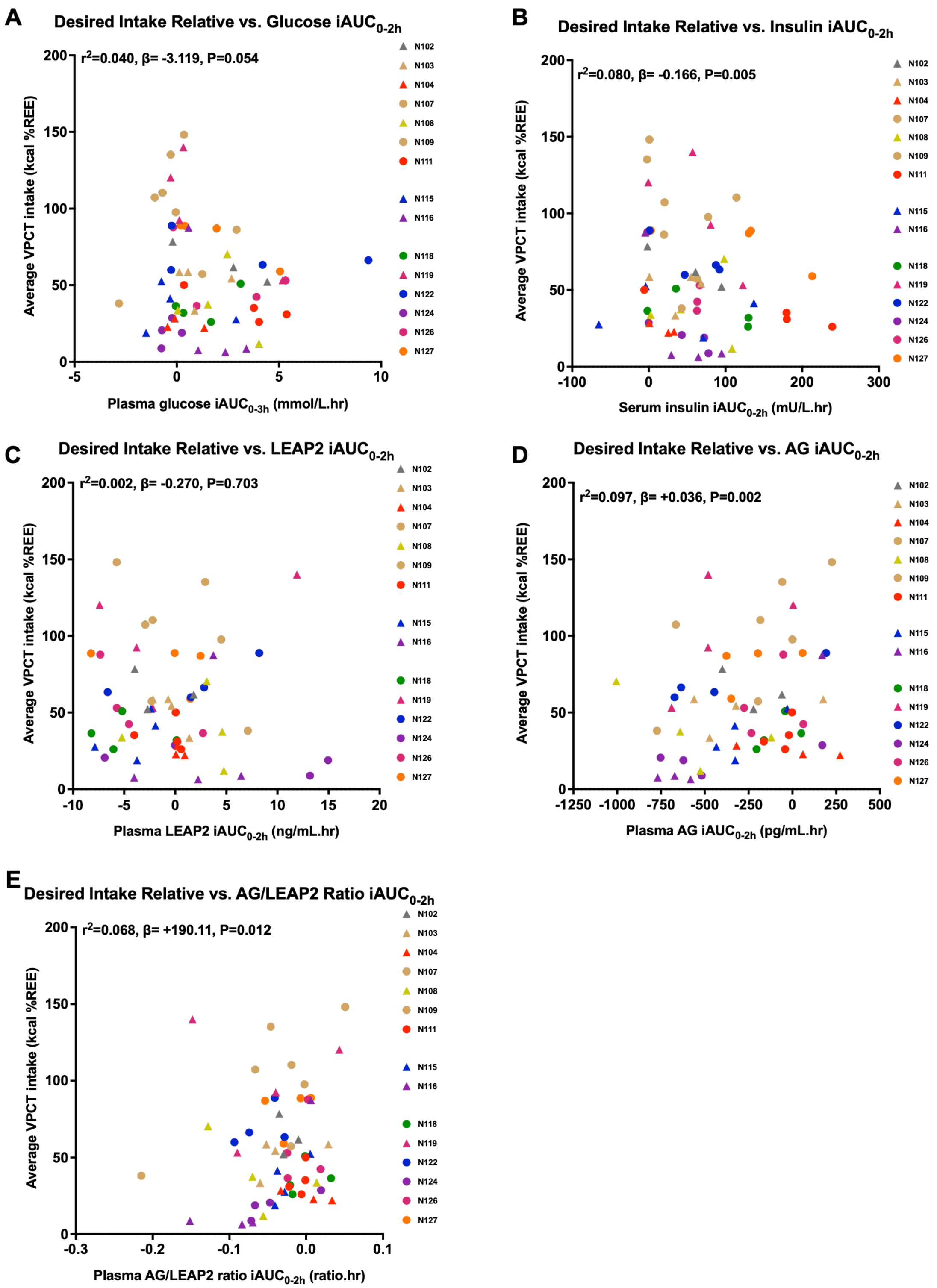

**Supplementary Figure S9. Correlations of blood glucose and hormones with relative desired food intake.** Effects of relative desired food intake (% kcal resting energy expenditure) with (A) plasma glucose, (B) serum insulin, (C) plasma LEAP2, (D) AG and (E) AG/LEAP2 ratio, with marginal value of pseudo-R squared ($r^2$), and estimated mean of β parameter from linear mixed model analysis based on data (A,B) 15 participants, 58 visits, (C-D) 15 visits, 57 visits. Males are represented as circles and females as triangles. Abbreviations: iAUC, incremental area under the curve. To convert glucose from mmol/L to mg/dL multiple by 18.02; to convert insulin from mU/L to pmol/L multiple by 6.94.

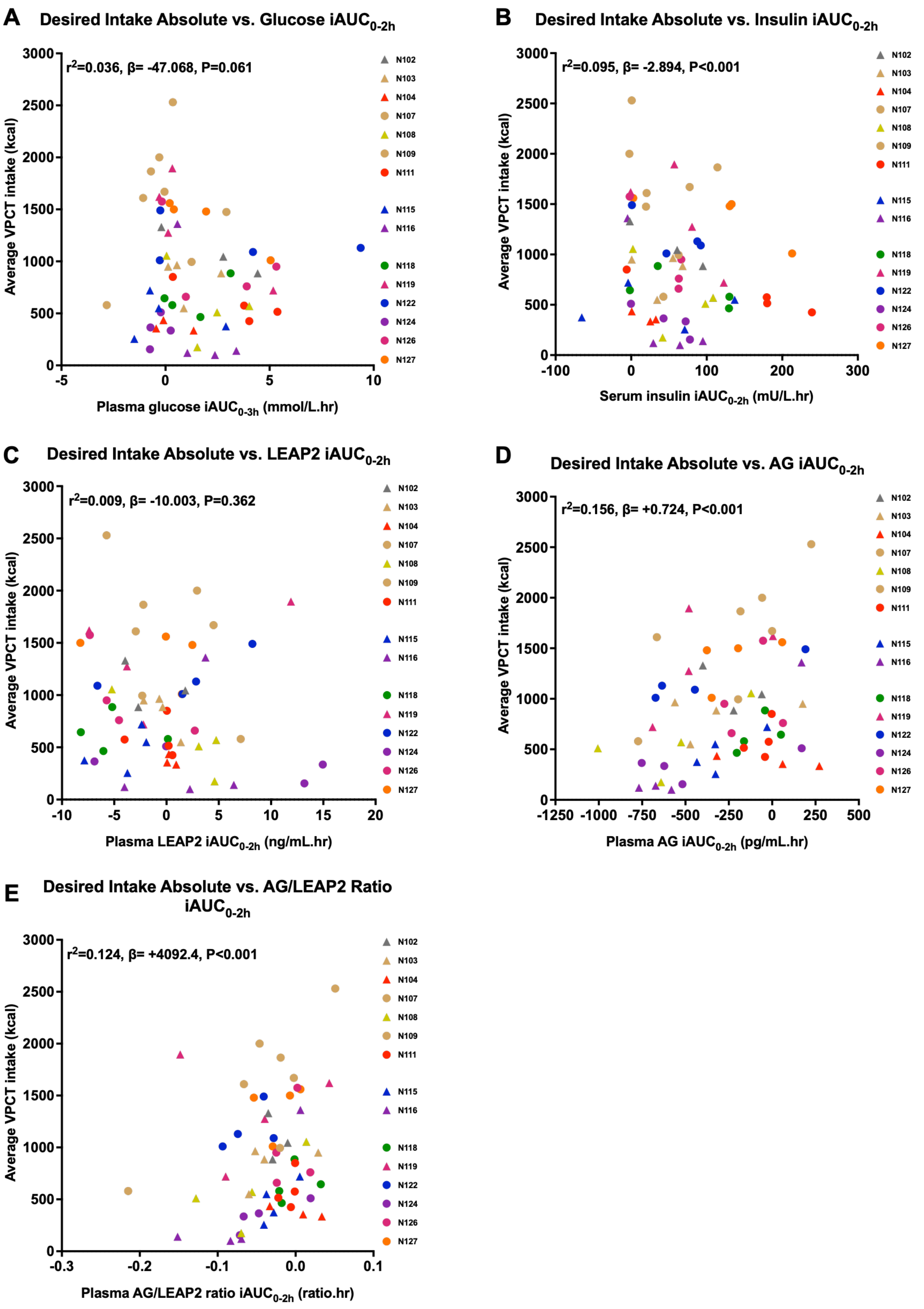

**Supplementary Figure S10. Correlations of blood glucose and hormones with absolute desired food intake.** Effects of absolute desired food intake (kcal) with (A) plasma glucose, (B) serum insulin, (C) plasma LEAP2, (D) AG and (E) AG/LEAP2 ratio, with marginal value of pseudo-R squared ($r^2$), and estimated mean of β parameter from linear mixed model analysis based on data from (A,B) 15 participants, 58 visits, (C-D) 15 visits, 57 visits. Males are represented as circles and females as triangles. Abbreviations: iAUC, incremental area under the curve.

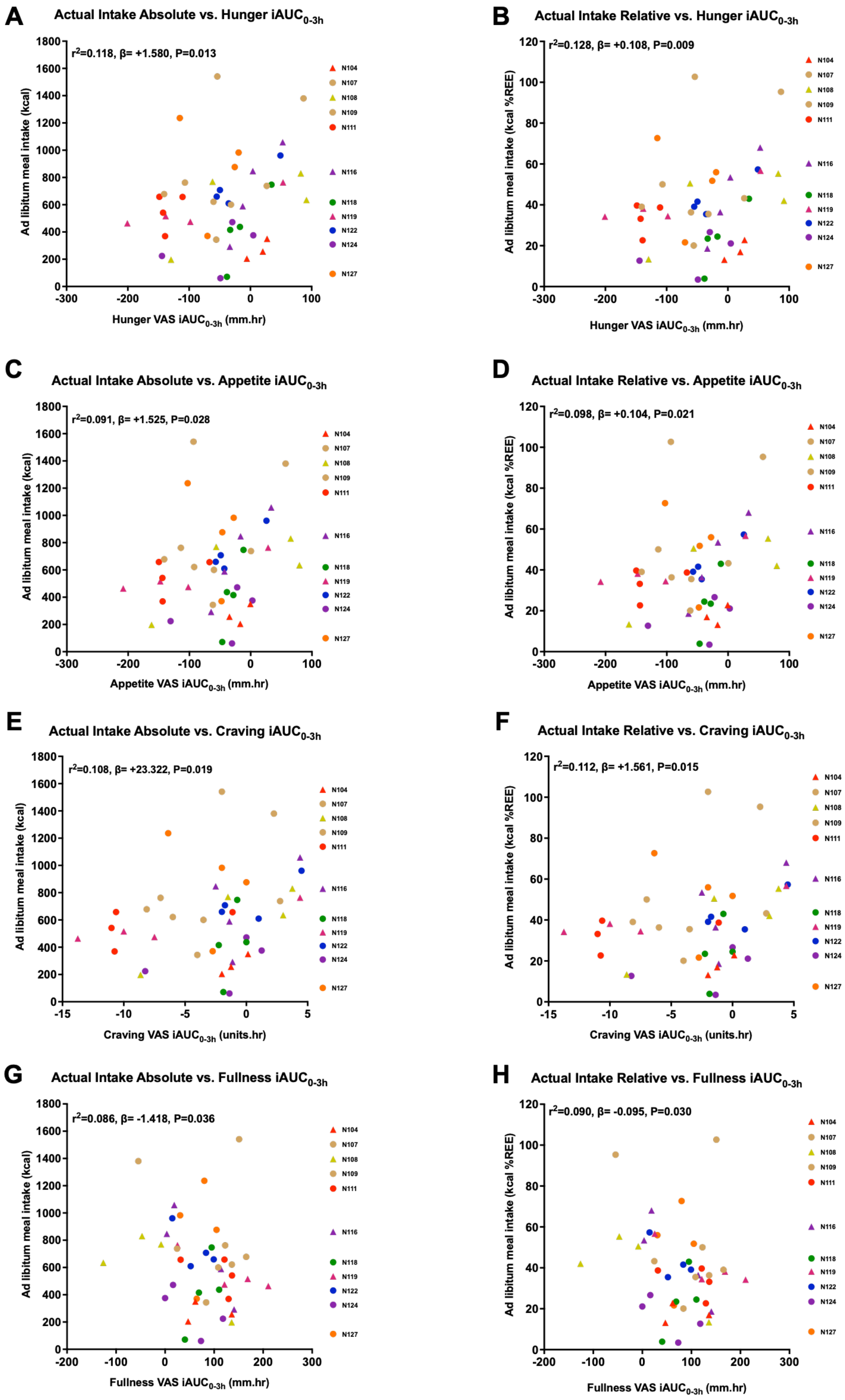

**Supplementary Figure S11. Correlations of appetite ratings with actual food intake.**

Correlations of (A,C,E,G) absolute (kcal), or (B,D,F,H) relative (kcal %REE) average desired food intake from VPCT, with VAS ratings of (A,B) hunger, (C,D) composite appetite, (E,F) food craving, (G,H) fullness, expressed as $iAUC_{0\text{-}3h}$, with marginal value of pseudo-R squared ($r^2$), P value, and estimated mean of β parameter from linear mixed model analysis based on data from 11 participants, 43 visits. Males are represented as circles and females as triangles. Abbreviations: iAUC, incremental area under the curve.

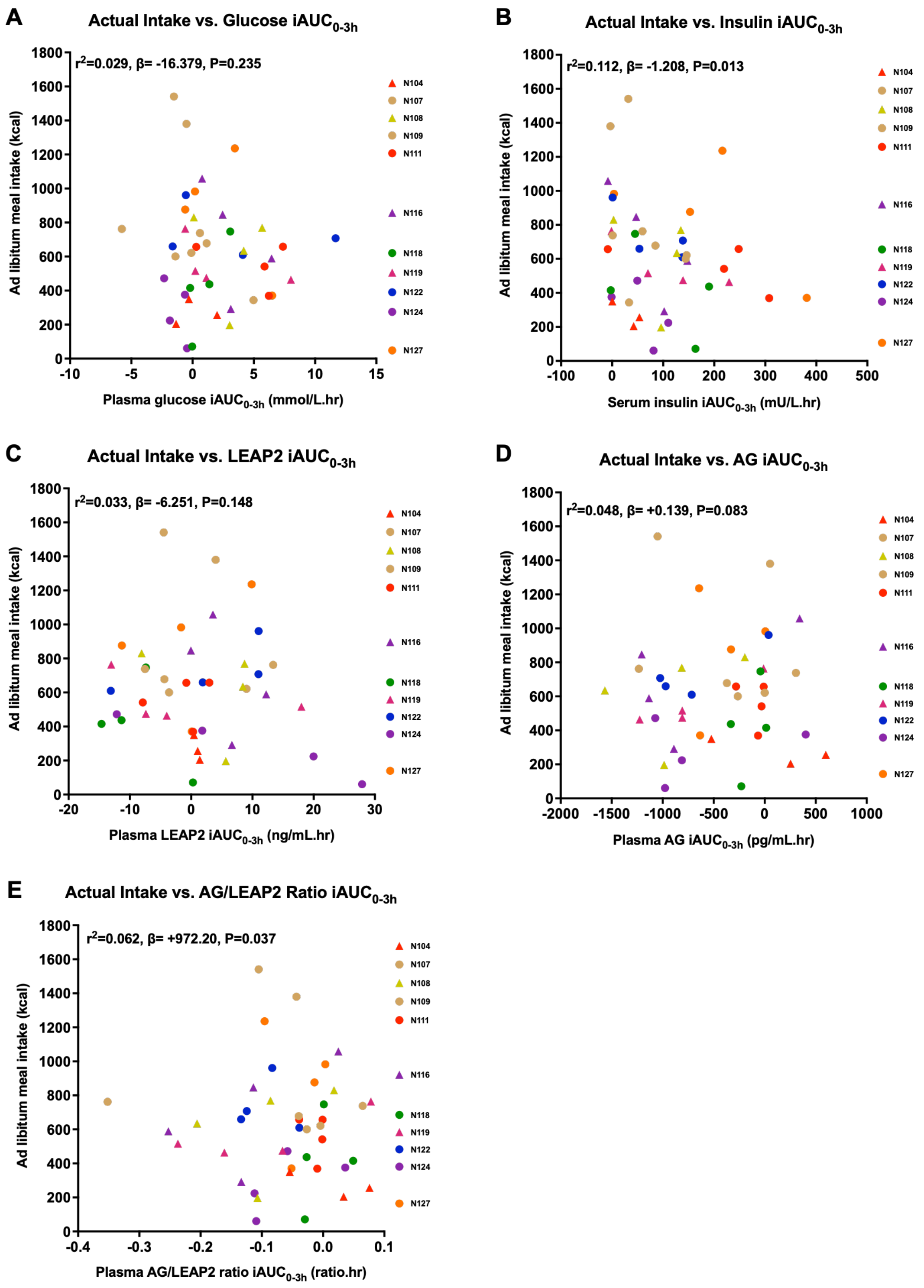

**Supplementary Figure S12. Correlations of blood glucose and hormones with absolute actual food intake.** Effects of absolute actual food intake (kcal) with (A) plasma glucose, (B) serum insulin, plasma (C) LEAP2, (D) AG and (E) AG/LEAP2 ratio, with marginal value of pseudo-R squared ($r^2$), and estimated mean of β parameter from linear mixed model analysis based on data from (A,B) 11 participants, 43 visits, (C-E) 11 participants, 42 visits. Males are represented as circles and females as triangles. Abbreviations: iAUC, incremental area under the curve. To convert glucose from mmol/L to mg/dL multiple by 18.02; to convert insulin from mU/L to pmol/L multiple by 6.94.

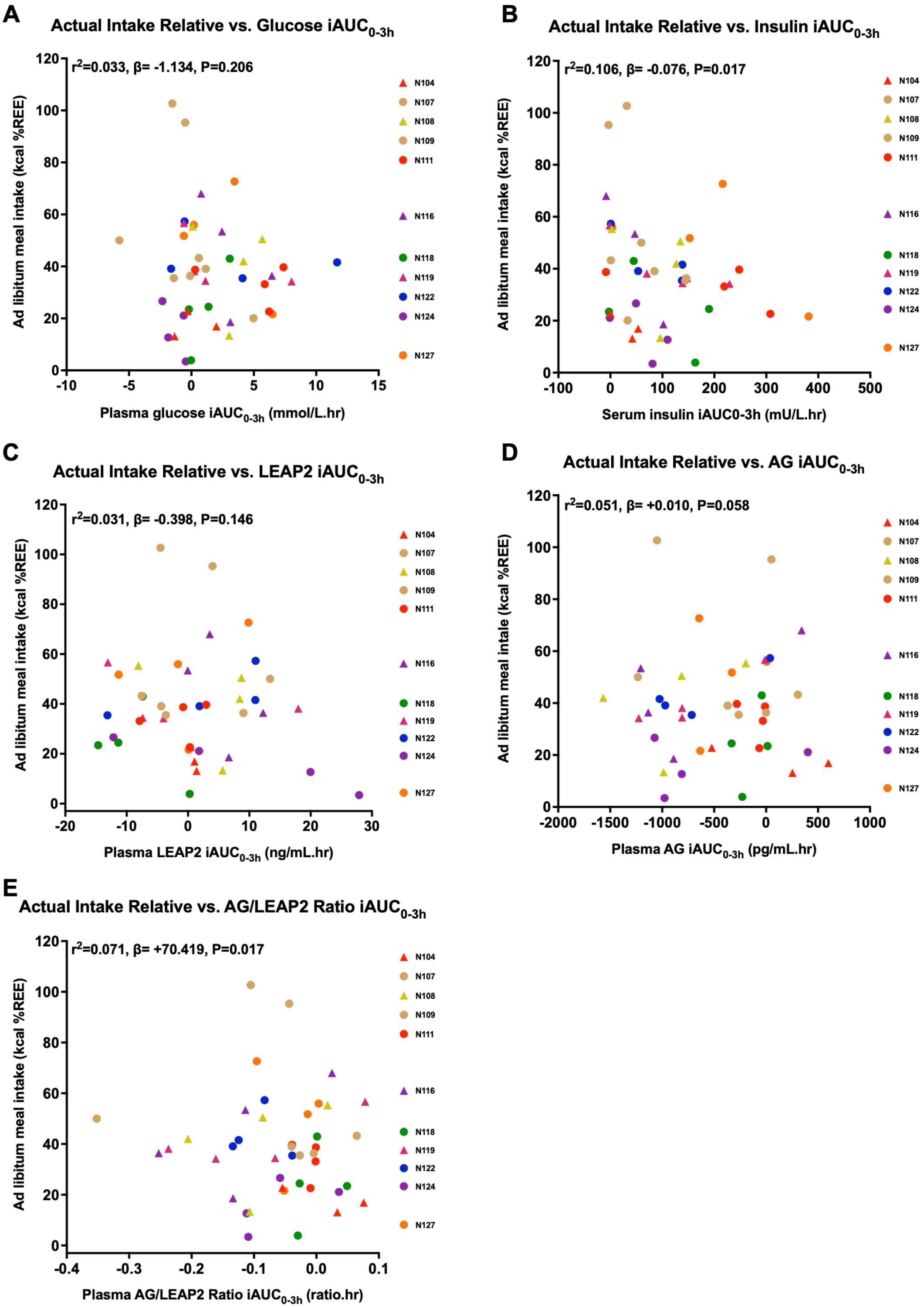

**Supplementary Figure S13. Correlations of blood glucose and hormones with relative actual food intake.** Effects of relative actual food intake (% kcal resting energy expenditure) with (A) plasma glucose, (B) serum insulin, plasma (C) LEAP2, (D) AG and (E) AG/LEAP2 ratio, with marginal value of pseudo-R squared ($r^2$), and estimated mean of β parameter from linear mixed model analysis based on data from 17 participants, 66 visits. Males are represented as circles and females as triangles. Abbreviations: iAUC, incremental area under the curve. To convert glucose from mmol/L to mg/dL multiple by 18.02; to convert insulin from mU/L to pmol/L multiple by 6.94.

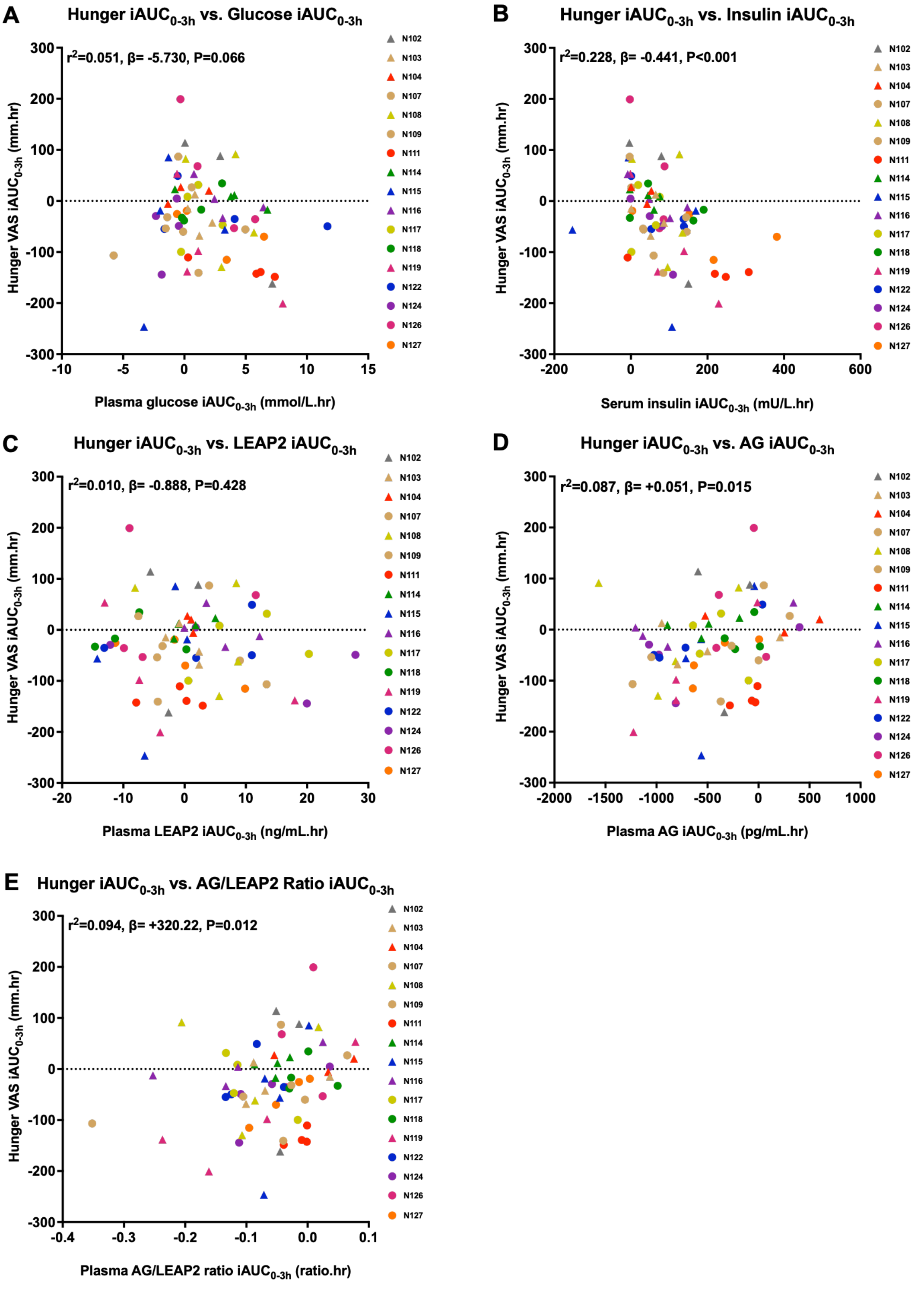

**Supplementary Figure S14. Correlations of blood glucose and hormones with hunger ratings.**

Effects of appetite ratings of hunger (0-100mm) with (A) plasma glucose, (B) serum insulin, plasma (C) LEAP2, (D) AG and (E) AG/LEAP2 ratio, with marginal value of pseudo-R squared ($r^2$), and estimated mean of β parameter from linear mixed model analysis based on data from (A,B) 17 participants, 66 visits, (C-E) 17 participants, 65 visits. Males are represented as circles and females as triangles. Abbreviations: iAUC, incremental area under the curve. To convert glucose from mmol/L to mg/dL multiple by 18.02; to convert insulin from mU/L to pmol/L multiple by 6.94.

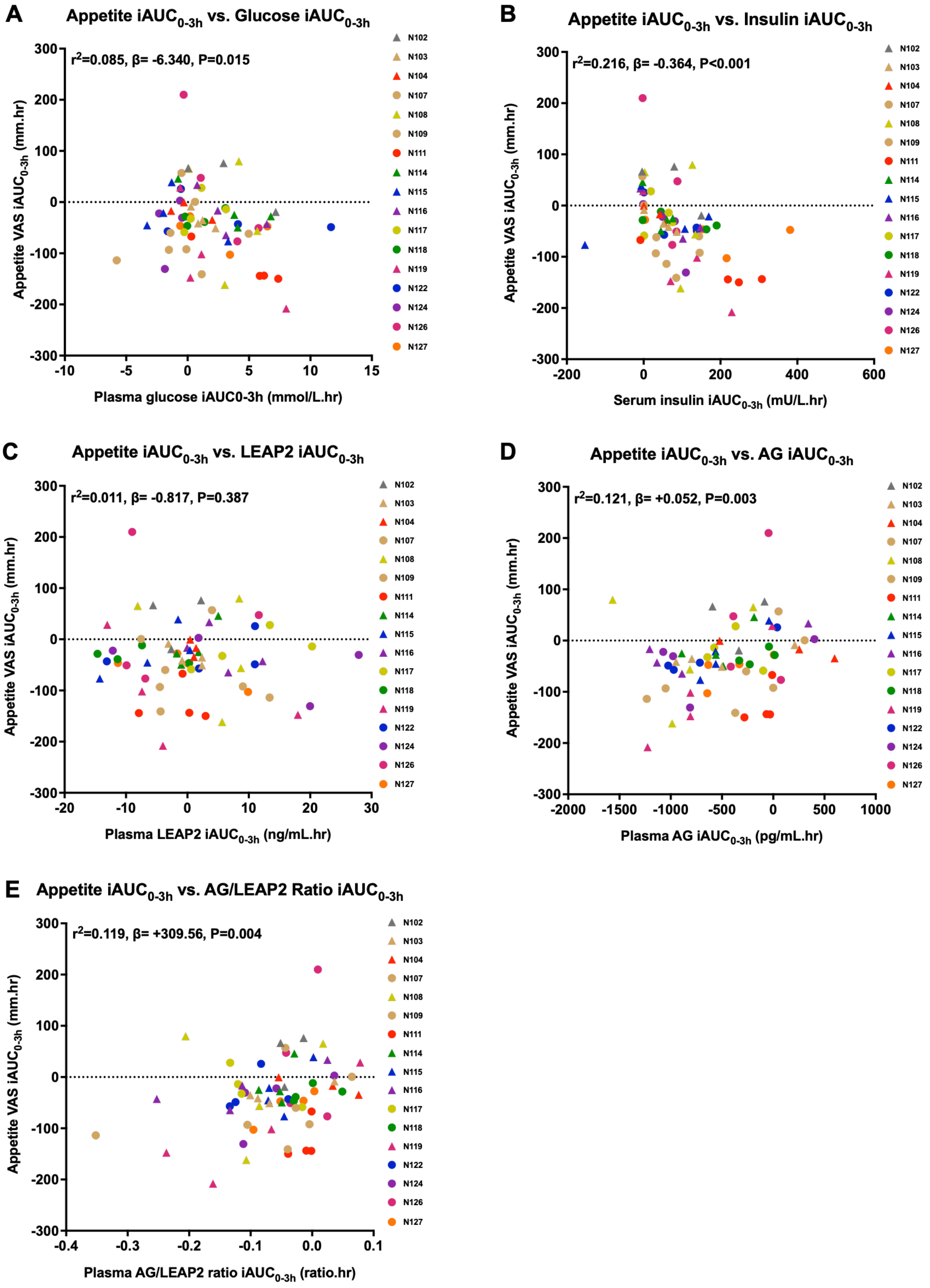

**Supplementary Figure S15. Correlations of blood glucose and hormones with composite appetite ratings.** Effects of composite appetite (0-100mm) with (A) plasma glucose, (B) serum insulin, (C) plasma LEAP2, (D) AG and (E) AG/LEAP2 ratio, with marginal value of pseudo-R squared ($r^2$), P value, and estimated mean of β parameter from linear mixed model analysis based on data from (A,B) 17 participants, 66 visits, (C-E) 17 participants, 65 visits. Males are represented as circles and females as triangles. Abbreviations: iAUC, incremental area under the curve. To convert glucose from mmol/L to mg/dL multiple by 18.02; to convert insulin from mU/L to pmol/L multiple by 6.94.

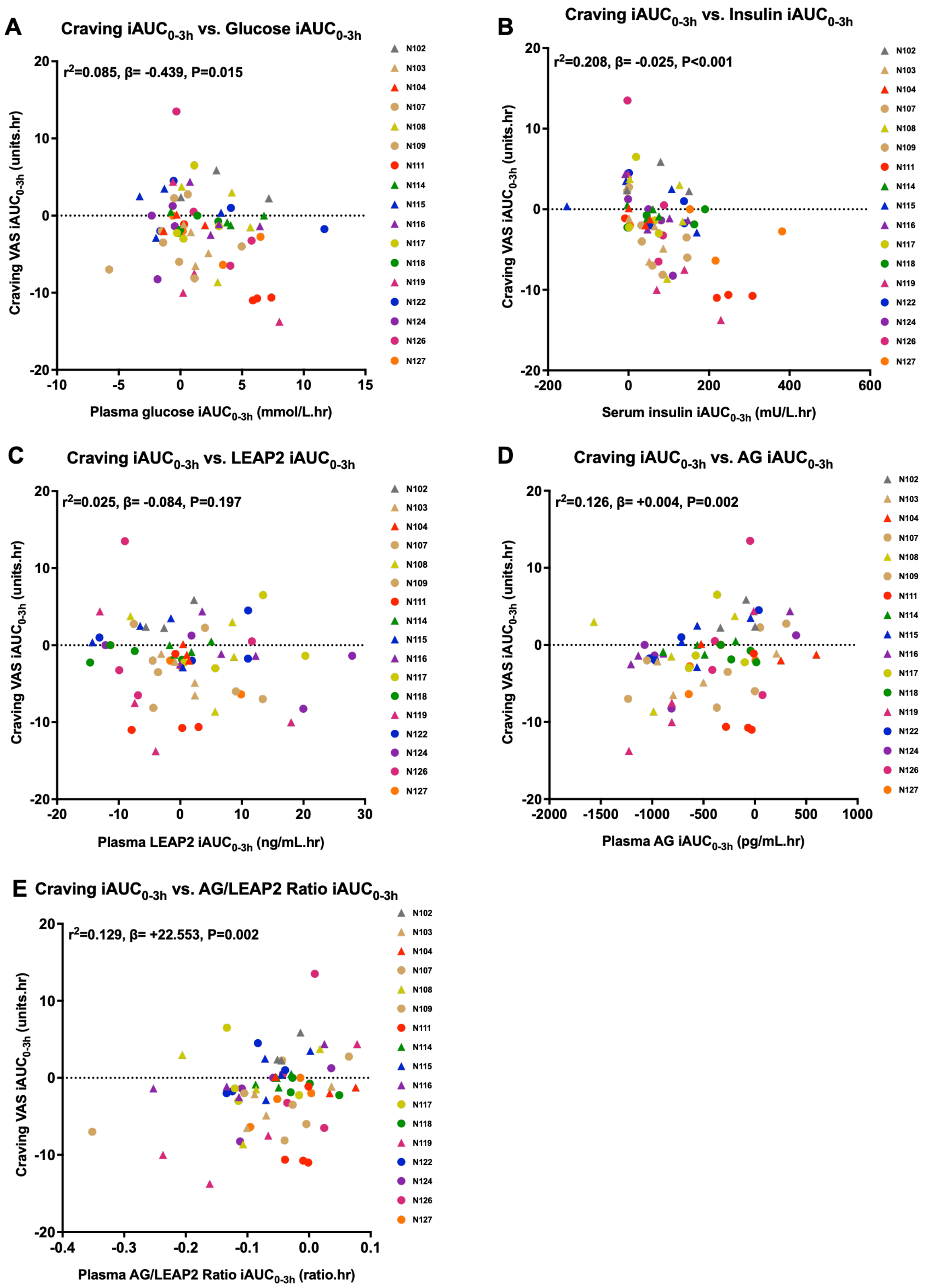

**Supplementary Figure S16. Correlations of blood glucose and hormones with food craving.**

Effects of food cravings (0-100mm) with (A) plasma glucose, (B) serum insulin, (C) plasma LEAP2, (D) AG and (E) AG/LEAP2 ratio, with marginal value of pseudo-R squared ($r^2$), P value, and estimated mean of β parameter from linear mixed model analysis based on data from (A,B) 17 participants, 66 visits, (C-E) 17 participants, 65 visits. Males are represented as circles and females as triangles. Abbreviations: iAUC, incremental area under the curve. To convert glucose from mmol/L to mg/dL multiple by 18.02; to convert insulin from mU/L to pmol/L multiple by 6.94.

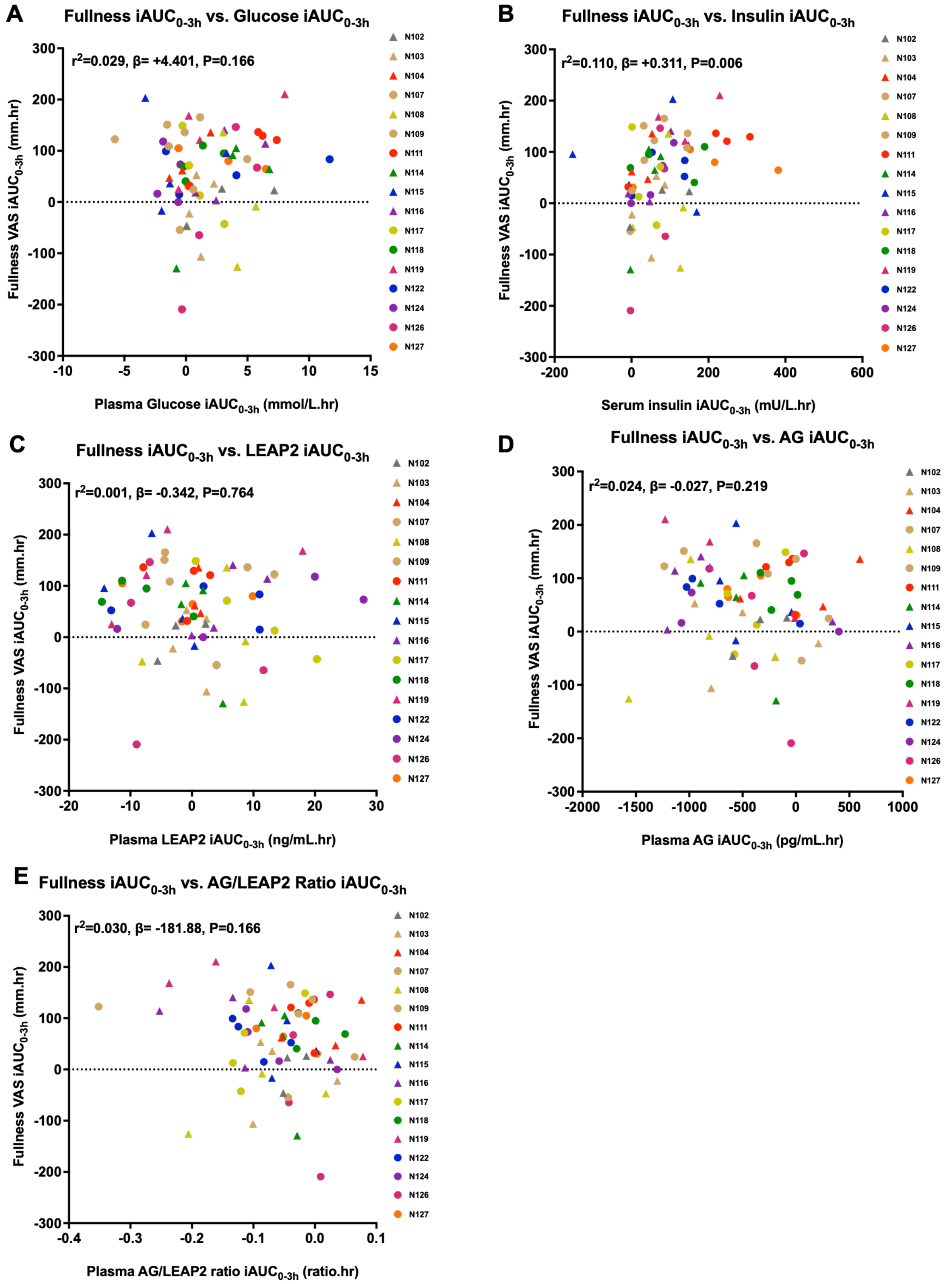

**Supplementary Figure S17. Correlations of blood glucose and hormones with fullness ratings.** Effects of appetite ratings of fullness (0-100mm) with (A) plasma glucose, (B) serum insulin, (C) plasma LEAP2, (D) AG and (E) AG/LEAP2 ratio, with marginal value of pseudo-R squared ($r^2$), P value, and estimated mean of β parameter from linear mixed model analysis based on data from (A,B) 17 participants, 66 visits, (C-E) 17 participants, 65 visits. Males are represented as circles and females as triangles. Abbreviations: iAUC, incremental area under the curve. To convert glucose from mmol/L to mg/dL multiple by 18.02; to convert insulin from mU/L to pmol/L multiple by 6.94.

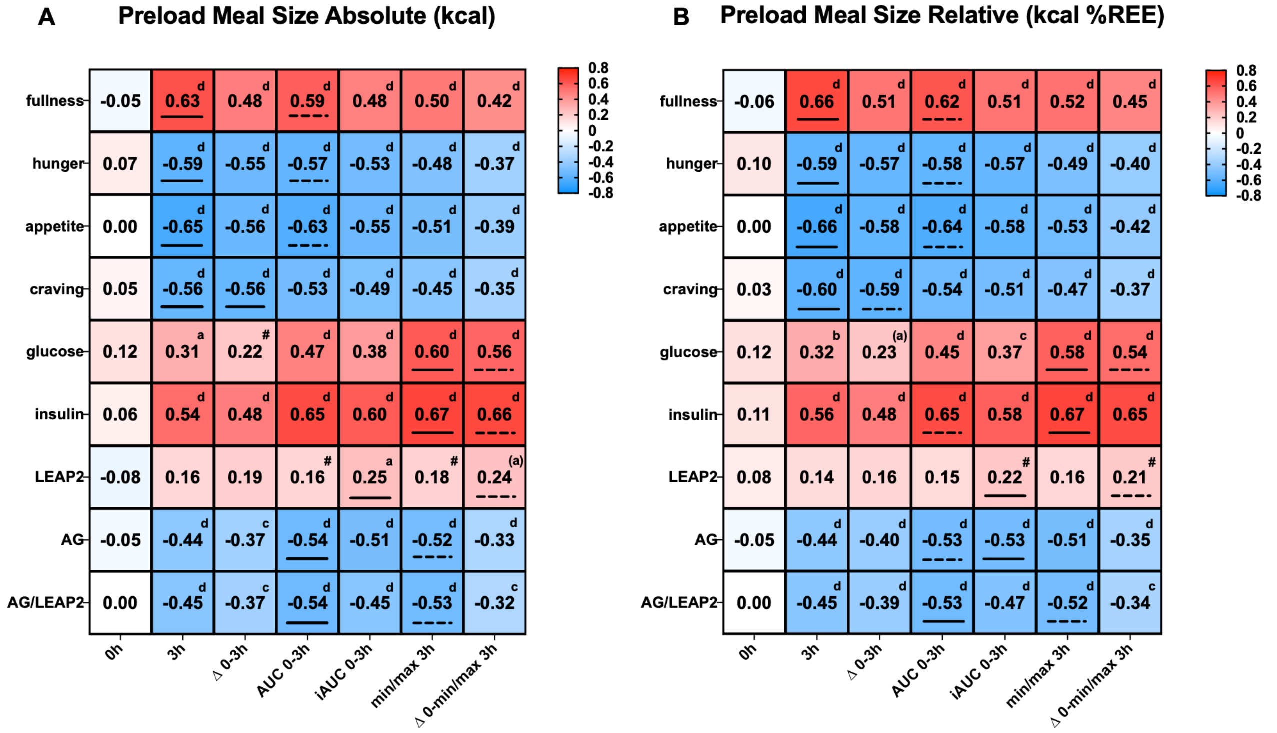

**Supplementary Figure S18. Summary heatmap for correlations of pre-load meal size with outcome measures using exploratory post-prandial calculations.**

Correlations of (A) *absolute* pre-load meal size and (B) *relative* pre-load meal size with visual analogue scale (VAS) ratings of fullness, hunger, composite appetite food craving, plasma glucose, serum insulin, plasma LEAP2, AG, AG/LEAP2 ration using following outcome calculations: 0 hour (baseline), 3 hour, change from 0 to 3 hour, absolute $AUC_{0-3h}$, $iAUC_{0-3h}$, minimum (hunger, composite appetite, craving, AG, AG/LEAP2 ratio) or maximum (fullness, glucose, insulin, LEAP2) after baseline, change from baseline to minimum/maximum. The stronger the red, the greater the positive correlation, and the stronger the blue, the greater the negative correlation, with the r value and direction given in the cell, and the superscript letters giving the P value: [#] P<0.1, [a] P<0.05, [b] P<0.01, [c] P<0.005, [d] P<0.001; letters in brackets indicate P>0.05 using Benjamini-Hochberg false discovery rate (FDR) correction. Abbreviations: Δ, delta; AUC, area under curve iAUC, incremental area under curve.

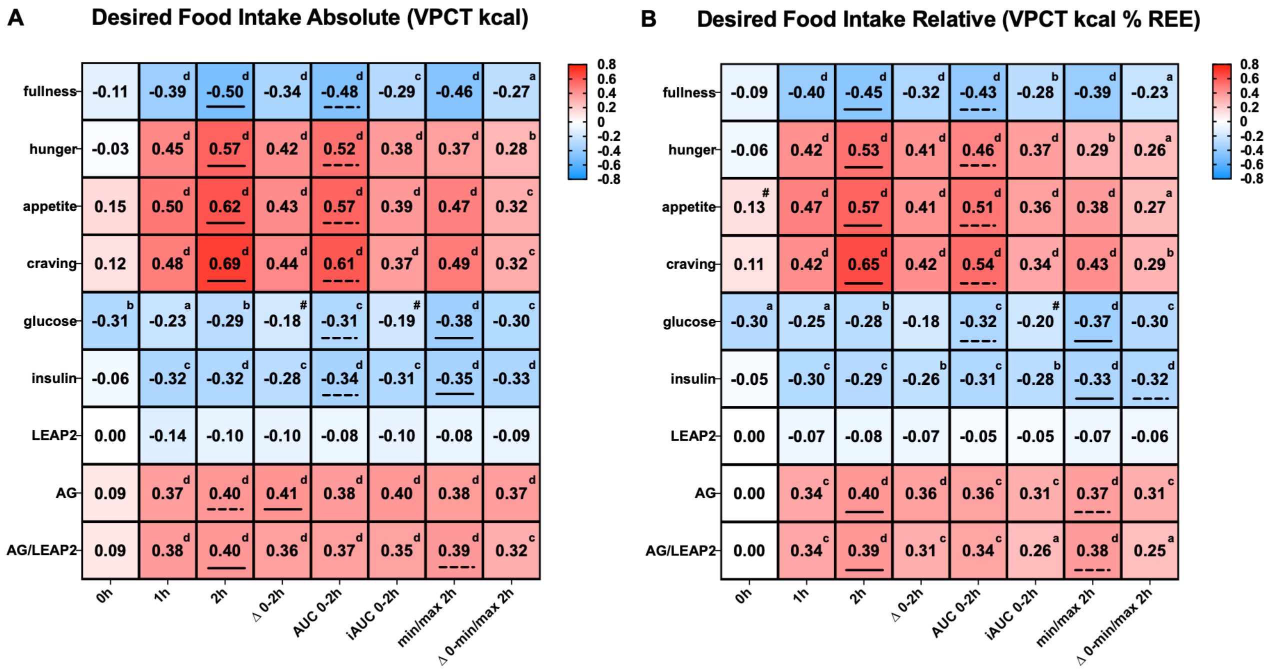

**Supplementary Figure S19. Summary heatmap for correlations of desired food intake with outcome measures using exploratory post-prandial calculations.**

Correlations of (A) *absolute* and (B) *relative* desired food intake with all outcomes of VAS appetite ratings and blood measures. The stronger the red, the greater the positive correlation, and the stronger the blue, the greater the negative correlation, with the r value and direction given in the cell, and the superscript letters giving the P value: [#] P<0.1, [a] P<0.05, [b] P<0.01, [c] P<0.005, [d] P<0.001; letters in brackets indicate P>0.05 using Benjamini-Hochberg false discovery rate (FDR) correction. The solid and dashed underlines indicate the highest and second highest r values respectively. See Supplementary Figure S18 for description of calculations for outcome variables.

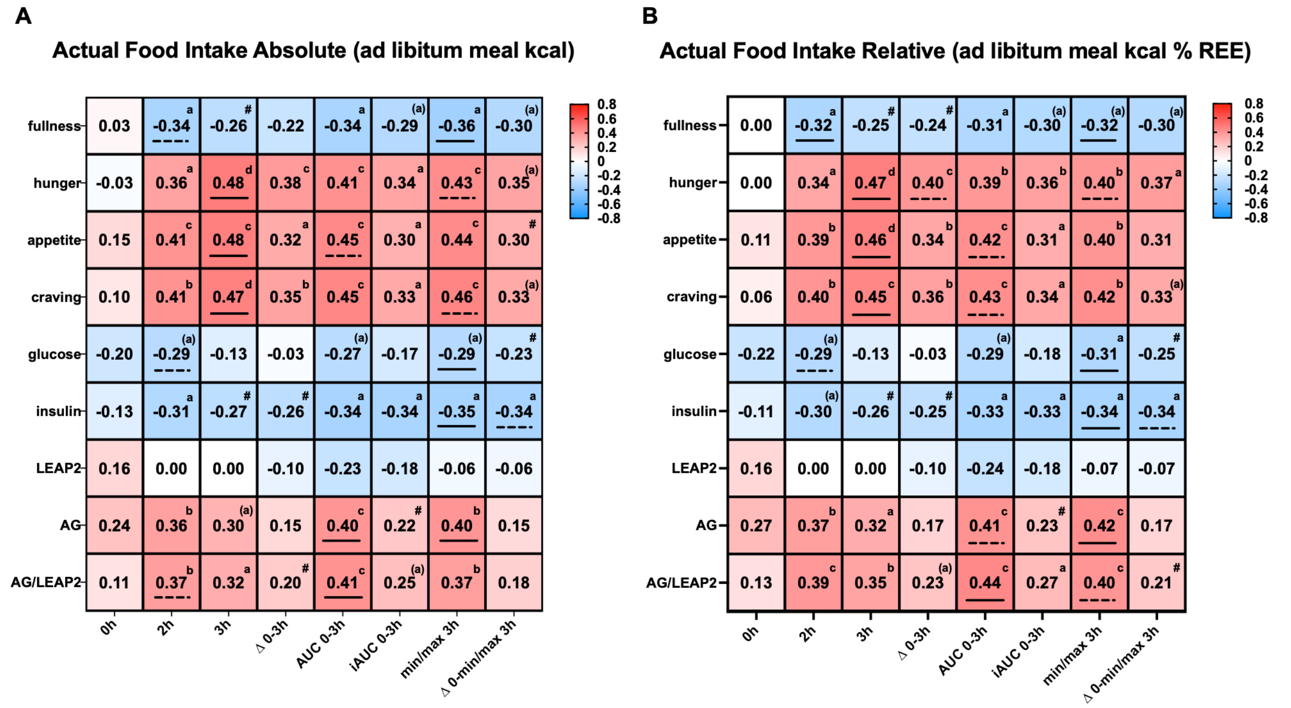

**Supplementary Figure S20. Summary heatmap for correlations of actual food intake with outcome measures using exploratory post-prandial calculations.**

Correlations of (A) *absolute* and (B) *relative* actual food intake with all outcomes of VAS appetite ratings and blood measures. The stronger the red, the greater the positive correlation, and the stronger the blue, the greater the negative correlation, with the r value and direction given in the cell, and the superscript letters giving the P value: $^{\#}$ P<0.1, $^{a}$ P<0.05, $^{b}$ P<0.01, $^{c}$ P<0.005, $^{d}$ P<0.001; letters in brackets indicate P>0.05 using Benjamini-Hochberg false discovery rate (FDR) correction. The solid and dashed underlines indicate the highest and second highest r values respectively. See Supplementary Figure S18 for description of calculations for outcome variables.

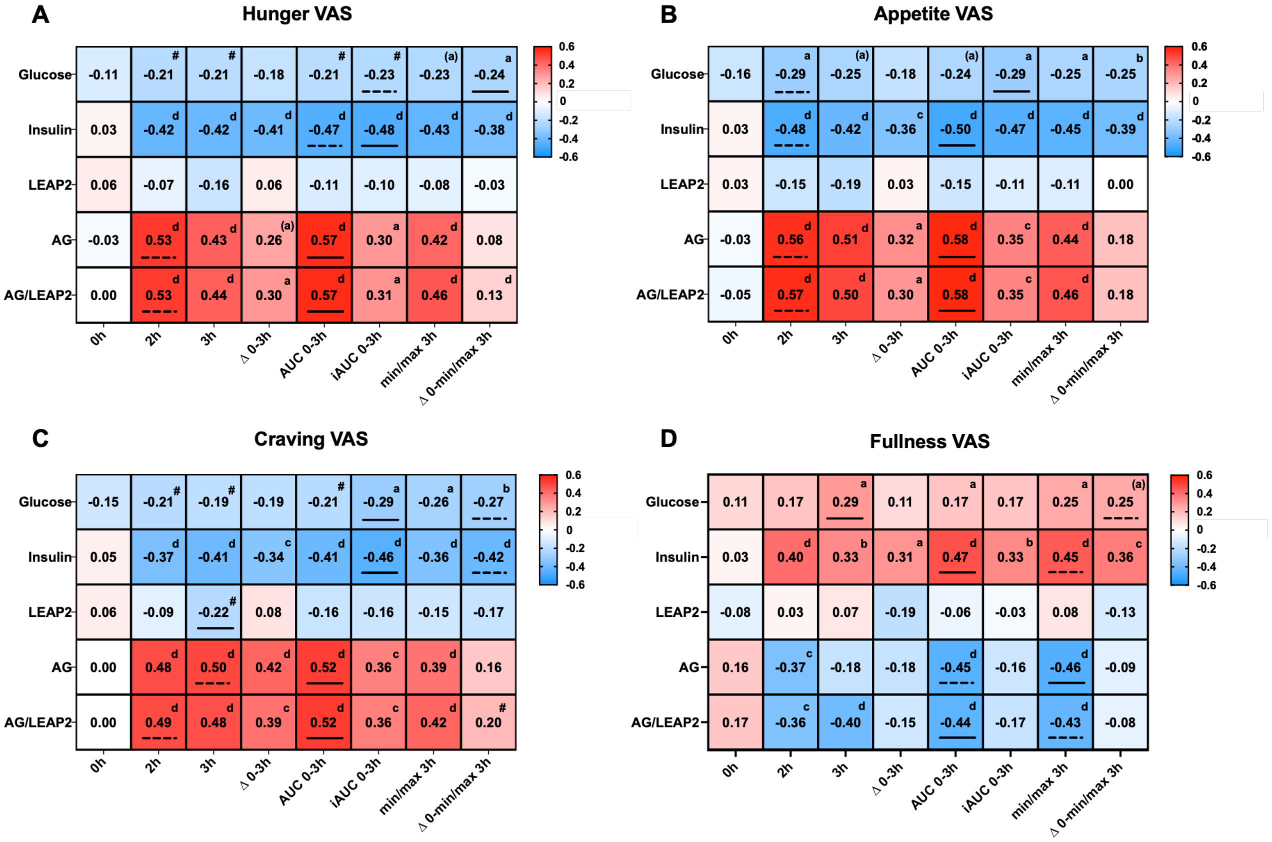

**Supplementary Figure S21. Summary heatmap for correlations of appetite ratings with blood glucose and hormones using exploratory post-prandial calculations.**

Correlations of (A) hunger, (B) composite appetite, (C) food craving, and (D) fullness, with blood measures. The stronger the red, the greater the positive correlation, and the stronger the blue, the greater the negative correlation, with the r value and direction given in the cell, and the superscript letters giving the P value: [#] P<0.1, [a] P<0.05, [b] P<0.01, [c] P<0.005, [d] P<0.001; letters in brackets indicate P>0.05 using Benjamini-Hochberg false discovery rate (FDR) correction. The solid and dashed underlines indicate the highest and second highest r values respectively. See Supplementary Figure S18 for description of calculations for outcome variables.

### SUPPLEMENTARY REFERENCES

American-Psychiatric-Association 2013. *Diagnostic and statistical manual of mental disorders: DSM-5 (5th ed.),* Arlington, VA, USA, American Psychiatric Publishing, Inc.

Beck AT, Steer RA, Ball R, Ranieri W. Comparison of Beck Depression Inventories -IA and -II in psychiatric outpatients. J Pers Assess. 67:588-597, 1996.

Fairburn CG, Beglin SJ. Assessment of eating disorders: interview or self-report questionnaire? Int.J.Eat.Disord. 16:363-370, 1994.

Luck AJ, Morgan JF, Reid F, O'Brien A, Brunton J, Price C, Perry L, Lacey JH. The SCOFF questionnaire and clinical interview for eating disorders in general practice: comparative study. BMJ. 325:755-756, 2002.

Saunders JB, Aasland OG, Babor TF, de la Fuente JR, Grant M. Development of the Alcohol Use Disorders Identification Test (AUDIT): WHO Collaborative Project on Early Detection of Persons with Harmful Alcohol Consumption--II. Addiction. 88:791-804, 1993.

Stunkard AJ, Messick S. The three-factor eating questionnaire to measure dietary restraint, disinhibition and hunger. J Psychosom Res. 29:71-83, 1985.

van Strien T, Frijters JER, Bergers GPA, Defares PB. The Dutch Eating Behavior Questionnaire (DEBQ) for assessment of restrained, emotional, and external eating behavior. Int J Eat Disord. 5:295-315, 1986.
